# A Large-Scale Genome-Wide Gene-Sleep Interaction Study in 732,564 Participants Identifies Lipid Loci Explaining Sleep-Associated Lipid Disturbances

**DOI:** 10.1101/2024.09.02.24312466

**Authors:** Raymond Noordam, Wenyi Wang, Pavithra Nagarajan, Heming Wang, Michael R Brown, Amy R Bentley, Qin Hui, Aldi T Kraja, John L Morrison, Jeffrey R O’Connel, Songmi Lee, Karen Schwander, Traci M Bartz, Lisa de las Fuentes, Mary F Feitosa, Xiuqing Guo, Xu Hanfei, Sarah E Harris, Zhijie Huang, Mart Kals, Christophe Lefevre, Massimo Mangino, Yuri Milaneschi, Peter van der Most, Natasha L Pacheco, Nicholette D Palmer, Varun Rao, Rainer Rauramaa, Quan Sun, Yasuharu Tabara, Dina Vojinovic, Yujie Wang, Stefan Weiss, Qian Yang, Wei Zhao, Wanying Zhu, Md Abu Yusuf Ansari, Hugues Aschard, Pramod Anugu, Themistocles L Assimes, John Attia, Laura D Baker, Christie Ballantyne, Lydia Bazzano, Eric Boerwinkle, Brain Cade, Hung-hsin Chen, Wei Chen, Yii-Der Ida Chen, Zekai Chen, Kelly Cho, Ileana De Anda-Duran, Latchezar Dimitrov, Anh Do, Todd Edwards, Tariq Faquih, Aroon Hingorani, Susan P Fisher-Hoch, J. Michael Gaziano, Sina A Gharib, Ayush Giri, Mohsen Ghanbari, Hans Jörgen Grabe, Mariaelisa Graff, C Charles Gu, Jiang He, Sami Heikkinen, James Hixson, Yuk-Lam Ho, Michelle M Hood, Serena C Houghton, Carrie A Karvonen-Gutierrez, Takahisa Kawaguchi, Tuomas O Kilpeläinen, Pirjo Komulainen, Henry J Lin, Gregorio V Linchangco, Annemarie I Luik, Jintao Ma, James B Meigs, Joseph B McCormick, Cristina Menni, Ilja M Nolte, Jill M Norris, Lauren E Petty, Hannah G Polikowsky, Laura M Raffield, Stephen S Rich, Renata L Riha, Thomas C Russ, Edward A Ruiz-Narvaez, Colleen M Sitlani, Jennifer A Smith, Harold Snieder, Tamar Sofer, Botong Shen, Jingxian Tang, Kent D Taylor, Maris Teder-Laving, Rima Triatin, Michael Y Tsai, Henry Völzke, Kenneth E. Westerman, Rui Xia, Jie Yao, Kristin L Young, Ruiyuan Zhang, Alan B Zonderman, Xiaofeng Zhu, Jennifer E Below, Simon R Cox, Michelle Evans, Myriam Fornage, Ervin R Fox, Nora Franceschini, Sioban D Harlow, Elizabeth Holliday, M. Arfan Ikram, Tanika Kelly, Timo A Lakka, Deborah A Lawlor, Changwei Li, Ching-Ti Liu, Reedik Mägi, Alisa K Manning, Fumihiko Matsuda, Alanna C Morrison, Matthias Nauck, Kari E North, Brenda WJH Penninx, Michael A Province, Bruce M Psaty, Jerome I Rotter, Tim D Spector, Lynne E Wagenknecht, Ko Willems van Dijk, Lifelines Cohort Study, Million Veteran Program, Cashell E Jaquish, Peter WF Wilson, Patricia A Peyser, Patricia B Munroe, Paul S de Vries, W James Gauderman, Yan V Sun, Han Chen, Clint L Miller, Thomas W Winkler, Dabeeru C Rao, Susan Redline, Diana van Heemst

## Abstract

We performed large-scale genome-wide gene-sleep interaction analyses of lipid levels to identify novel genetic variants underpinning the biomolecular pathways of sleep-associated lipid disturbances and to suggest possible druggable targets. We collected data from 55 cohorts with a combined sample size of 732,564 participants (87% European ancestry) with data on lipid traits (high-density lipoprotein [HDL-c] and low-density lipoprotein [LDL-c] cholesterol and triglycerides [TG]). Short (STST) and long (LTST) total sleep time were defined by the extreme 20% of the age- and sex-standardized values within each cohort. Based on cohort-level summary statistics data, we performed meta-analyses for the one-degree of freedom tests of interaction and two-degree of freedom joint tests of the main and interaction effect. In the cross-population meta-analyses, the one-degree of freedom variant-sleep interaction test identified 10 loci (P_int_<5.0e-9) not previously observed for lipids. Of interest, the *ASPH* locus (TG, LTST) is a target for aspartic and succinic acid metabolism previously shown to improve sleep and cardiovascular risk. The two-degree of freedom analyses identified an additional 7 loci that showed evidence for variant-sleep interaction (P_joint_<5.0e-9 in combination with P_int_<6.6e-6). Of these, the *SLC8A1* locus (TG, STST) has been considered a potential treatment target for reduction of ischemic damage after acute myocardial infarction. Collectively, the 17 (9 with STST; 8 with LTST) loci identified in this large-scale initiative provides evidence into the biomolecular mechanisms underpinning sleep-duration-associated changes in lipid levels. The identified druggable targets may contribute to the development of novel therapies for dyslipidemia in people with sleep disturbances.

## Introduction

Low levels of high-density lipoprotein cholesterol (HDL-c), and high levels of low-density lipoprotein cholesterol (LDL-c) and triglycerides (TG) are well-characterized risk factors for atherosclerotic cardiovascular disease^1-4^. High LDL-c and TG concentrations have also been shown to causally impact atherosclerotic cardiovascular disease development^5; 6^. Serum lipid levels are influenced by both environmental and genetic factors^7^, and large-scale efforts have identified hundreds of loci associated with increased lipid levels^8-15^.

Sleep disturbances are increasingly recognized as important modifiable risk factors for various metabolic diseases including atherosclerotic cardiovascular disease and type 2 diabetes^16; 17^. In 2022, sleep duration was added to the Life’s Essentials by the American Heart Association, highlighting the recognition of sleep duration as being important in cardiovascular prevention ^18^. Both short and long self-reported habitual sleep duration have been associated with adverse (atherogenic) lipid profiles in epidemiological cohort studies^19-23^, and recent Mendelian Randomization studies suggest that both short and long habitual sleep durations as potential causal risk factors for atherogenic cardiovascular disease^24-26^. However, despite these findings, the biomolecular mechanisms underpinning sleep-associated atherogenic cardiovascular disease risk are still poorly understood. Examining gene-lifestyle interactions can be an important tool to identify additional genetic variants associated with the trait of interest as well as provide insights into the biomolecular mechanisms underpinning the trait-outcome association^27; 28^. In previously conducted gene-lifestyle interaction projects performed within the Cohort for Heart and Aging Research in Genomic Epidemiology (CHARGE) consortium^29; 30^ Gene-Lifestyle Working Group^27^, we identified multiple loci interacting with lifestyle exposures to lipid levels^31-34^. In particular, we performed a meta-analysis of 126,926 individuals (predominantly European-ancestry; 20% of the participants defined as having either short or long sleep duration), which identified multiple loci associated with lipid profiles in the context of short and long sleep duration. Our results suggested that the effect of long sleep duration and short sleep duration may modify lipid profiles through distinct biological pathways^31^.

In recent years, more data has become available from large biobank initiatives (i.e., UK Biobank and the Million Veteran Program^35; 36^). These data provide an opportunity to increase the sample size substantially in a more diverse study population to allow improved statistical power for the detection of additional gene-by-sleep duration interactions on serum lipid levels. Ultimately, such efforts can improve our understanding of the biomolecular mechanisms underpinning sleep-associated lipid disturbances. Here, we conducted a new and updated multi-population sleep duration-by-gene interaction study on lipid profiles in 732,564 participants from five population groups (African [AFR], East Asian [EAS], European [EUR], Hispanic/Latino [HIS] and South Asian [SAS]).

## Methods

### Overall study design

The study was designed to include cohorts that collected questionnaire-based data on habitual total sleep time and measured blood lipids levels (TG, LDL-c and/or HDL-c). Genome-wide gene×sleep interaction analyses were performed separately by each participating study (and separately for each population group: (AFR, EAS, EUR, HIS, and SAS) following a standardized analysis protocol. Participants 18 years and older were included if they reported a total sleep time between 3 and 14 hours. For studies having habitual total sleep time and lipid levels collected at multiple rounds of visits, the visit with the largest sample size was selected for analysis. Statistical analyses were performed for men and women combined as well as separately for men and women to observe potential effect modification of the variant-sleep interaction effect by sex. Data were subsequently aggregated centrally for quality control and meta-analyses. When applicable, the analysis protocol was reviewed and approved by institutional review boards. Each contributing study was approved by local medical ethics committees and each participant provided written informed consent, in line with the declaration of Helsinki. More information on the individual cohorts is presented in the **Online Supplement**.

### Harmonization of Exposure Variables

Data on habitual total sleep time were collected through questionnaires using questions like “On an average day, how long do you sleep?” to calculate short total sleep time (STST) and long total sleep time (LTST). STST and LTST were derived by regressing sleep time on age, sex, and age×sex, or as indicated otherwise (**Table S2**). The derived residuals’ 20^th^ and 80^th^ percentiles were used as cutoffs: STST=1 if ≤ 20^th^ percentile (otherwise “0”); LTST=1 if ≥ 80^th^ percentile (otherwise “0”).

### Harmonization of Outcome Variables

We considered 3 lipid traits as outcome variables: LDL-c, HDL-c and TG. For most cohorts, fasting (≥8 hours) LDL-c and TG were used. In UK Biobank (N = 359,962 for the combined sample; 49.1% of the total sample) participants were not asked to fast prior to blood samples, and therefore the vast majority (>90%) had no ≥8 hours fasting time. For LDL-c and TG, analyses in UK Biobank were done separately for those meeting the fasting criteria and those who did not, and considered as separate cohorts in subsequent meta-analyses. LDL-c was either directly assayed or derived using the Friedewald equation (the latter restricted to those with TG□≤□400□mg/dL)^37^. LDL-c was corrected for the use of lipid-lowering drugs, defined as any use of a statin drug or any unspecified lipid-lowering drug after the year 1994 (when statin use became common in general clinical practice). If LDL-c was directly assayed, the concentration of LDL-c was corrected by dividing the LDL-c concentration by 0.7. Otherwise (i.e. if LDL-c was derived using the Friedewald equation), then we first divided the concentration of total cholesterol by 0.8 before LDL-c calculationDue to the skewed distribution of HDL-c and TG, we natural log-transformed the concentration prior to the analyses. No transformation for LDL-c was required. All lipid levels were winsorized at 6 standard deviations from the (transformed, if applicable) mean.

### Individual cohort data analyses

Genotype data were restricted to autosomal chromosomes, imputation quality R^2^ ≥0.3 and minor allele frequency (MAF) ≥0.05% (**Table S1**). After data harmonization, each population-group specific cohort ran 2 regression models for 18 phenotype-exposure-sex combinations (3 phenotypes x 2 exposures x All/Men/Women). Below E denotes the sleep exposure (STST or LTST), Y denotes the lipid trait (LDL-c, HDL-c, TG), and C denotes the vector of covariates mentioned above specific to E. Analyses were preferably conducted by each cohort using either of the three software : LinGxEScanR v1.0 (https://github.com/USCbiostats/LinGxEScanR), GEM v1.4.1 (https://github.com/large-scale-gxe-methods/GEM), or MMAP (https://github.com/MMAP/MMAP.github.io) with robust standard errors (SEs) enforced ^38^ (**Table S1**). In cohorts with related participants, null model residuals (regressing lipid traits on a kinship matrix/genetic covariance matrix) were formulated as the lipid outcome.

The two regression models performed included one-degree of freedom (df) tests for examining the variant-sleep interaction effects, and the two-df-joint test that simultaneously assesses the variant-main and variant-sleep interaction effects ^39^. Covariates included population specific principal components of the genotype matrix, cohort-specific confounders (e.g., study center), age, age^2^, sex, age×S/LTST, age^2^×S/LTST, and sex×S/LTST. Finally, for a fair comparison of our results with the previous work (e.g., standard genome-wide association model for comparison ^15^), we also conducted a standard marginal genetic effect model without the consideration of STST or LTST within the same study sample.

### Centralized cohort-level and meta-level quality control

Cohort-level summary statistics were processed centrally. For quality control (QC), we used the EasyQC2 software (www.genepi-regensburg.de/easyqc2) package in R ^40^. Data were filtered for degrees of freedom ≥20 calculated as minor allele count x imputation quality within the unexposed, the exposed, and the total sample. When required, hg37 genomic coordinates were lifted over to hg38 genomic coordinates. Allele frequency discrepancies relative to population-matched TOPMed-imputed 1000G reference panels (Trans-Omics for Precision Medicine imputed 1000Genomes) were assessed, along with genomic control (GC) lambda inflation. Next, meta-level quality control was conducted within population groups (AFR: 13 cohorts, EAS: 5 cohorts, EUR: 30 cohorts, HIS: 7 cohorts, SAS: 1 cohort), with the evaluation of the improper transformation of the outcome variables, unstable numerical computation, or alarming inflation.

### Meta-analyses

Meta-analyses were performed for each population group separately and further combined in a cross-population meta-analyses (CPMA). This resulted in a total of 18 meta-analyses per combination of sleep exposure and lipid trait: five population groups (EUR, HIS, EAS, AFR, SAS) and CPMA, and 3 sex groups (all, women, men). Four tests were considered: the marginal genetic effect (B_M2_G_), the main genetic effect from the interaction model (B_M1_G_), the interaction effect (B_M1_GxE_), and the joint main and interaction effects (B_M1_G,GxE_) with cohort-level GC correction to correct for possible inflation^41^. METAL software for meta-analysis with inverse-variance weights^28^ was used to combine evidence across studies for each of the four tests. CPMA was subsequently executed on the resultant population-specific METAL output results, with population-level GC correction. Due to the low numbers of participants contributing to the HIS, EAS and SAS analyses, these population-specific results were not interpreted seperatly, but only as a part of the CPMA. Furthermore, we only considered variants analyzed in at least 40,000 participants in the main analysis for discovery.

### Identification of independent genomic loci

We used EasyStrata2 software in R to prioritize top loci from significant results identified in the one-df interaction and two-df joint tests^42^. We excluded variants within 1 Mb distance of the major histocompatibility complex (MHC) region. Significant variants were identified using the threshold criteria detailed below. (1) Variants with significant one-df interaction effect (P_int_<5 × 10^−9^, FDR<0.05) and (2) variants with significant two-df joint effect (P_joint_<5 ×10^−9^ with FDR<0.05) were selected as top variants. To prioritize lead variants from the two-df joint analysis with evidence for having variant-sleep interaction, we evaluated the two-df joint lead variants for one-df interactions and used a Bonferroni correction for the number of two-df joint variants identified in the respective population-specific group (CPMA, EUR, AFR) ^43^. Note that the two-df joint test and the one-df interaction effect tests are correlated, so the former procedure does not offer formal statistical evidence of interaction. Nevertheless, it provides a fast and easy prioritization of variants most likely to be involved in interaction with the sleep variables. All such variants were narrowed down to loci based on a 250 kB distance. Finally, within these regions, independent loci were identified by linkage disequilibrium (LD) r^2^ threshold <0.1 using TOPMed-imputed 1000G reference panels. If variants were missing in the LD panels, then the most significant variant within each 500kb region was retained. From the lead variants identified, we additionally extracted the variant information from the sex-stratified analyses to test for heterogeneity of the interaction effects by sex. The heterogeneity of the variant-sleep interaction effect between men and women was tested by performing two-sample Z-tests assuming independence, which were conducted for each interaction loci in the meta-analysis of men and women combined^44^.

### Gene mapping, functional annotation, and follow-up phenotypic annotations

For the lead variants identified, variant mapping was primarily performed using Functional Mapping and Annotation of Genome-wide Association Studies v1.6.0 (FUMA)^45^, and Locuszoom (https://my.locuszoom.org)^46; 47^. At the genomic region level, FUMA’s SNP2GENE pipeline was used to annotate a comprehensive list of genes for each top locus, incorporating genomic position, chromatin interaction (FDR <=1 × 10^−6^, 250bp upstream -500 bp downstream of the transcription startsite [TSS]), and GTEXv8 eQTL evidence with the top variant or its variants in LD (r^2^>0.1 within 500kb)^45; 48^. At the variant level, PheWeb and Open Target Genetics were queried for significant trait associations (p<5 × 10^−8^) from past GWAS analyses^49; 50^. At the gene level, we explored the International Mouse Phenotyping Consortium release 19.1 (IMPC), Online Mendelian Inheritance in Man (OMIM; https://omim.org/), PheWeb, Phenotype-Genotype Integrator (PheGenI), Open Target Genetics, and the online drugbank for retrieving information on the genes as potential drug targets (https://go.drugbank.com)^49-52^. All identified mapped protein-coding genes were then queried using FUMA’s GENE2FUNC pipeline to identify significant (adjusted p-value<0.05) pathways and traits^45^.

### Druggability analysis

We investigated the potential druggability of the sleep duration-lipid trait candidate interacting gene targets as previously described^53^. In short, we first used the Drug-Gene Interaction database (DGIdb; v4.2.0) to query high or medium priority sleep-lipid interacting genes to determine the potential druggability of the candidate gene targets. We annotated genes for implicated pathways and functions using the Kyoto Encyclopedia of Genes and Genomes (KEGG) database. We annotated the druggability target categories and queried all interacting drugs reported in 43 databases (BaderLabGenes, CarisMolecularIntelligence, dGene, FoundationOneGenes, GO, HingoraniCasas, HopkinsGroom, HumanProteinAtlas, IDG, MskImpact, Oncomine, Pharos, RussLampel, Tempus, CGI, CIViC, COSMIC, CancerCommons, ChemblDrugs, ChemblInteractions, ClearityFoundationBiomarkers, ClearityFoundationClinicalTrial, DTC, DoCM, DrugBank, Ensembl, Entrez, FDA, GuideToPharmacology, JACX-CKB, MyCancerGenome, MyCancerGenomeClinicalTrial, NCI, OncoKB, PharmGKB, TALC, TEND, TTD, TdgClinicalTrial, Wikidata). We queried protein targets for available active ligands in ChEMBL. We queried gene targets in the druggable genome using the most recent druggable genome list established from the NIH Illuminating the Druggable Genome Project (https://github.com/druggablegenome/IDGTargets) available through the Pharos web platform (https://pharos.nih.gov/targets). We also queried FDA-approved drugs, late-stage clinical trials and disease indications in the DrugBank, ChEMBL, and ClinicalTrials.gov databases. We provided results for the top MESH and DrugBank indications and clinical trials.

## Results

### Study overview

Data from 55 cohorts including five population groups were included: AFR (13 cohorts, N=48,851 [7%]), EAS (4 cohorts, N=8,097 [1%]), EUR (30 cohorts, N=637,166 [87%]), HIS (7 cohorts, N=32,508 [4%]), and SAS (1 cohort, N=7,619 [1%]). The total sample size was 732,564 participants in the CPMA with 149,210 participants with STST and 147,603 participants with LTST. Additional information on the characteristics of each of study sample is presented in **Tables S1-3**.

### Findings from the one-df variant-sleep interaction analyses

One-df interaction CPMA identified 9 loci displaying evidence for genetic associations with the lipid traits modified by either STST or LTST (*P*_int_ < 5 ×10^−9^ in combination with an FDR < 0.05) (**Figure 1**; **Table 1; Figures S1-3** for -log(P_int_) and QQ plots). Of these, we identified 4 variants for TG, 2 variants for LDL-c and 3 variants for HDL-c. These variants have not been observed before in studies on lipid levels (i.e., ^15^) nor did we find evience of potential variant main effects in the same study sample (**Table S4**). Of the lead variants identified, the 13:50374420:C_T locus (rs14172636; Minor Allele Frequency [MAF] = 0.0087), mapped to the *DLEU1* gene, interacted with STST in its association with both TG (*P*_int_ = 2.40 × 10^−16^) and HDL-c (*P*_int_ = 4.10 × 10^−12^). To illustrate, among those reporting STST, the rs14172636-C allele was associated with 0.26 units lower log-transformed TG (equivalent to an approximate additive decrease of 22.9%) and 0.132 units higher log-transformed HDL-c (equivalent to an approximate additive increase of 14.1%) compared to those without STST. We did not find evidence for a 1-df rs14172636 interaction effect on LDL-c (P_int_ = 0.07). The 8:61617696:C_T locus (rs147261056; MAF: 0.0048), mapped to *ASPH* and *CLVS1*, interacted with LTST in its association with TG (*P*_int_ = 2.78 × 10^−13^). The 11:10411707:C_CT locus (rs1847639939; MAF: 0.46), mapped to the *AMPD3* gene, interacted with LTST in its association with LDL-c (*P*_int_ = 4.72 × 10^−9^). Other variants identified in the CPMA included 3:162278901A_T (rs162278901; *OTOL1*, LDL-c with LTST; *P*_int_ = 2.78 × 10^−13^), 7:72156448:A_G (rs573762901; *CALN1*, HDL with LTST; P_int_ = 1.43 × 10^−10^),2:186808058:G_T (rs6760240; *ZSWIM2*, TG with STST; *P*_int_ = 1.47 × 10^−9^),7:102460277:G_T (rs543672875; *ALKBH4*, HDL-c with STST; *P*_int_ = 1.51 × 10^−9^) and2:184828292:C_T (rs190975828; *ZNF804A*, LDL-c with LTST; P_int_ = 4.72 × 10^−9^).

**Figure 1:**
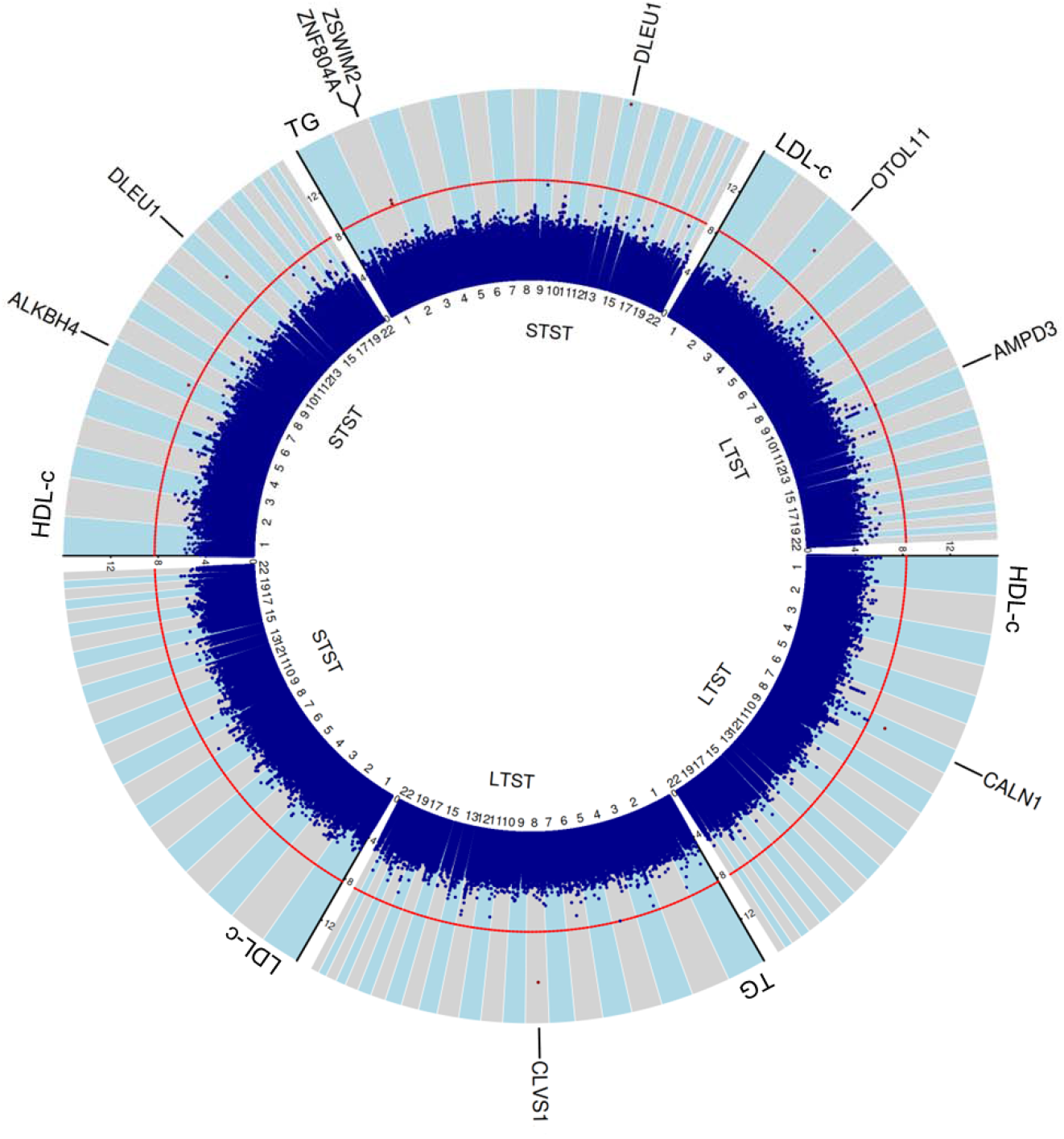
Circular -log10(Pint) plot of all the 6 main analyses in the cross-population meta-analysis of men and women combined. ASPH (TG and LTST) maps also at the *CLVS1* locus.

**Table 1.** Nine variants identified through the 1 degree of freedom interaction analyses in the meta-analyses of men and women combined.

|  |  |  |  |  |  |  |  |  |  |  |  |
| --- | --- | --- | --- | --- | --- | --- | --- | --- | --- | --- | --- |
| Variant | RSid | Effect allele | Exposure | Trait | EAF | Sample Size | Sample | Mapped gene | Interaction Beta | SE | 1df p-value |
| 2:184828292:C_T | rs190975828 | C | STST | TG | 0.9909 | 557,910 | CPMA | *ZNF804A* | -0.102 | 0.0171 | 2.99E-09 |
| 2:186808058:G_T | rs6760240 | G | STST | TG | 0.0075 | 188,049 | CPMA | *ZSWIM2* | 0.184 | 0.0304 | 1.47E-09 |
| 3:162278901:A_T | rs162278901 | A | LTST | LDL-c | 0.0062 | 41,379 | CPMA | *OTOL1*<sup>1</sup> | 25.360 | 3.5968 | 1.78E-12 |
| 4:127678773:C_G | rs192018195 | C | LTST | TG | 0.9849 | 208,087 | EUR | *INTU,*<br>*SLC25A31,*<br>*HSPA4L* | -0.0956 | 0.0145 | 4.81E-11 |
| 7:72156448:A_G | rs573762901 | A | LTST | HDL-c | 0.0033 | 42,445 | CPMA | *CALN1* | 0.148 | 0.0230 | 1.43E-10 |
| 7:102460277:G_T | rs543672875 | G | STST | HDL-c | 0.0024 | 42,445 | CPMA | *ALKBH4* | 0.140 | 0.0231 | 1.51E-09 |
| 8:61617696:C_T | rs147261056 | T | LTST | TG | 0.0071 | 212,110 | CPMA | *CLVS1, ASPH* | 0.185 | 0.0253 | 2.78E-13 |
| 11:10411707:C_CT | rs1847639939 | C | LTST | LDL-c | 0.4588 | 654,182 | CPMA | *AMPD3* | 0.934 | 0.1594 | 4.72E-09 |
| 13:50374420:C_T | rs14172636 | C | STST | HDL-c | 0.9919 | 196,379 | CPMA | *DLEU1* | 0.132 | 0.0190 | 4.10E-12 |
| 13:50374420:C_T | rs14172636 | C | STST | TG | 0.9913 | 188,528 | CPMA | *DLEU1* | -0.266 | 0.0325 | 2.40E-16 |

Abbreviations: CPMA, Cross Population Meta-Analysis; EAF, Effect Allele Frequency; EUR, European; HDL-c, High-Density Lipoprotein
Cholesterol; LDL-c, LowDensity Lipoprotein Cholesterol; SE, standard error; TG, Triglycerides. 1) Identified variant not found in FUMA or in
LocusZoom, *OTOL1* gene is closest gene.

One additional variant was identified in the EUR meta-analysis only. The variant 4:12768773:C_G (rs192018195; *P*_int_ = 4.81 × 10^−11^, MAF = 0.0151) mapping to *INTU/SLC25A31/HSPA4L* was identified in the STST analysis on TG, and was just outside the significance boundaries in the CPMA (*P*_int_ = 5.03 × 10^−9^). Some of the more rare variants identified in these efforts were unable to be investigated further in the population-specific subgroup analyses as variants did not pass post-meta-analysis QC (**Figure 2**). Of the remaining variants, we only found evidence that 11:10411707:C_CT (rs1847639939) was associated with LDL-c in the EUR sample (*P*_int_ = 1.61 × 10^−8^), and not in the AFR meta-analysis (*P*_int_ = 0.74) (**Figure 2**).

**Figure 2:**
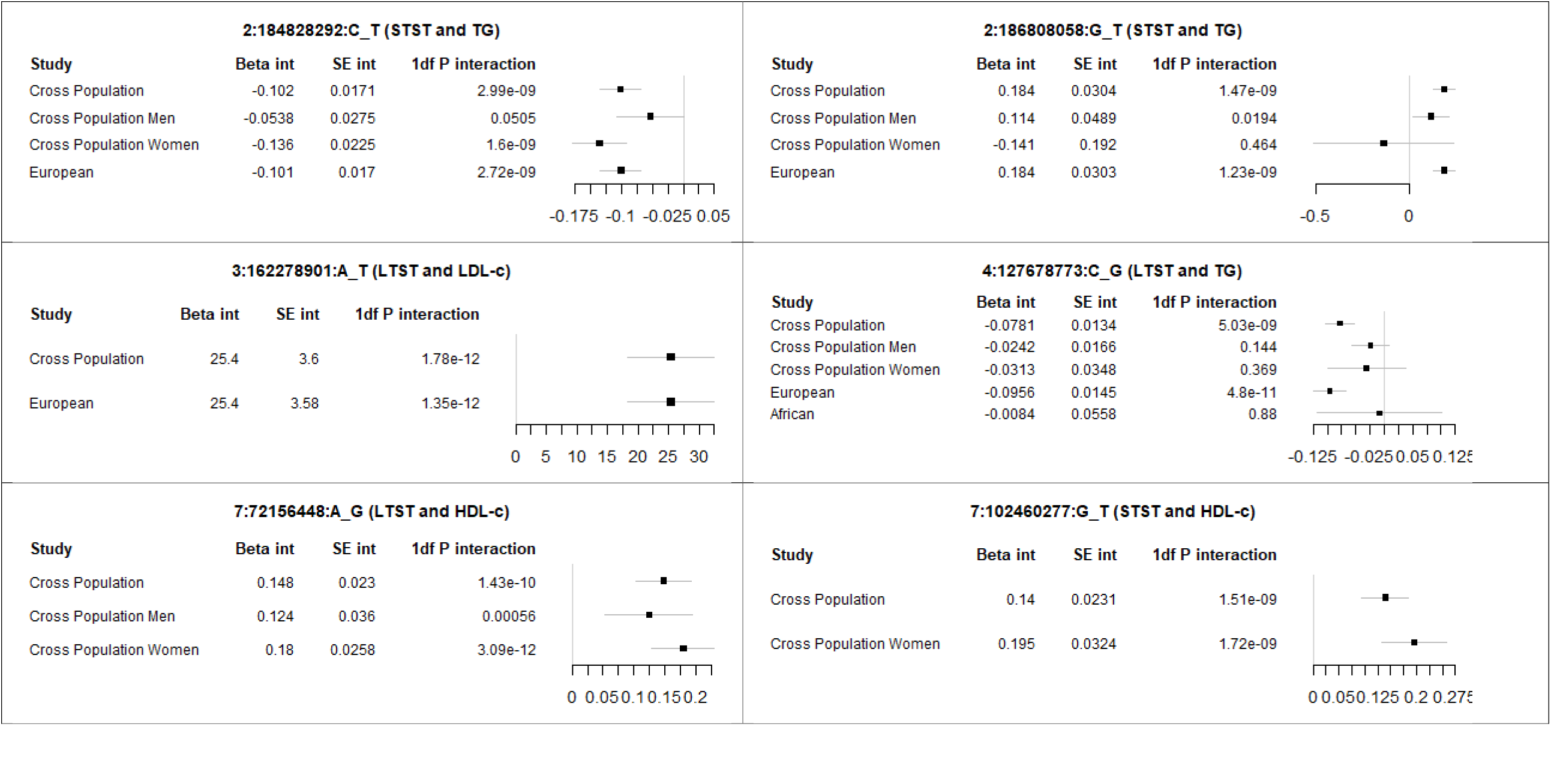
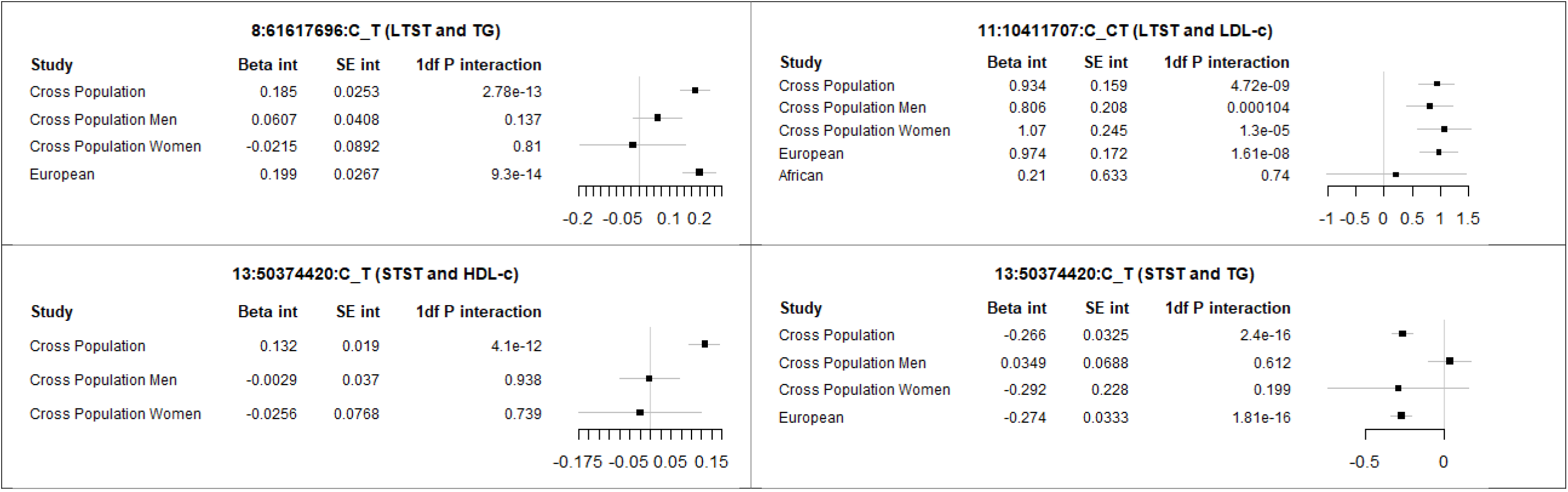
Main results from the 1-degree of freedom interaction analyses in different subgroups. Presented results are the additive variant-interaction effects (log units for TG and HDL-c; mg/dL). Only variants passing all post meta-analysis QC steps were presented. Abbreviations: HDL-c, high-density lipoprotein cholesterol. LDL-c, low-density lipoprotein cholesterol; LTST, long total sleep time; STST, short total sleep time; TG, triglycerides.

An extensive summary of the primary results, including reporting of the results in the sex-specific and population-specific analyses when passing post meta-analysis QC, are presented in **Table S4**; additional information on the region of the identified variants is presented in regional plots presented in **Figure S4**. With the exception of the lead variants mapped to *ASPH* and *DLEU1*, none were noncoding. No additional variants were identified in the sex-stratified analyses nor did we observe evidence for sex differences (*P*_sex-Int_ > 0.05) for variants identified with the one-df interaction test.

### Loci identified through the two-df variant-sleep interaction meta-analyses

Additional analyses were performed to prioritize potential variant-sleep interactions identified by the two-df joint main and interaction effect meta-analyses. In the two-df CPMA (**Table S4 and 5; Figure S5**), we identified (*P*_2df_ < 5 × 10^−9^ and FDR < 0.05) a total of 1,190 lead variants for the TG-LTST analysis (covering 371 genomic regions), 1,156 lead variants for the TG-STST analysis (covering 312 genomic regions), 1,185 lead variants for the HDL-c-LTST analyses (covering 362 genomic regions), 1,178 lead variants for the HDL-c-STST analyses (covering 358 genomic regions), 1,433 lead variants for the LDL-c-LTST analyses (covering 264 genomic regions), and 1,431 lead variants for the LDL-c-STST analyses.

These lead variants were then tested for one-df interaction. Here, we used a less stringent *P*-value cut off for one-df interactions based on the total number of lead variants identified in the CPMA sample for the three traits and two exposure groups combined (*P*_int_ < 6.60 × 10^−6^ = 0.05/7,573, Bonferroni-corrected, see **Methods**). Through this process we identified seven additional genetic lead variants showing evidence for one-df interaction (**Table 2**); of these, five variants were identified for TG (one with LTST, four for STST), one variant for HDL-c (for LTST), and one variant for LDL-c (for LTST) not previously identified for lipid levels nor associated with the lipid trait in the model when not incorporating sleep duration in the same study sample (**Table 2 and Table S4 and S5** and for detailed information). Regional plots of the one-df interaction results of these variants are presented in **Supplementary Figure 8**. In particular, we identified 20:51830403:A_G (rs150607032; *ATP9A/NFATC2/SALL4*, TG with STST, Pint = 3.59 × 10^−8^), 11.13058160:C_T (rs59374498; *TEAD1/RASSF10*, TG with LTST, P_int_ = 5.71 × 10^−8^),10:97769146:A_G (rs191757273; *PYROXD2*, LDL-c with LTST, P_int_ = 7.41 × 10^−8^),21:35272725:A_T (rs114083565; *RUNX1*, TG with STST, P_int_ = 8.40 × 10^−7^),18:55378517:A_T (rs9949541; *TCF4*, HDL-c with LTST), 2:40094191:A_T (rs34771893;*SLC8A1*, TG with STST, P_int_ = 4.12 × 10^−6^), and 20:23353740:A_G (rs73319497;*GZF1/NPAB/CASTL1/CAST11/NXT1*, TG with STST, *P*_int_ = 4.47 × 10^−6^). No evidence was observed that the interaction terms differed for men and women (sex-difference *P*_sex-Int_ > 0.05) (**Supplementary Table 4**). We identified no additional variants among the two-df joint findings showing evidence for one-df interaction (*P*_int_ > 1.10 × 10^−5^ and >1.36 × 10^−4^, respectively; **Table S6 and S7 and Figures S6 and S7**).

**Table 2.** Additional 7 variants identified through the 2 degree of freedom interaction analyses after prioritization for joint effects in the meta-analyses of men and women combined.

|  |  |  |  |  |  |  |  |  |  |  |  |  |
| --- | --- | --- | --- | --- | --- | --- | --- | --- | --- | --- | --- | --- |
| Variant | RSid | Exposure | Effect allele | Trait | EAF | Sample Size | Sample | Mapped gene | 2df joint p-value | Interaction Beta | SE | 1df interaction p-value |
| 20:51830403:A_G | rs150607032 | STST | A | TG | 0.0066 | 532,172 | CPMA | *APT9A, NFATC2, SALL4* | 1.77E-17 | 0.098 | 0.0178 | 3.59E-08 |
| 11:13058160:C_T | rs59374498 | LTST | C | TG | 0.9752 | 661,725 | CPMA | *TEAD1, RASSF10* | 3.34E-38 | -0.0358 | 0.0066 | 5.71E-08 |
| 10:97769146:A_G | rs191757273 | LTST | A | LDL-c | 0.0023 | 52,159 | CPMA | *PYROXD2* | 8.51E-16 | -23.9315 | 4.4475 | 7.41E-08 |
| 21:35272725:A_T | rs114083564 | STST | A | TG | 0.9855 | 43,202 | CPMA | *RUNX1* | 4.45E-29 | 0.1284 | 0.0261 | 8.40E-07 |
| 18:55378517:A_T | rs9949541 | LTST | A | HDL-c | 0.9748 | 71,290 | CPMA | *TCF4* | 2.64E-73 | 0.0478 | 0.01 | 1.92E-06 |
| 2:40094191:A_T | rs34771893 | STST | A | TG | 0.0055 | 557,910 | CPMA | *SLC8A1* | 1.97E-18 | 0.1018 | 0.0221 | 4.12E-06 |
| 20:23353740:A_G | rs73319497 | STST | A | TG | 0.9781 | 666,234 | CPMA | *GZF1, NAPB, CASTL1, CST11, NXT1* | 4.31E-32 | -0.036 | 0.0078 | 4.47E-06 |

Abbreviations: CPMA, Cross Population Meta-analysis; EAF, Effect Allele Frequency; HDL-c, High-Density Lipoprotein Cholesterol; LDL-c,
Low-Density Lipoprotein Cholesterol; SE, standard error; TG, Triglycerides.

### Follow-up analyses

Based on the findings identified in the TG-STST analyses (the lipid-sleep combination with most identified variants in the one-df and two-df interaction analyses), and using the GTEx v8 databases, we did not observe evidence for eQTLs enrichment in any particular tissue (*P* >0.05). Some evidence (p-value = 0.01) was found for enrichment of the Vitamin D receptor pathway (based on the *SLC8A1, NFATC2* and *SALL4* genes; based on Wikipathways using the GENE2FUNC in FUMA ^45^) (**Figure S9**). No evidence for tissue and pathway enrichment was observed for the other loci identified in the exposure-trait combinations.

### Druggability analyses

We first queried mapped gene targets from the different analyses using the Drug-Gene Interaction database (DGIdb), which identified seven genes annotated as clinically actionable or members of the druggable genome (**Table S8a**). Several of these gene targets are implicated in calcium signaling (*SLC25A31, SLC8A1, ASPH*), amino acid or purine metabolism (*PYROXD2, AMPD3*), and regulation of gene transcription (*TEAD1, NFATC2, RUNX1*). We identified seven gene targets of FDA-approved drugs evaluated in late-stage clinical trials using DrugBank and ClinicalTrials.gov databases (**Table S8b**). *SLC8A1* is a target of the nutraceutical icosapent (a modified version of omega-3 fatty acid ethyl eicosapentaenoic acid (EPA)), which is used to treat patients with hypertriglyceridemia. *SLC8A1* is also a target of the small molecule inhibitor caldaret, which was investigated for preventing acute myocardial infarction and treating patients with congestive heart failure.

*SLC8A1* is also a target of FDA-approved antiarrhythmic dronedarone to treat patients with atrial fibrillation or atrial flutter. We also identified *SLC25A31, ASPH*, and *PYROXD2* as targets of commonly prescribed drugs: beta-blocker metoprolol, anticoagulant warfarin, and the attention deficit hyperactivity disorder (ADHD) drug methylphenidate, respectively, all drugs with indications that are frequently observed in people with sleep disorders^54-56^.

## Discussion

This large-scale effort identified several variant-lipid trait associations that were modified by either STST or LTST, without overlap, including 10 loci previously-unidentified in relation to lipid levels that interact with either STST or LTST to blood lipid levels. Using joint meta-analyses, in which the main effect of the variant and the variant-sleep interaction effects are tested jointly, 7 additional genetic lead variants were identified that also showed evidence for interaction with STST or LTST. One of the variants mapped to *DLEU1* and was identified for 2 traits (HDL-c and TG). Moreover, we found distinct variants for STST and LTST interactions– a pattern we also previously reported in a smaller sample for generally higher frequency alleles-suggesting that short and long sleep duration affect the lipid traits through distinct biomolecular mechanisms. Some of the identified genes (most notably *SLC8A1, SLC25A31* and *ASPH*) were previously identified as targets for the prevention or treatment of cardiovascular disease and, therefore show promise as future targets for further validation and clinical translation.

The variants identified in the present study have not been associated previously with sleep duration^57^, other sleep phenotypes (i.e., chronotype, insomnia symptoms or daytime napping)^58-60^, or the blood lipid levels that were considered in the present study ^15^. The majority of the previously unreported findings in this study are low-frequency variants, with the notable exception of 11:10411707C_CT (rs1847639939), that were unlikely to be found in previous studies because they were either not included in the used imputation panels or there was insufficient power. Of the variants identified in the one-df interaction analyses, only the lead variants identified mapped to *ASPH* and *DLEU1* were upstream/downstream transcript variants; all other variants were intronic variants. These findings support the importance of gene-phenotype interaction testing in large studies to explore mechanisms and potential health preventive targets.

A number of the variants identified in the present effort are supported by biological follow-up analyses. Interestingly, we identified *DLEU1* (Deleted In Lymphocytic Leukemia 1), a gene originally identified as a possible tumour suppressor gene and often deleted in patients with B-cell chronic lymphocytic leukemia^61^, in both the variant-STST analyses on HDL-c and TG (and not LDL-c). Previously, genome-wide association studies have also increasingly identified this gene with, amongst others, lipid levels^62^, fatty acid ^63^, anthropometrics^64; 65^, immune markers^66^, and blood pressure^67^. Furthermore, epigenetic changes in peripheral blood in this gene have been identified in acute myocardial infarction^68^. Although *DLEU1* has not been identified with the habitual sleep variables^57-59; 69^, *DLEU1* has been identified to sleep apnea, which is often associated with poor sleep quality and altered sleep duration^70^. We found that the rs14172636 C-allele in *DLEU1* was associated with lower TG and higher HDL-c in individuals reporting short sleep duration, indicative of a lower atherogenic profile. Whether short sleep duration is protective of *DLEU1*-related dyslipidemia, or this variant modifies adverse effects of short sleep duration on lipid levels, cannot be sorted out. The *ASPH* gene was found to be a target for the supplemental Aspartic acid and Succinic acid. Succinate metabolism has been hypothesized as a novel target for myocardial reperfusion injury^71^, and elevated plasma succinate levels have been associated with higher levels of cardiovascular risk factors^72^.

Our druggability analysis results suggest there are potential drug repurposing opportunities to intervene in common signaling and metabolic pathways implicated in sleep behaviour and lipid metabolism, which could help attenuate serious cardiovascular complications in high-risk patients. One of our top plausible gene targets identified in the 2-degree interaction analyses, *SLC8A1*, is targeted by nutraceutical icosapent. Furthermore, *SLC8A1* has previously been described as a target for the investigational drug caldaret. Caldaret, which acts as a cardioprotective drug modulating intracellular calcium levels, has been previously investigated to reduce infarct sizes in patients with acute myocardial infarction, although did not show positive results^73; 74^. Of interest, *SLC8A1* is affected by the renin angiotensin system^75^, which is altered by different sleep conditions^76; 77^. These might present an effective strategy to reduce elevated triglycerides in patients with short sleep duration at risk for cardiovascular complications (e.g., acute myocardial infarction or atrial fibrillation). We also identified several FDA-approved compounds with decades of safe use, which could be evaluated in future preclinical or clinical studies. It is also worth noting the limitations of these predicted drug interactions, which could potentially reflect medication side effects on sleep duration and lipid traits and thus should be interpreted with caution.

We found preliminary evidence for the involvement of the Vitamin D receptor pathway in the association between STST and TG. Although vitamin D itself has not been shown to play any significant role in the onset of cardiovascular disease based on data from randomized clinical trials and Mendelian randomization^78; 79^, the vitamin D receptor appears to be involved in lipid metabolism^80^. Furthermore, genetic variation in the vitamin D receptor gene (*VDR*) has been associated with cardiovascular disease^81^. Accelerated atherosclerosis was observed in *VDR* knock-out mice^82^, suggesting that vitamin D receptor signaling inhibits atherosclerosis development. Finally, vitamin D levels have been reported to vary with various sleep outcomes^83^, and vitamin D supplementation has been hypothesized to improve sleep^84^. Nevertheless, the role of the vitamin D receptor in the association between sleep and lipid disturbances should be explored in greater detail.

The present study used the largest study sample possible, by considering as many cohorts as possible with available data on genomics, self-reported sleep duration, and concurrent lipid levels. Furthermore, we attempted to standardize the self-reported dichotomous sleep-exposure variables as much as possible by first taking the age- and sex-adjusted residuals of total sleep time. Despite our efforts to increase sample size in combination with increased ancestry diversity compared with our previous effort ^31^, the vast majority of our study still consisted of cohorts with mainly EUR participants. It is very likely that population-specific variant-sleep interactions were missed in the meta-analyses of the non-European populations due to a lack of sufficient statistical power; furthermore, because of low statistical power, we did not present the results from the Hispanic and Asian specific meta-analyses. Future efforts in non-European cohorts, when more data become available, should be further expanded. Although some of the identified loci, despite having low allele frequencies, had some evidence of biological plausibility, they should be further explored in independent samples as we did not have the power to separate cohorts into discovery and independent replication analyses. The present study used information on habitual sleep duration collected through self-report, which may have measurement error, possibly resulting in lower statistical power. Note that phenotypic and genetic correlations between sleep duration assessed through questionnaire and accelerometry are low to modest at most^57; 69; 85^, which suggests phenotypes derived by these methodologies reflect different sleep aspects. Finally, the present study considered sleep as a single dimension, whereas sleep is largely acknowledged to be highly dimensional and complex^86^. Indeed, joint associations between sleep duration and sleep quality have been observed in relation to atherosclerotic cardiovascular disease^26; 87; 88^. However, detailed data on sleep quality measures were not available in many cohorts, nor was it possible to harmonize these measures when available. Identified variants should, therefore, also be explored in independent samples, as they become available with other sleep variables.

In summary, the present study identified several novel genetic loci associated with lipid traits that were modified by self-reported short- and long total sleep time. The findings yield new insights into the biology underpinning the observed (causal) association between sleep duration and atherosclerotic cardiovascular disease. The observed targets for treatment yield insights into possible prevention of atherosclerotic cardiovascular disease in relation to sleep duration. Additional functional follow-up is required to further characterize the identified genetic variants and to translate the findings to more biological and clinical context.

## Data Availability

All data produced in the present study are available upon reasonable request to the authors

## ACKNOWLEDGMENTS

This project was largely supported by two grants from the U.S. National Heart, Lung, and Blood Institute (NHLBI), the National Institutes of Health, R01HL118305 and R01HL156991. Furthermore, RN and DvH were supported by a grant from the Dutch Research Council (NWO, Dutch National Research Agenda, Research along routes by consortia, 2021–2026, BioClock: the circadian clock in modern society). This research was supported in part by the Intramural Research Program of the National Human Genome Research Institute in the Center for Research on Genomics and Global Health (CRGGH— Z01HG200362). HW and PN were supported by R01HL153814 and R21HL165324. CRGGH is also supported by the National Institute of Diabetes and Digestive and Kidney Diseases (NIDDK), the Center for Information Technology, and the Office of the Director at the National Institutes of Health. DAL and QY contributions were supported by the British Heart Foundation (CH/F/20/90003) and UK Medical Research Council (MC_UU_00032/05). TOK was supported by grants from the Novo Nordisk Foundation (NNF18CC0034900, NNF21SA0072102, NNF22OC0074128, NNF23SA0084103). PBM acknowledges support from the National Institute for Health and Care Research (NIHR) Biomedical Research Centre at Barts (NIHR202330). WJG and JM were supported by PO1CA196569. Study-specific funding information is provided in the online supplement.

## Declaration of interests

HJG has received travel grants and speakers honoraria from Neuraxpharm, Servier, Indorsia and Janssen Cilag. LMR is a consultant for the TOPMed Administrative Coordinating Center (through Westat). TDS is co-founder and shareholder of ZOE Ltd. All other co-authors declare to have no conflicts of interest.

## Notes

### Author Declarations

When applicable, the analysis protocol was reviewed and approved by institutional review boards. Each contributing study was approved by local medical ethics committees and each participant provided written informed consent, in line with the declaration of Helsinki. More information on the individual cohorts is presented in the Online Supplement.

### Summary of Updates

Added Million Veteran Program in the author banner. Updated their Acknowledgements.

