## Supplementary Figures for "A Large-Scale Genome-Wide Gene-Sleep Interaction Study in 732,564 Participants Identifies Lipid Loci Explaining Sleep-Associated Lipid Disturbances"

**Supplementary Table 1: -log(p) and accompanying Q-Q plots for the combined one-df sleep interaction analyses (CPMA)**

| HDL-c and LTST |  |
| --- | --- |
| 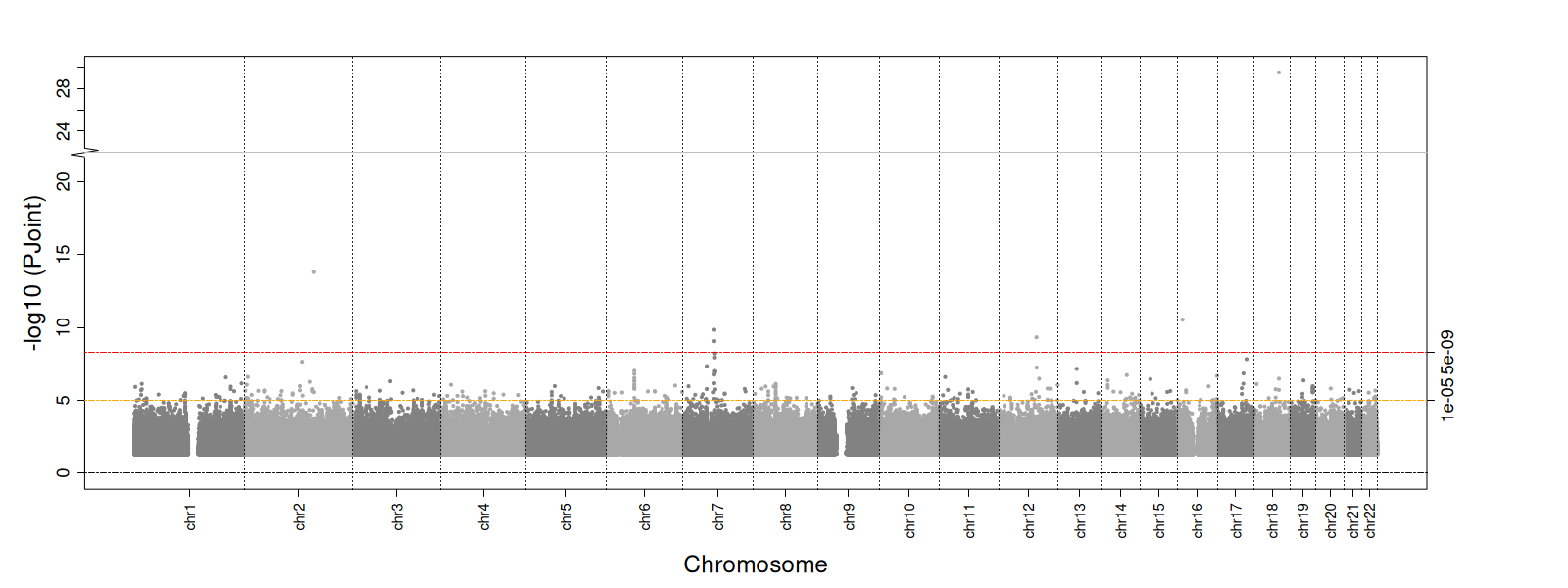 | 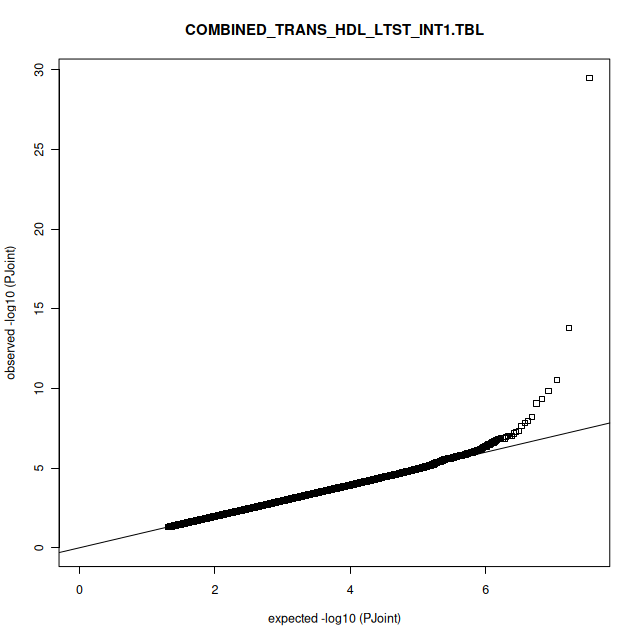 |
| HDL-c and STST |  |
| 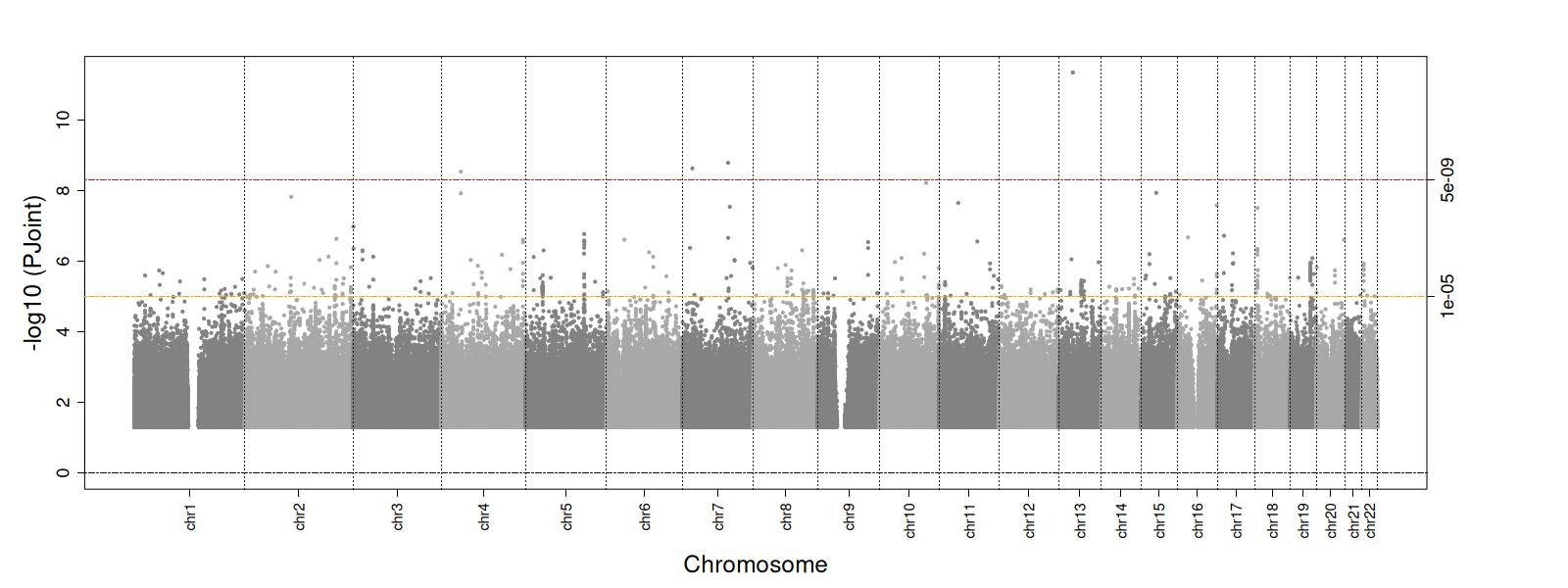 | 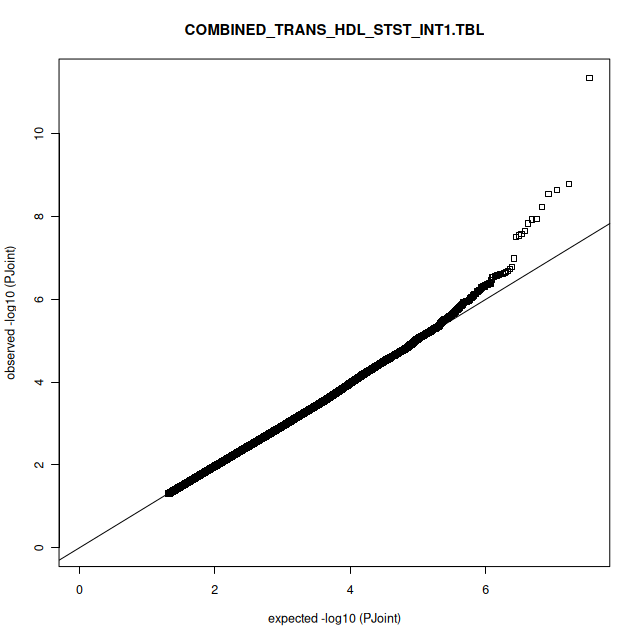 |
| LDL-c and LTST |  |
| 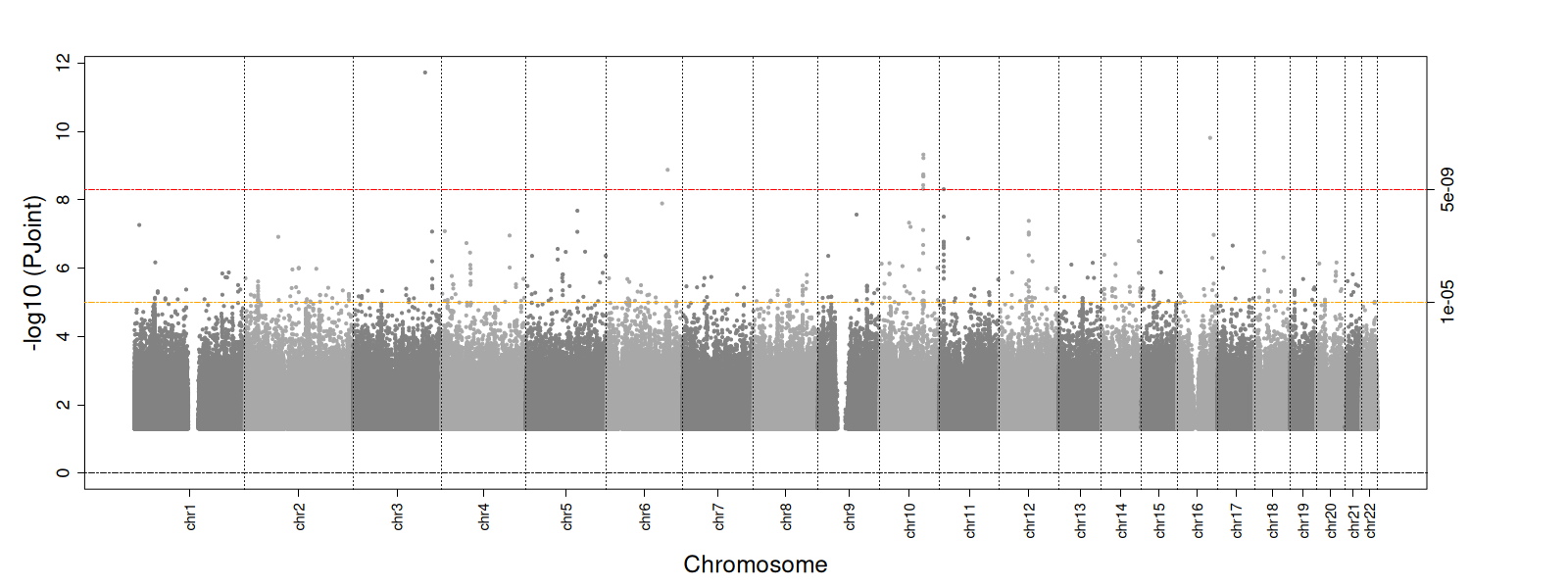 | 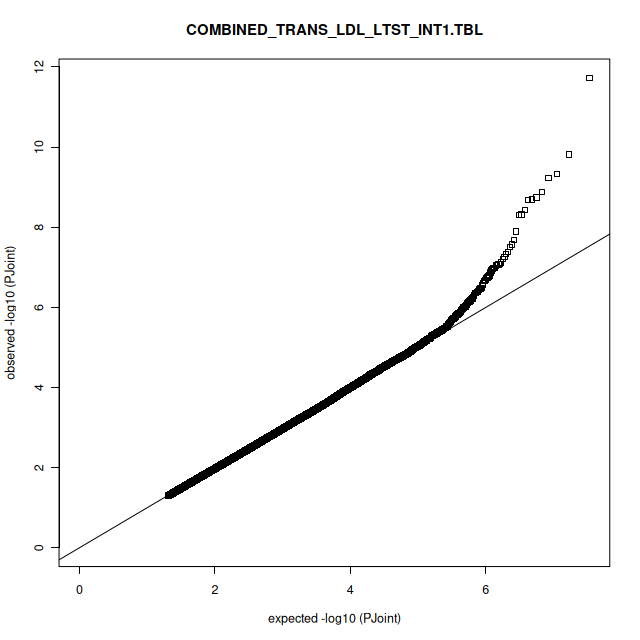 |
| LDL-c and STST |  |
| 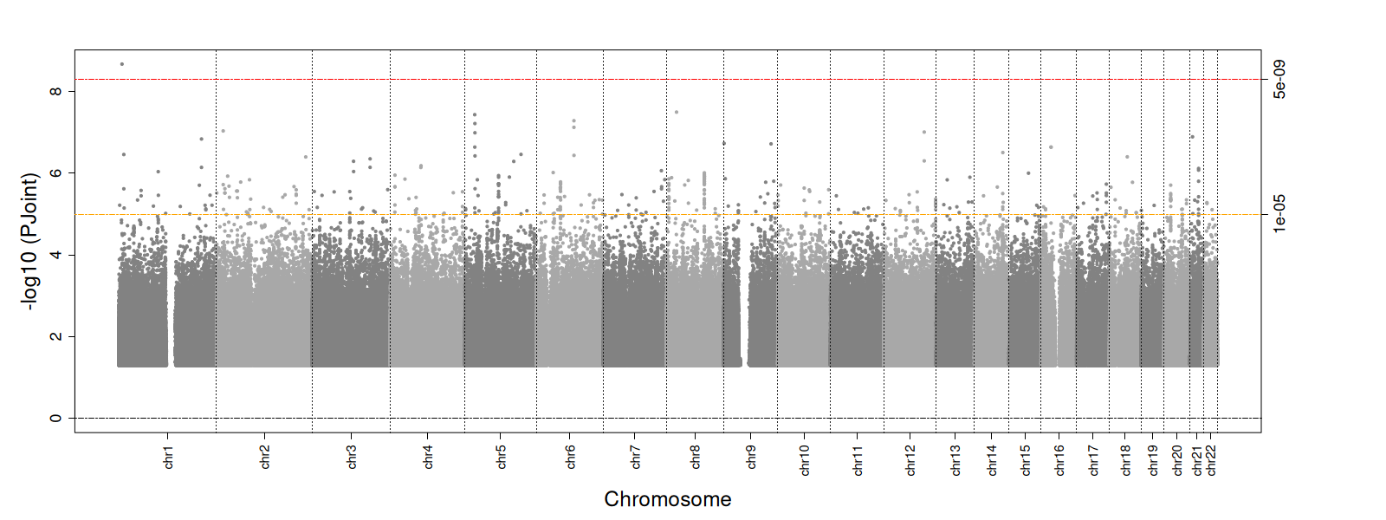 | 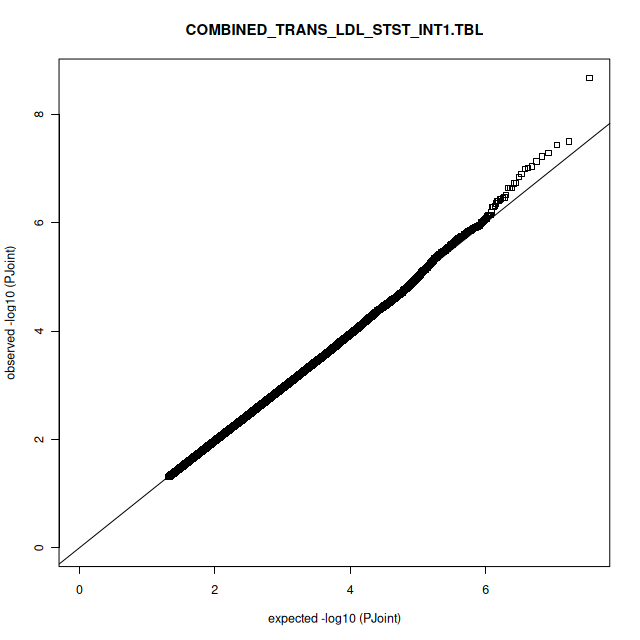 |
| TG and LTST |  |
| 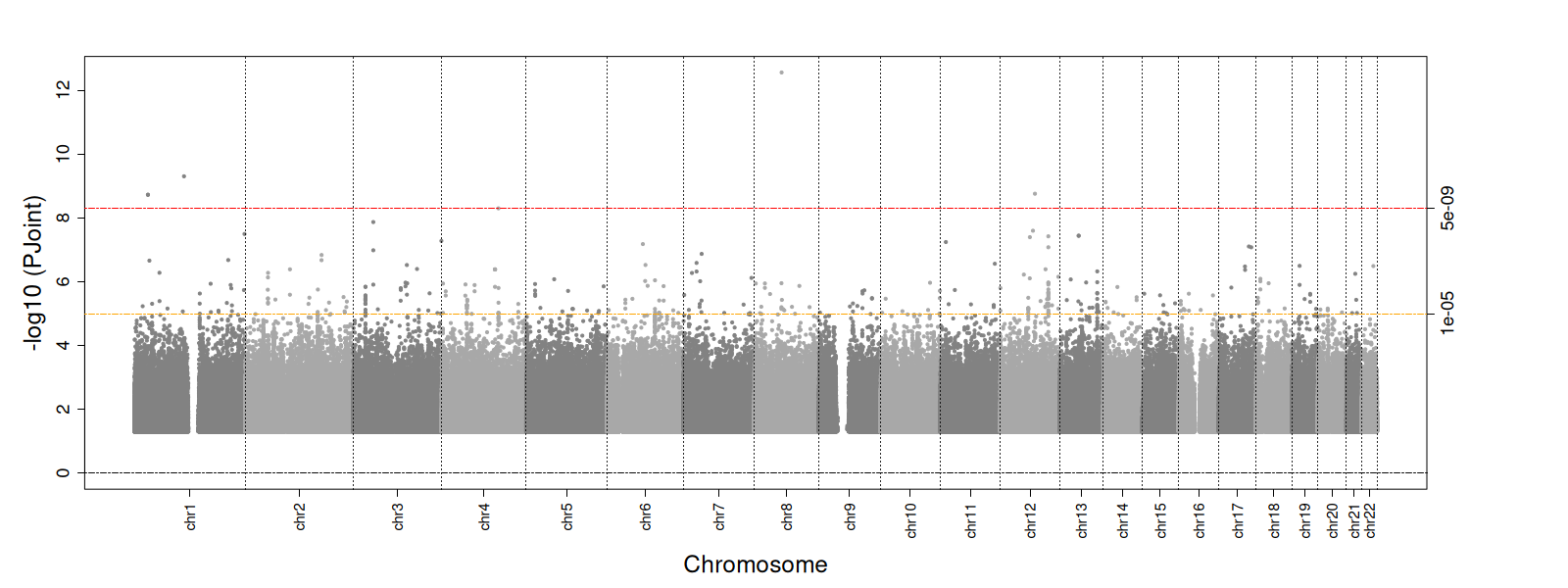 | 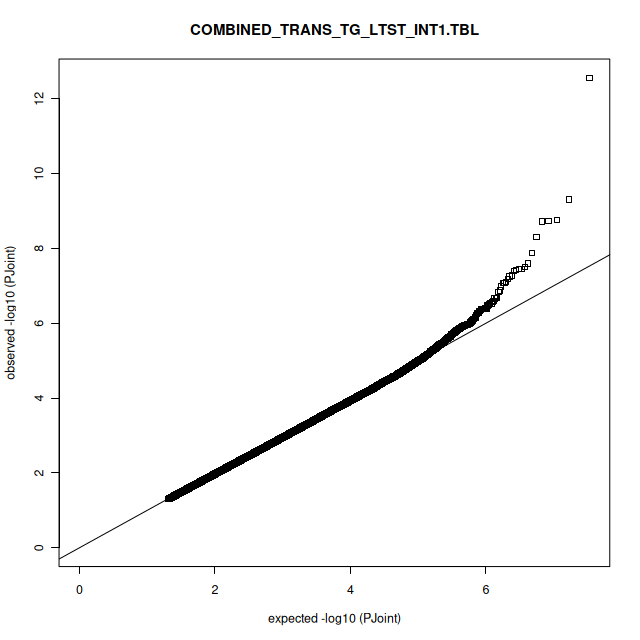 |
| TG and STST |  |
| 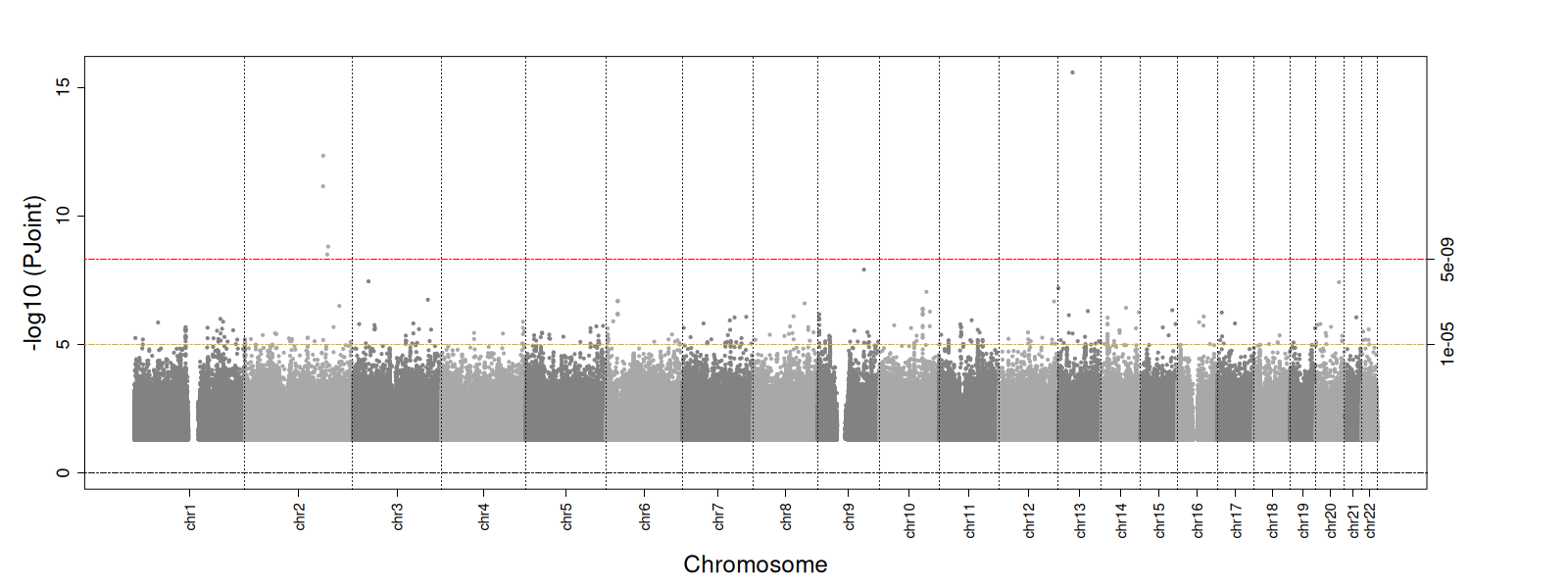 | 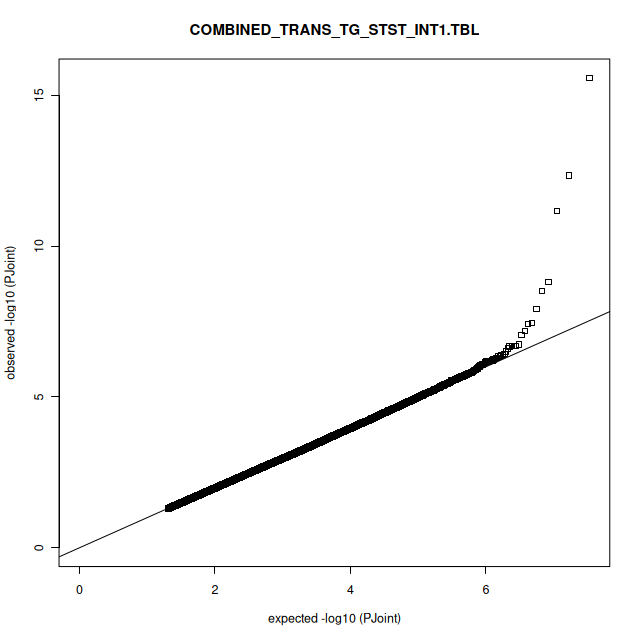 |

**Supplementary Table 2: -log(p) and accompanying Q-Q plots for the European one-df freedom sleep interaction analyses**

| HDL-c and LTST |  |
| --- | --- |
| 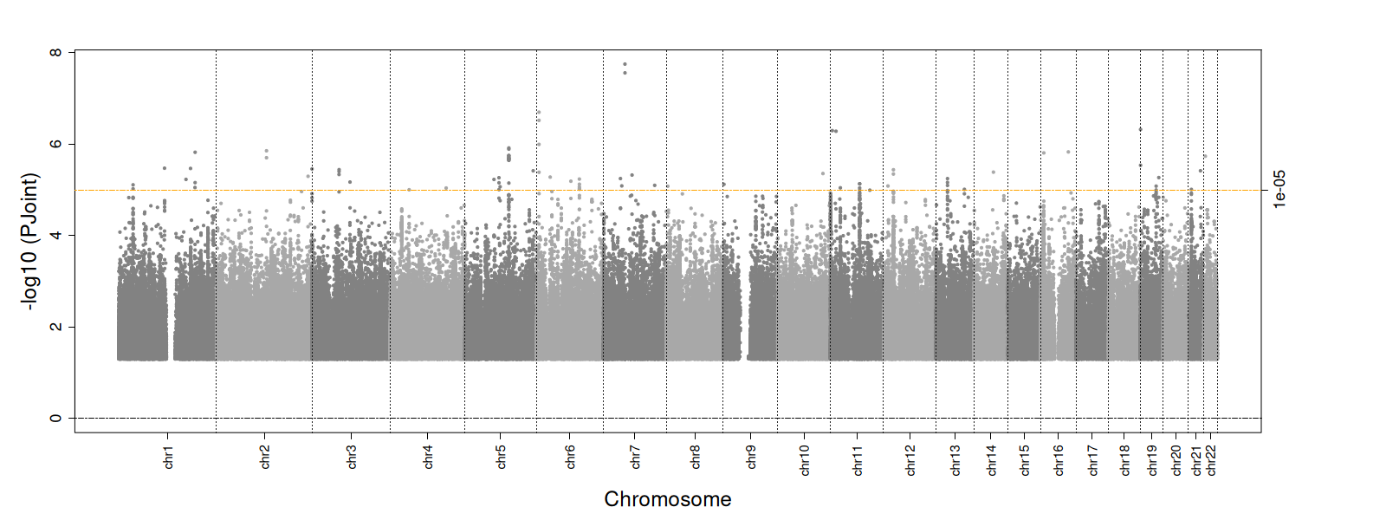 | 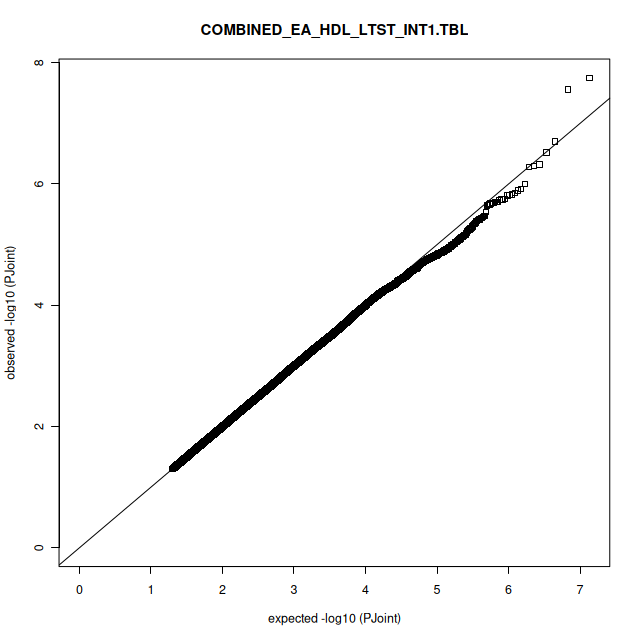 |
| HDL-c and STST |  |
| 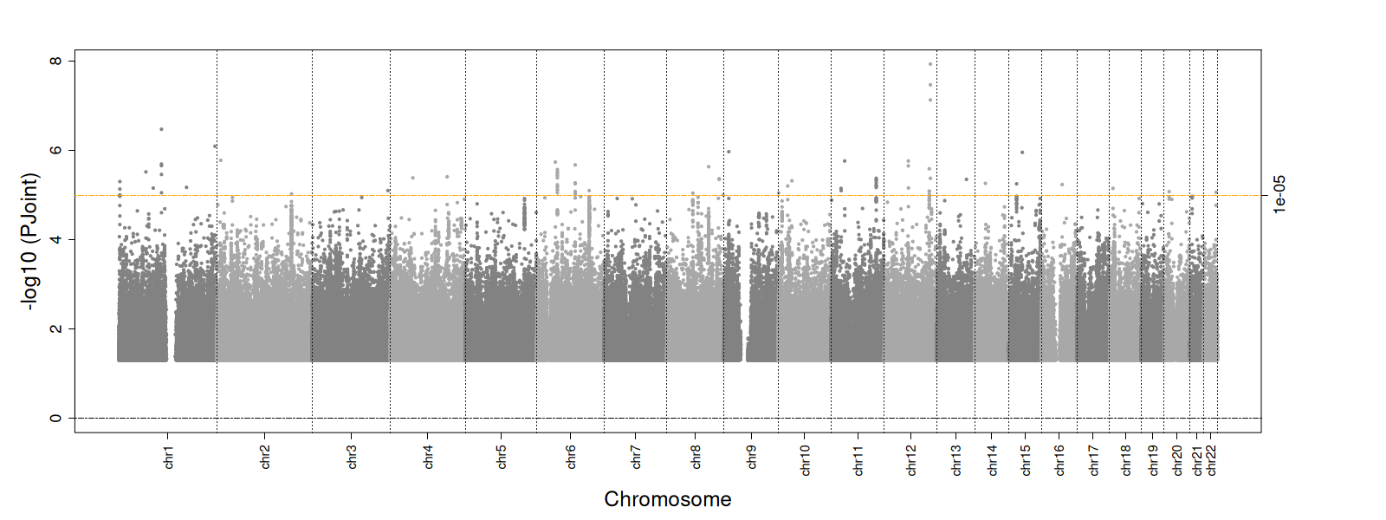 | 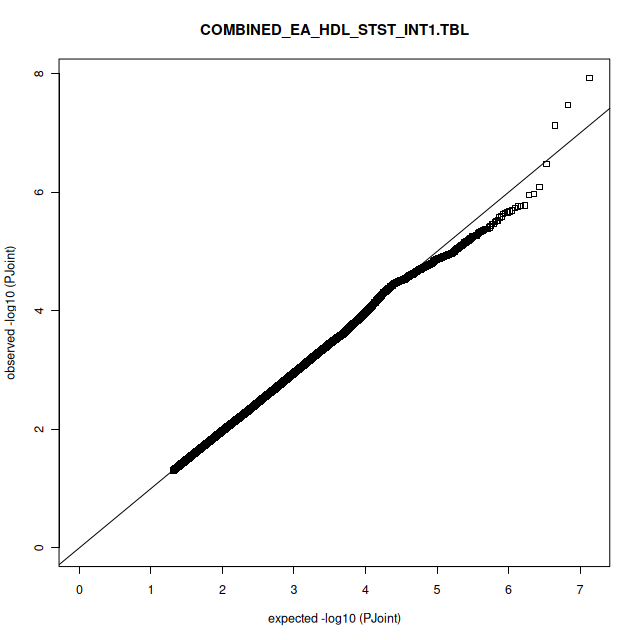 |
| LDL-c and LTST |  |
| 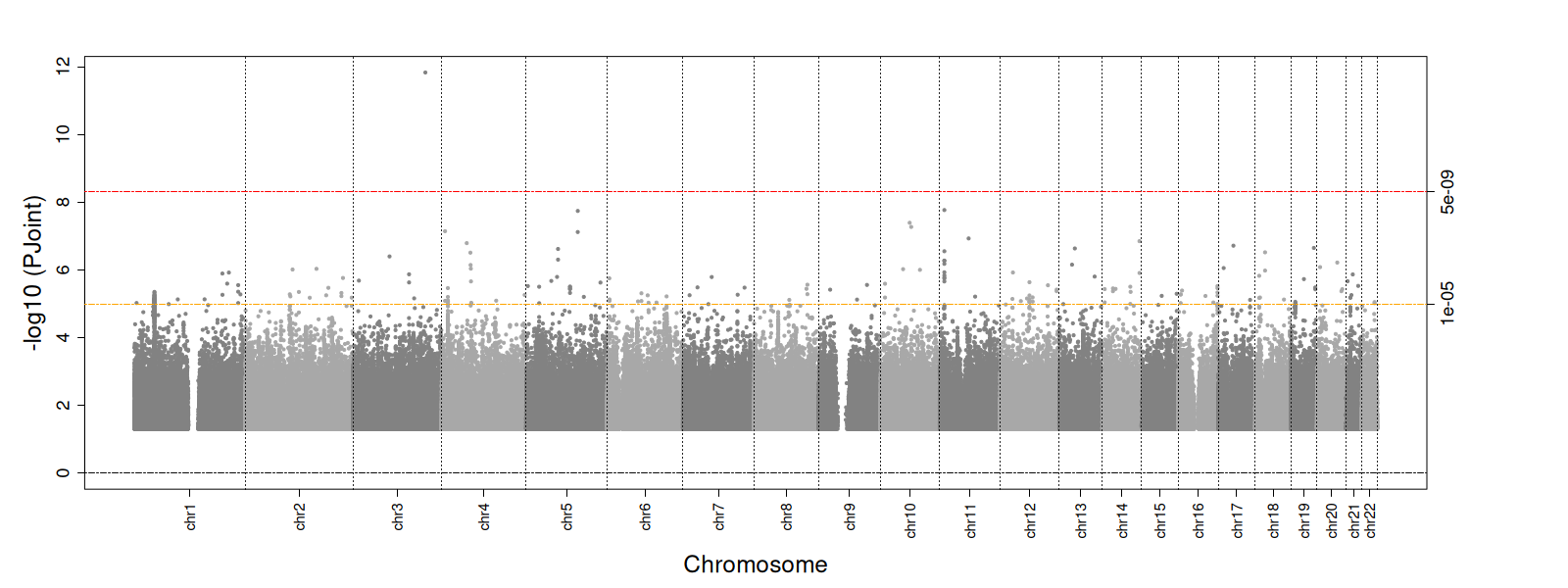 | 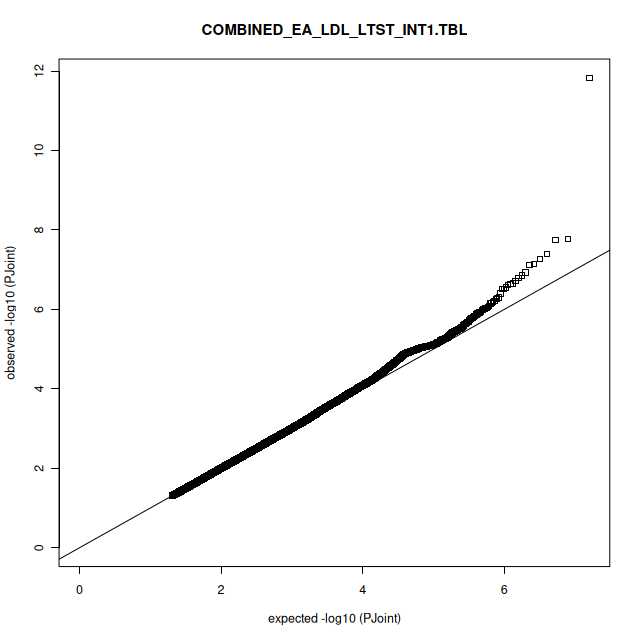 |
| LDL-c and STST |  |
| 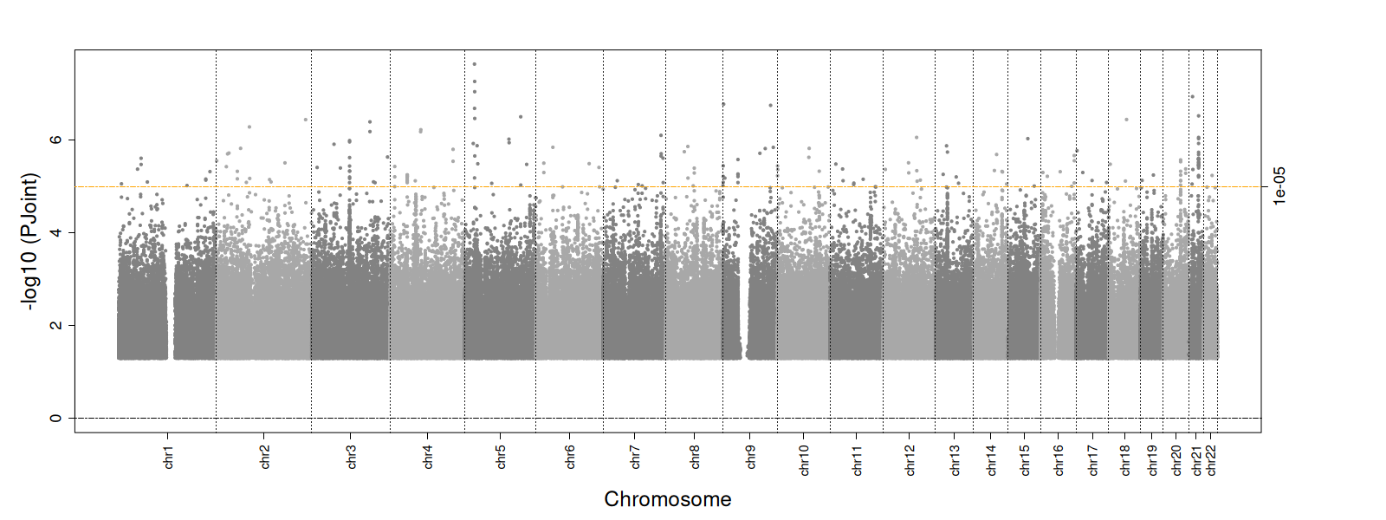 | 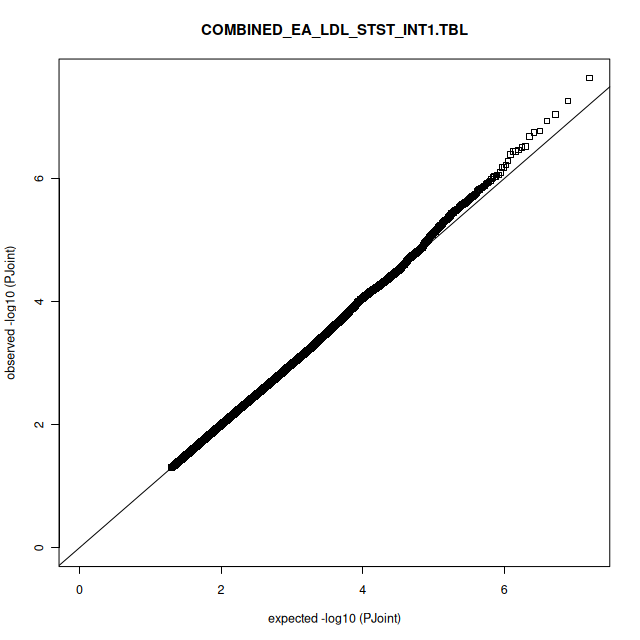 |
| TG and LTST |  |
| 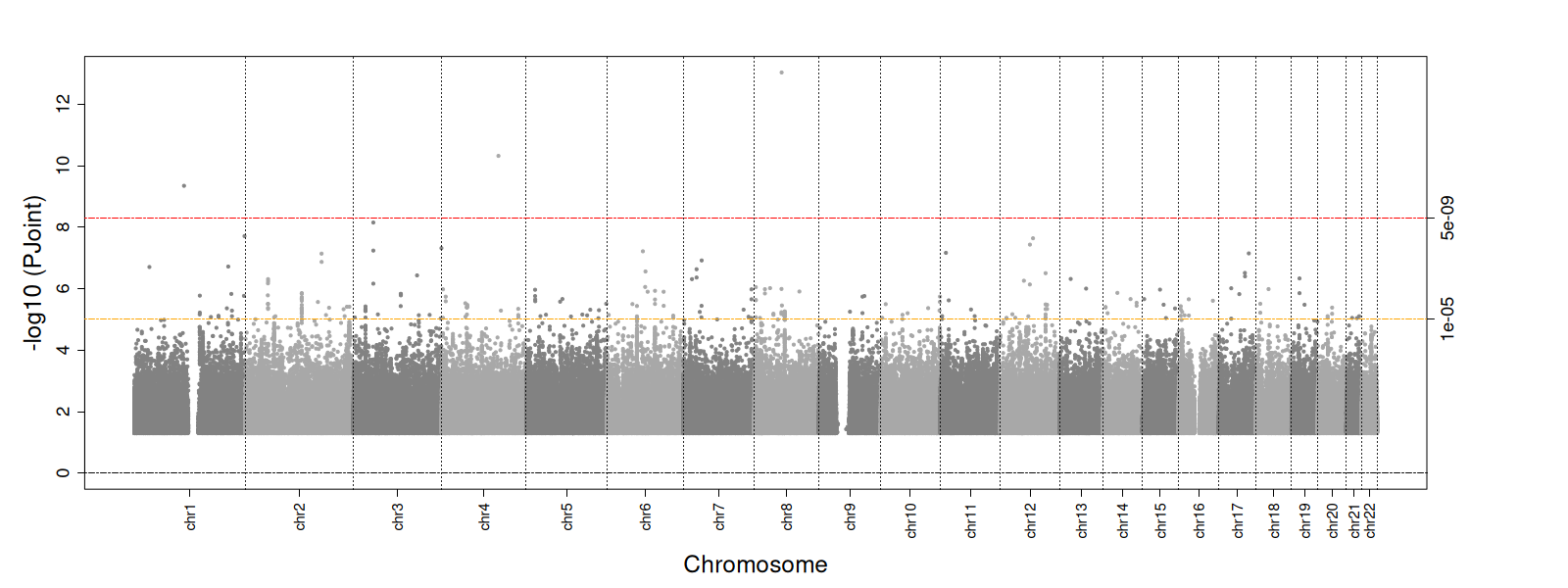 | 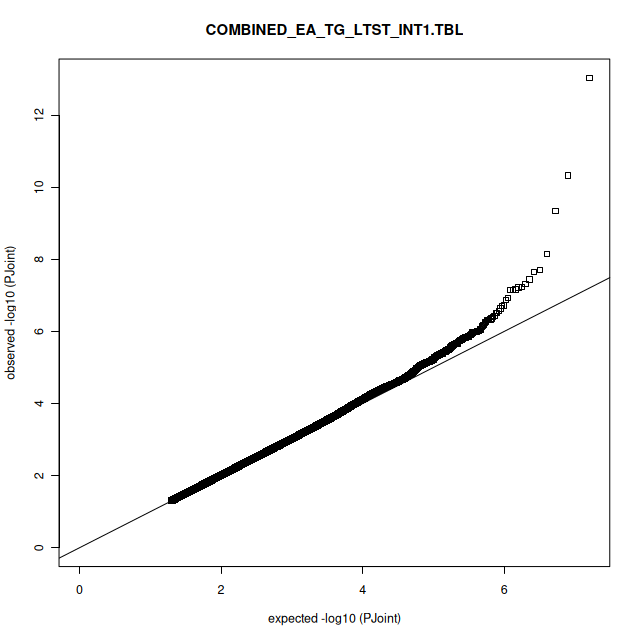 |
| TG and STST |  |
| 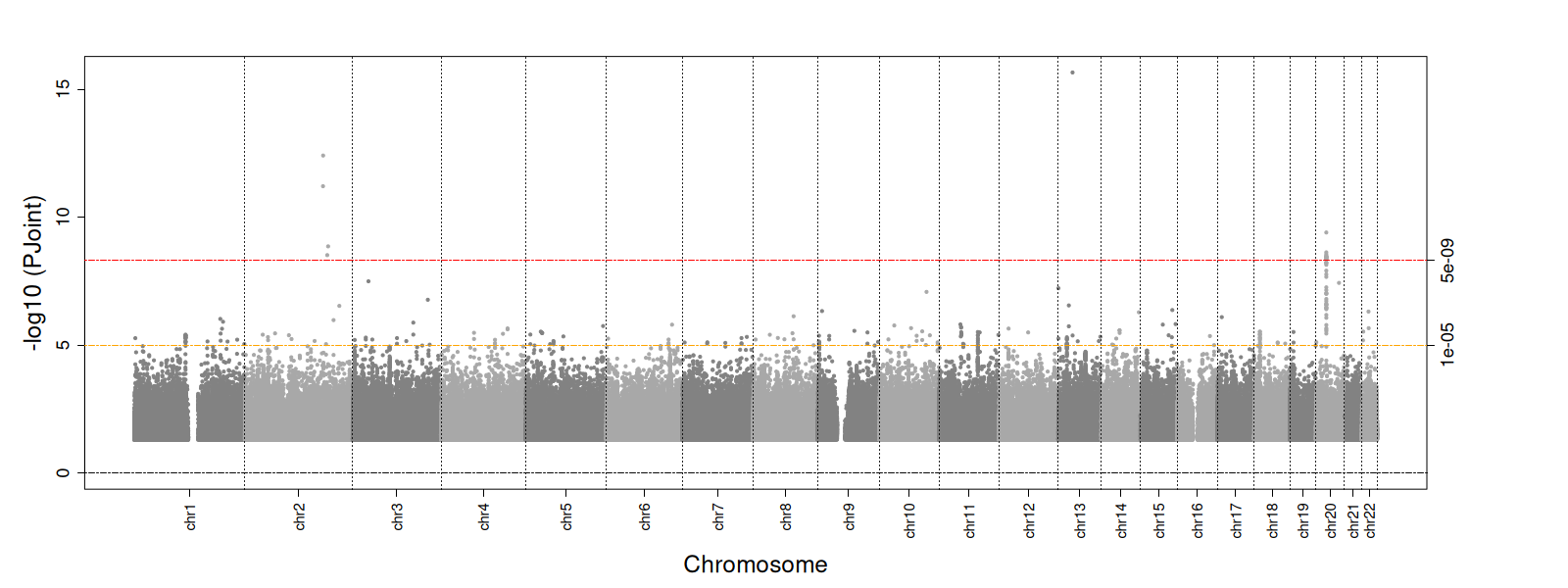 | 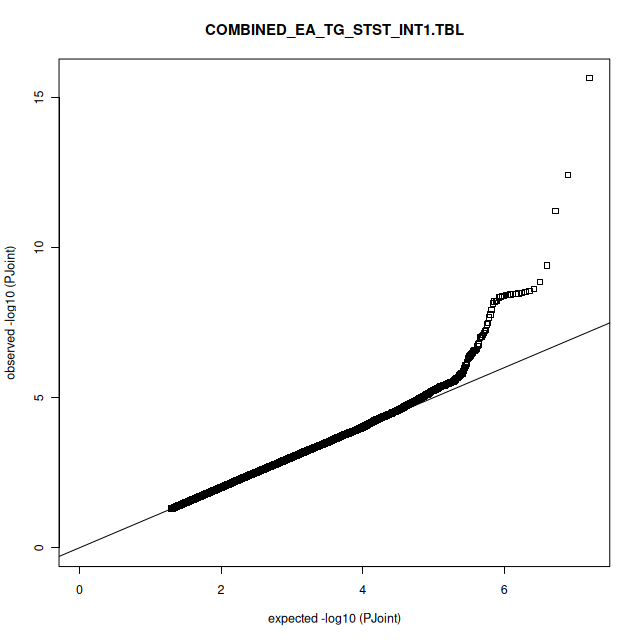 |

**Supplementary Table 3: -log(p) and accompanying Q-Q plots for the African one-df freedom sleep interaction analyses**

| HDL-c and LTST |  |
| --- | --- |
| 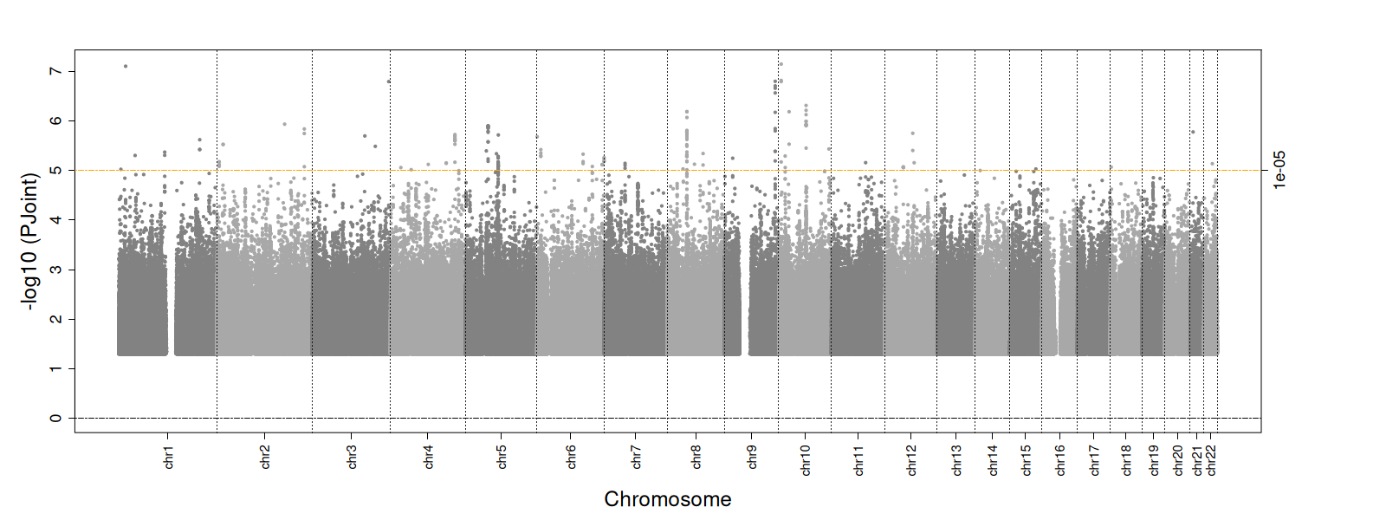 | 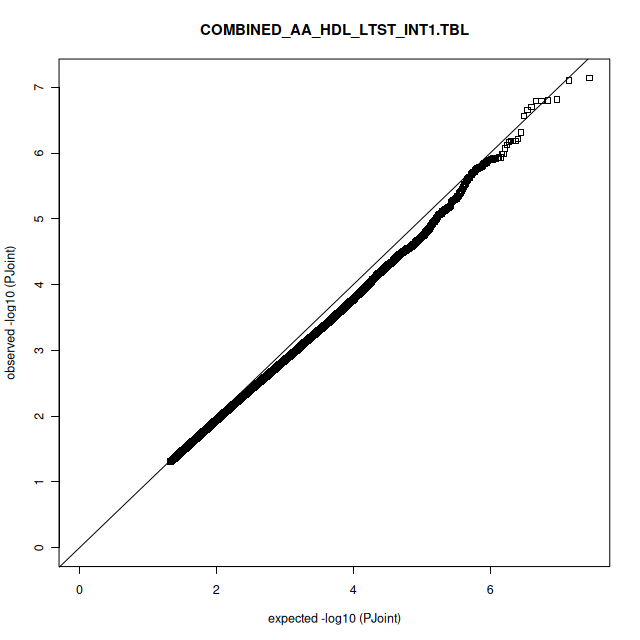 |
| HDL-c and STST |  |
| 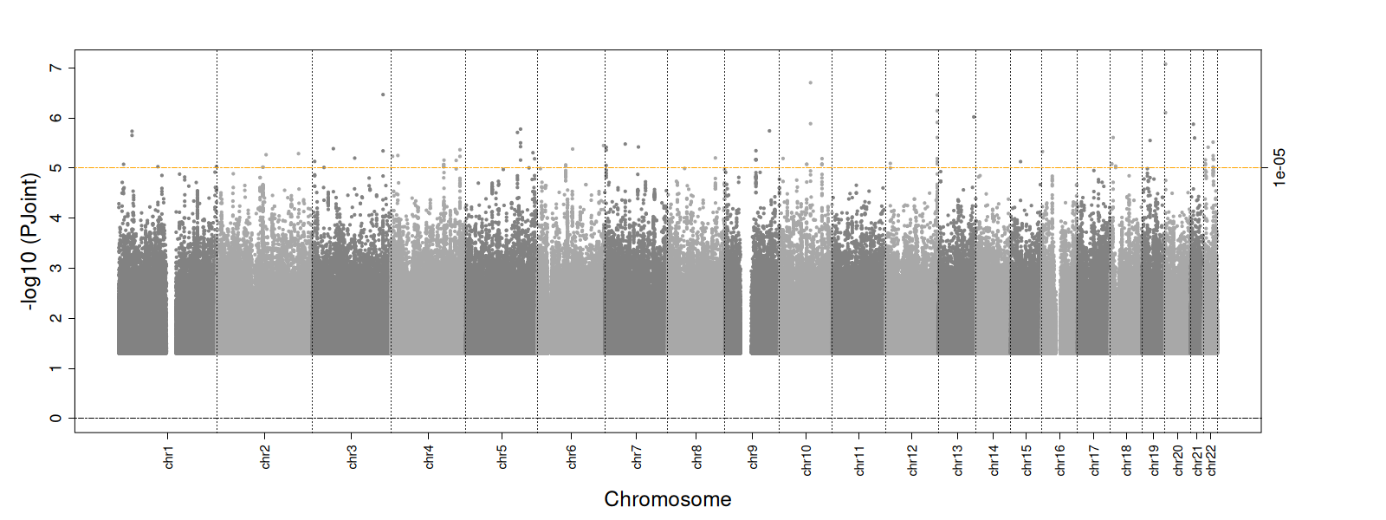 | 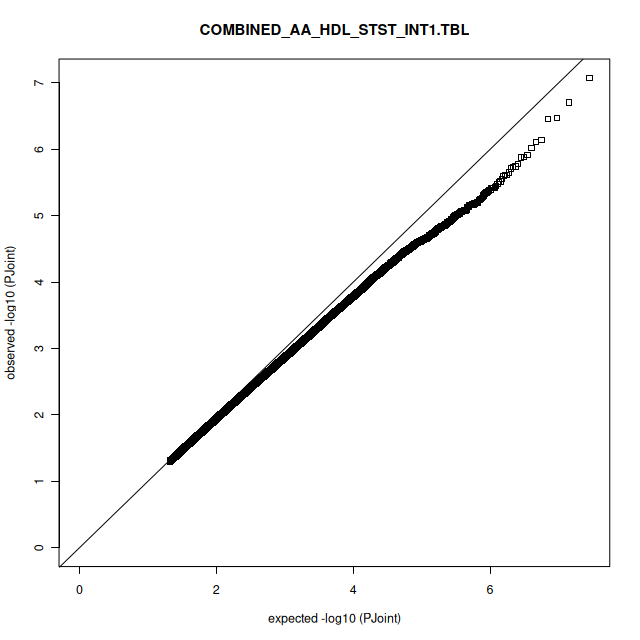 |
| LDL-c and LTST |  |
| 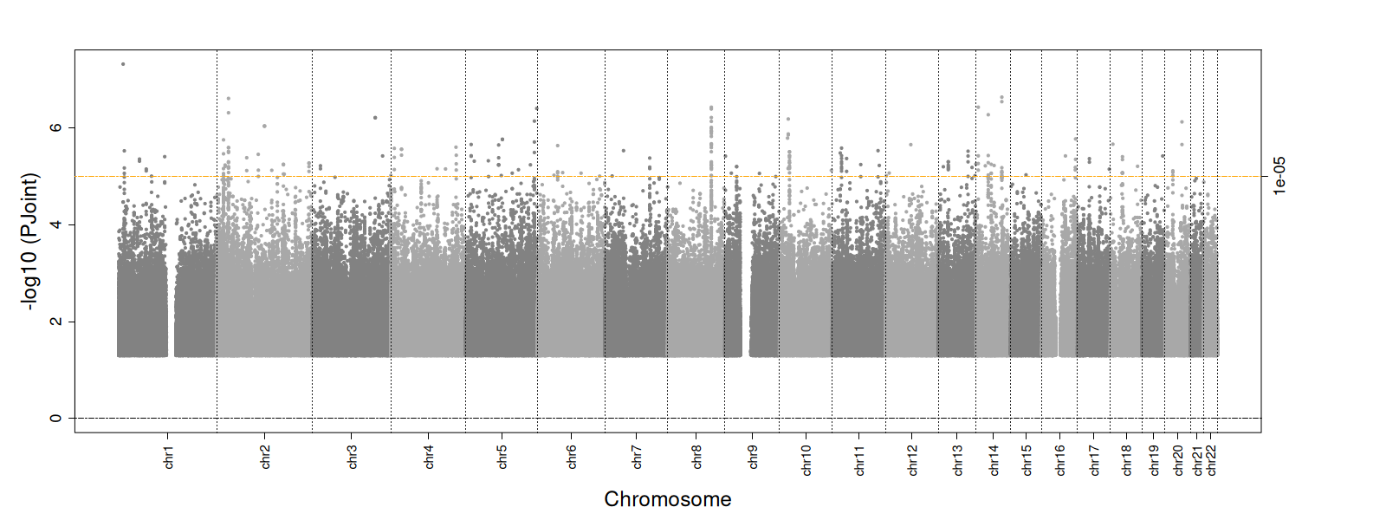 | 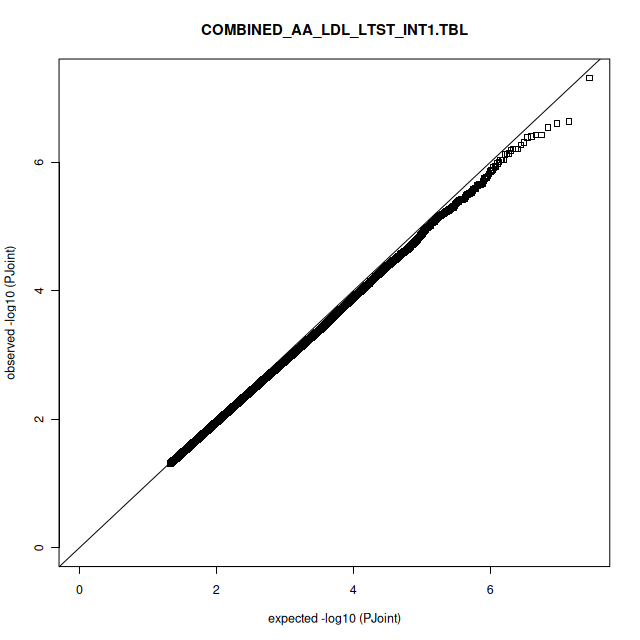 |
| LDL-c and STST |  |
| 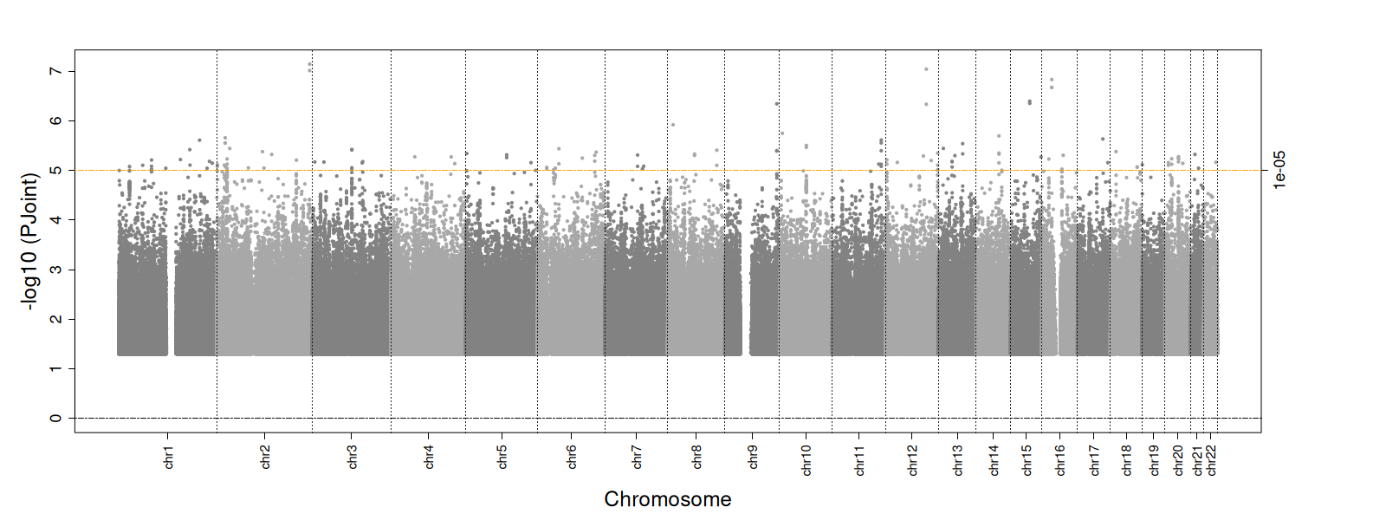 | 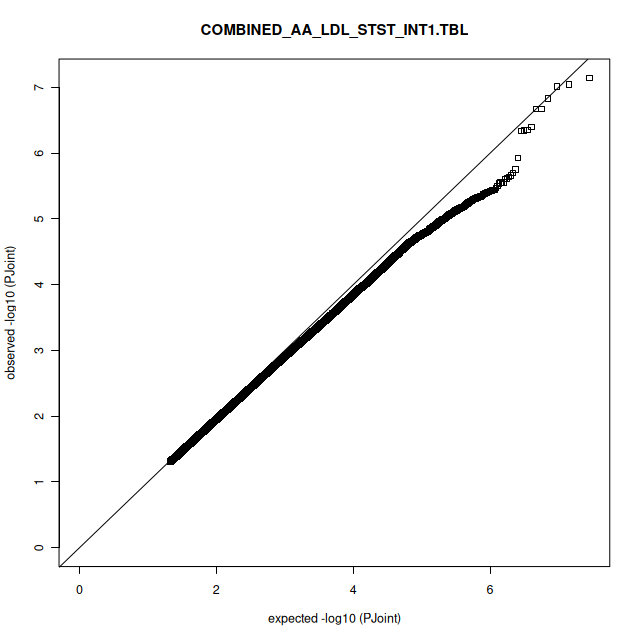 |
| TG and LTST |  |
| 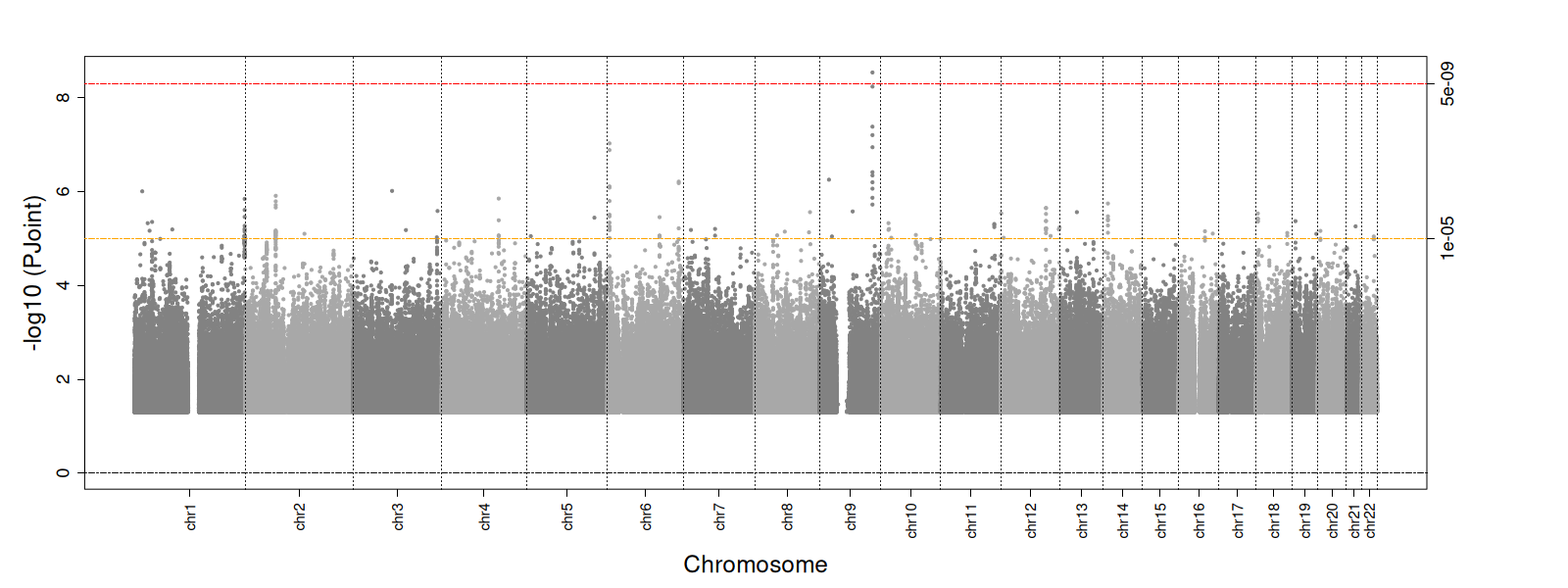 | 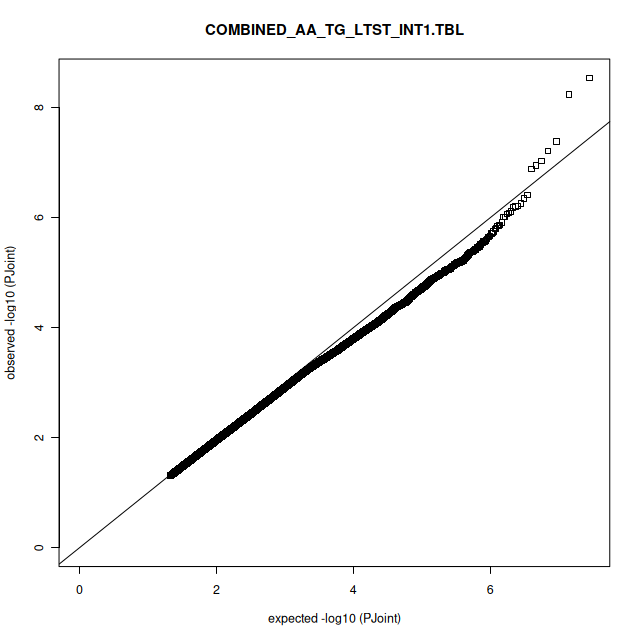 |
| TG and STST |  |
| 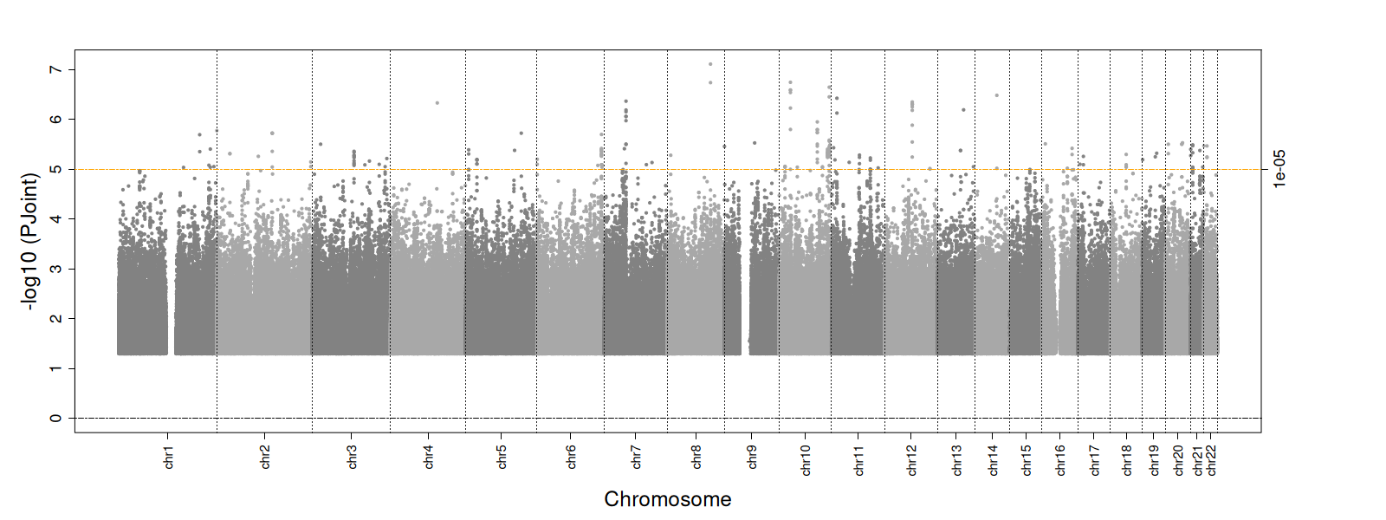 | 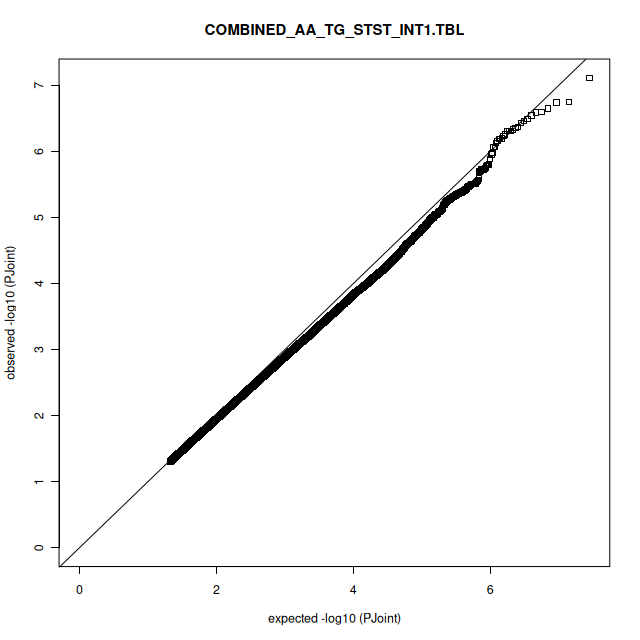 |

**Supplementary Figure 4:** **Regional plots for variants identified through the one-degree of freedom interaction analyses**. Linkage Disequilibrium (LD) information is presented in the figure through colors (blue represents low LD, yellow represents moderate LD, and red high LD). For variants colored in grey, no LD information was available.

A) 13:50374420:C_T

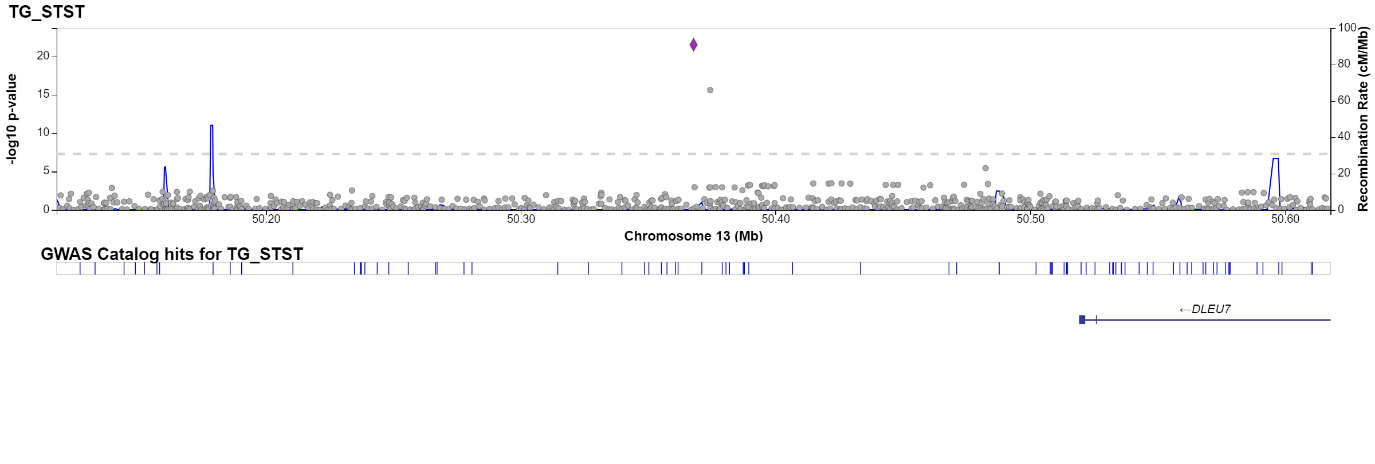

B) 8:61617696:C_T

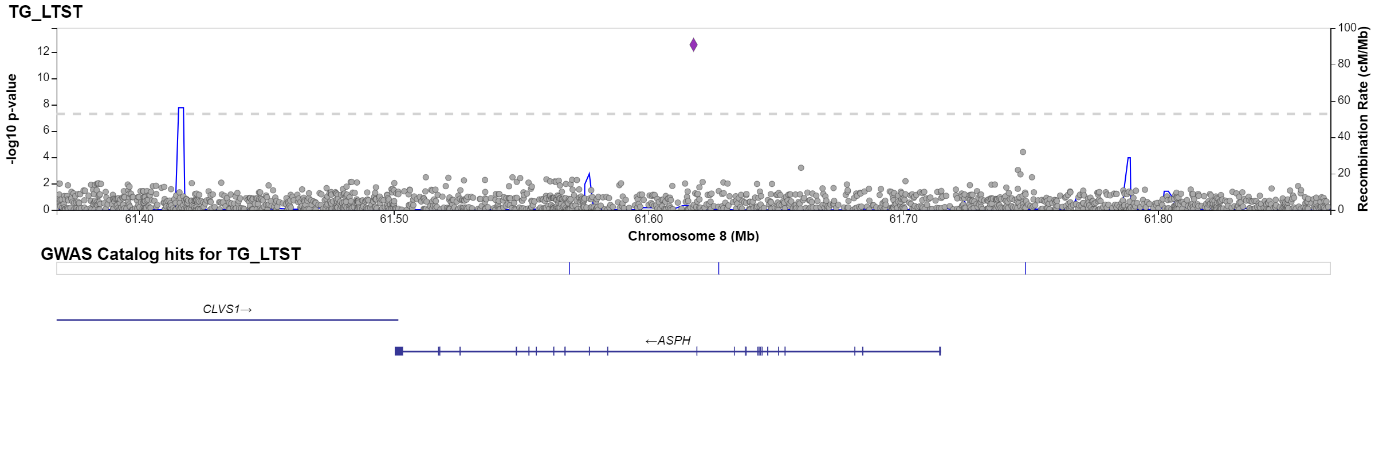

C) 3:162278901:A_T

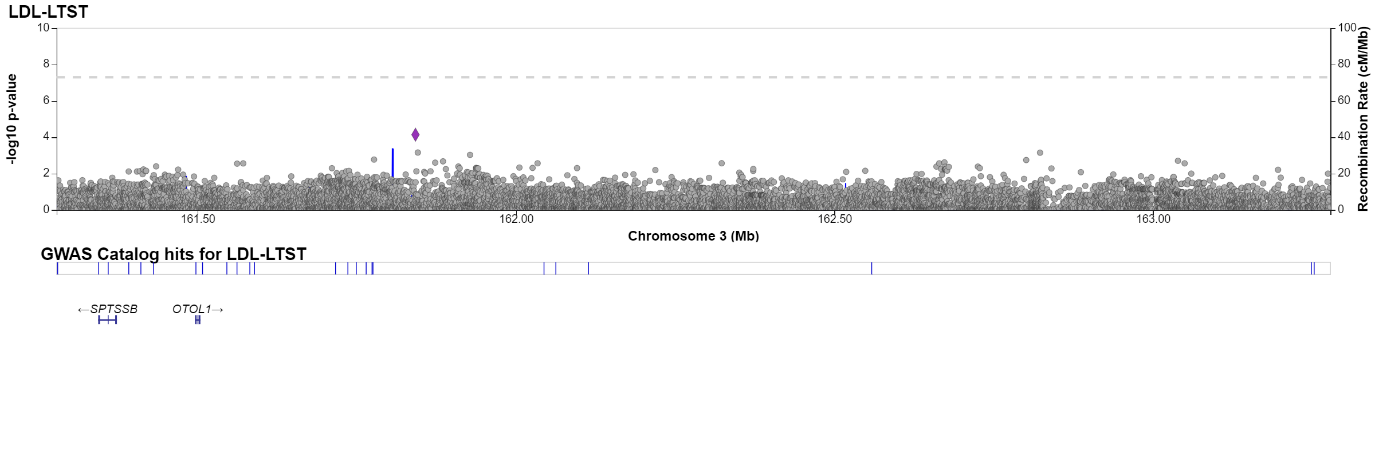

D) 13:50374420:C_T

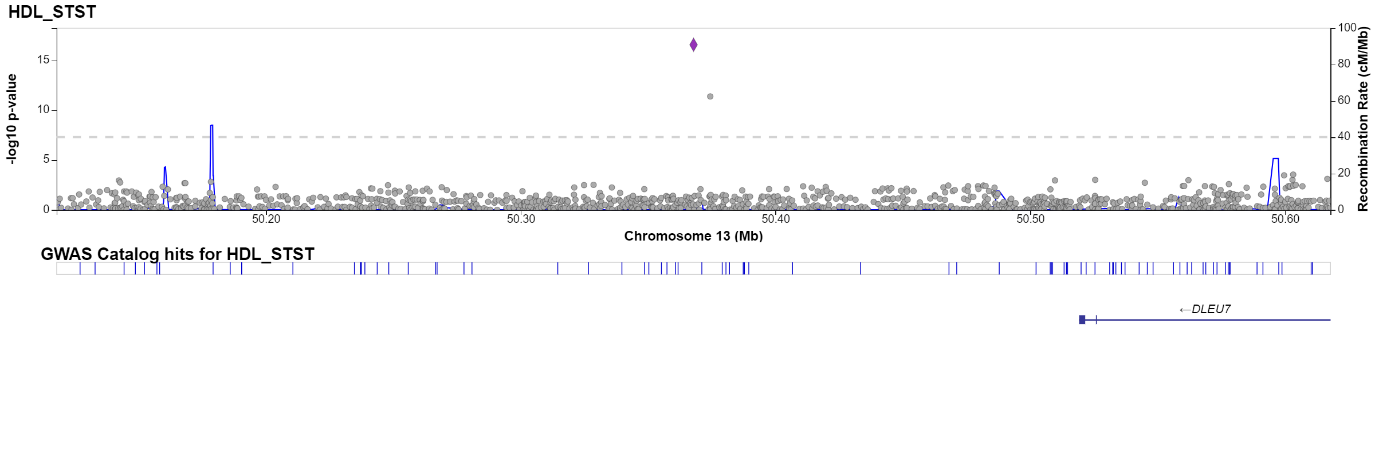

E) 4:127678773:C_G

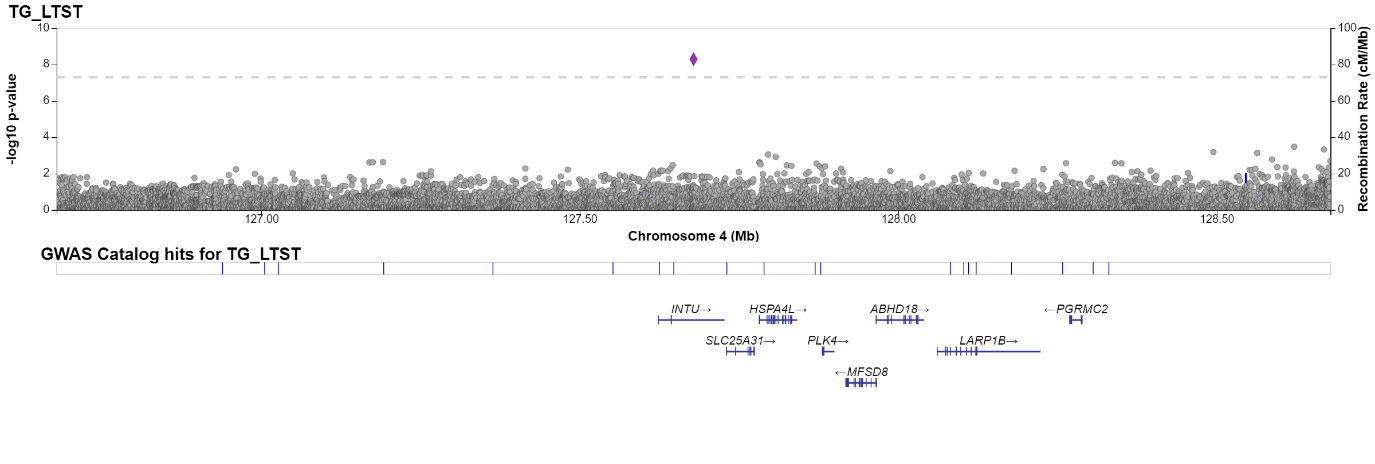

F) 2:186808058:G_T

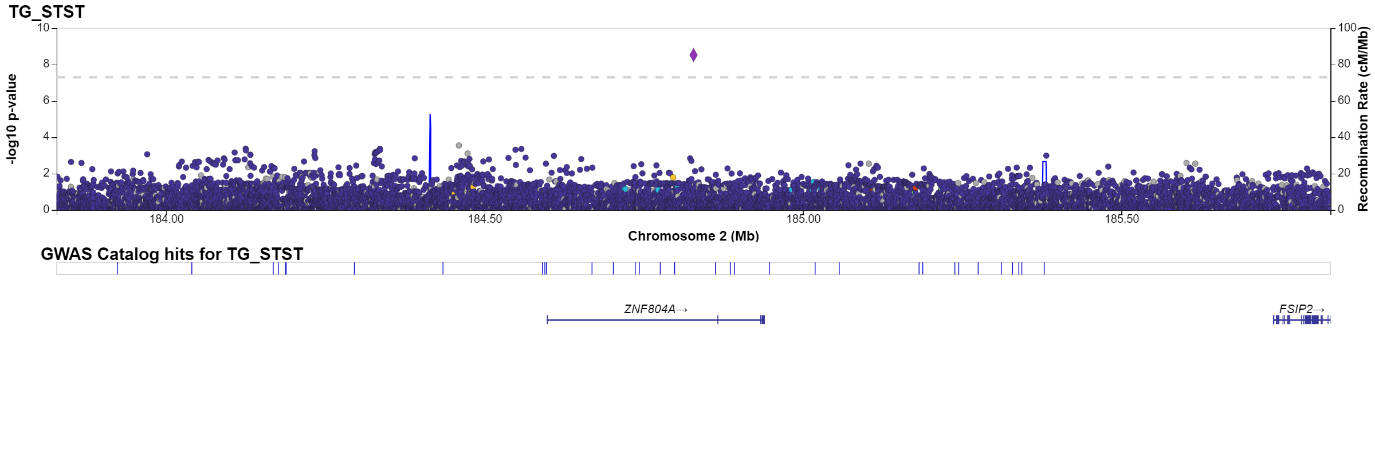

G) 7:102460277:G_T

No plot generated

H) 2:184828292:C_T

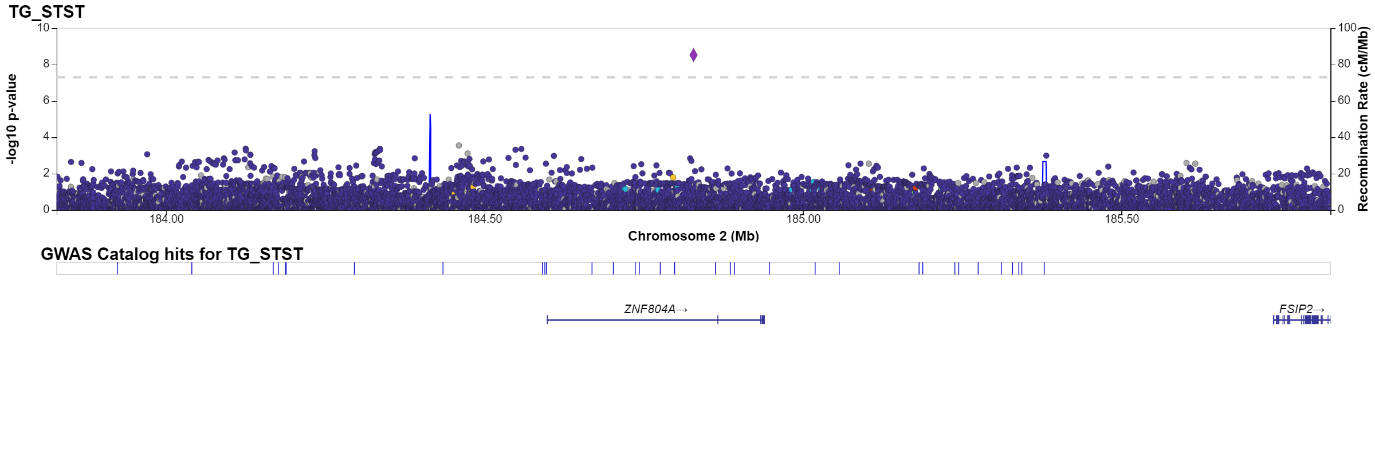

I) 11:10411707:C_CT

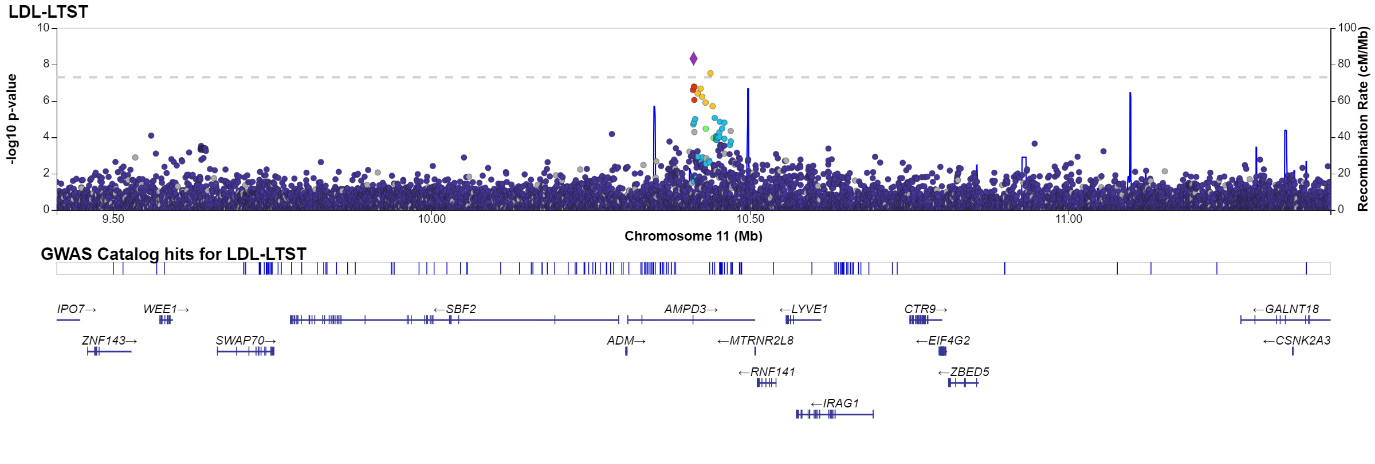

**Supplementary Figure 5: -log(p) and accompanying Q-Q plots for the combined two-df freedom sleep interaction analyses (CPMA)**

| HDL-c and LTST |  |
| --- | --- |
| 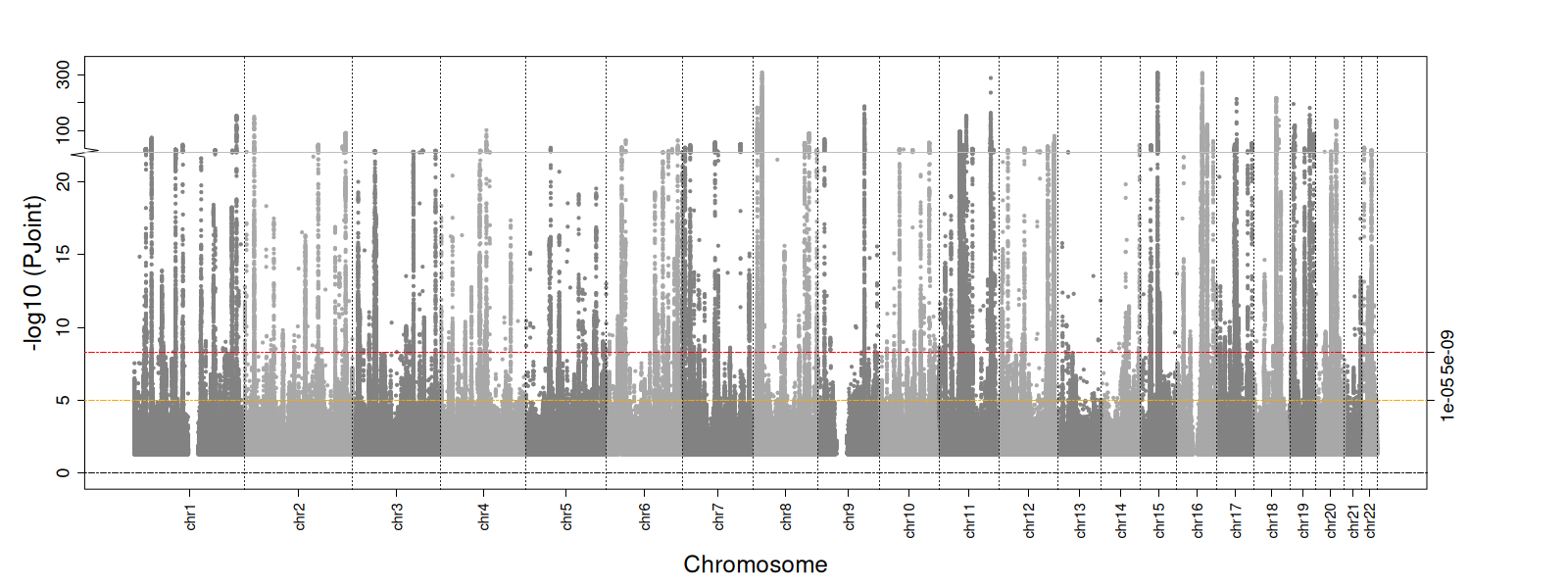 | 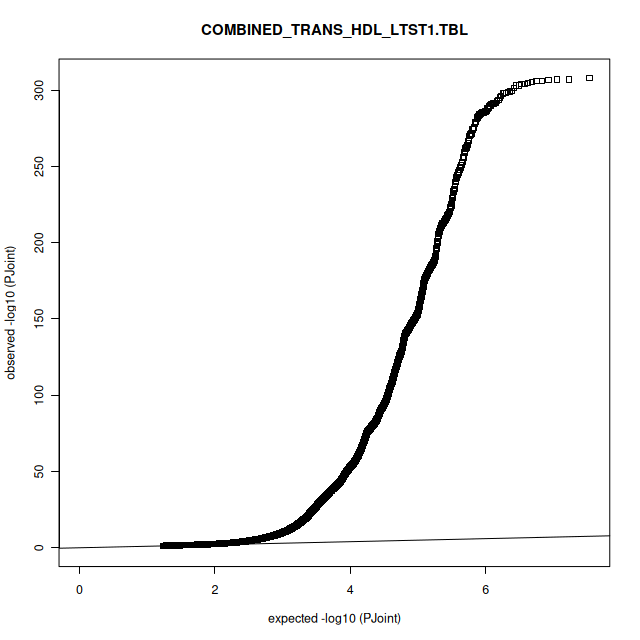 |
| HDL-c and STST |  |
| 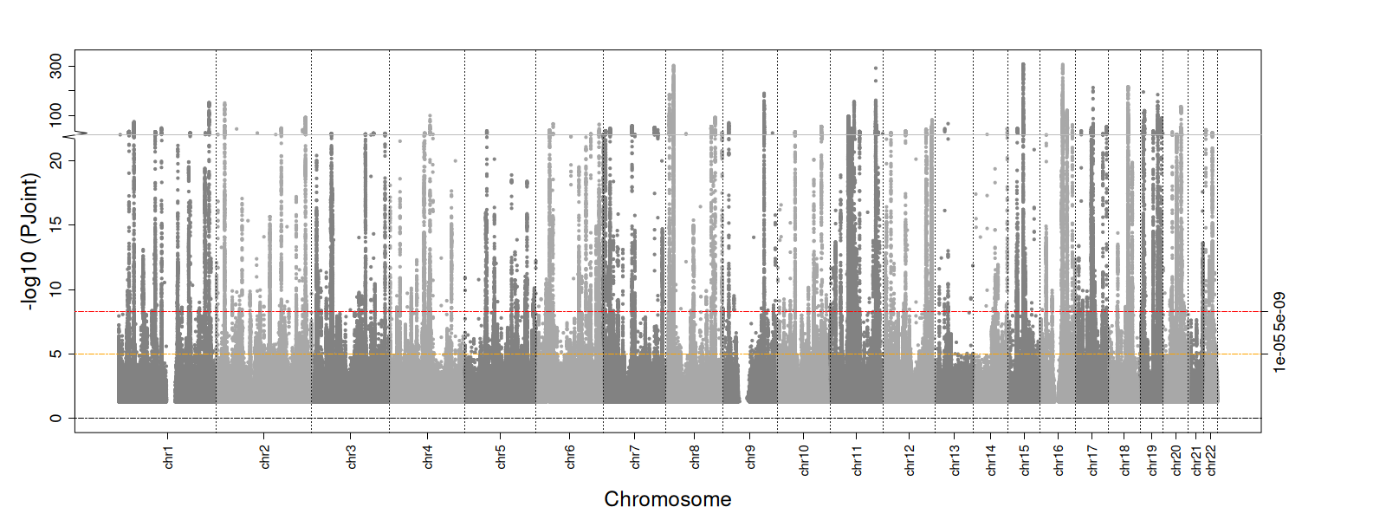 | 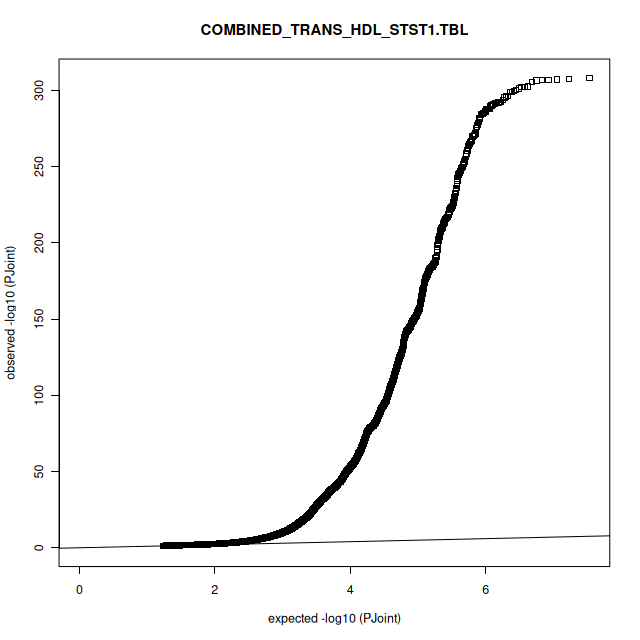 |
| LDL-c and LTST |  |
| 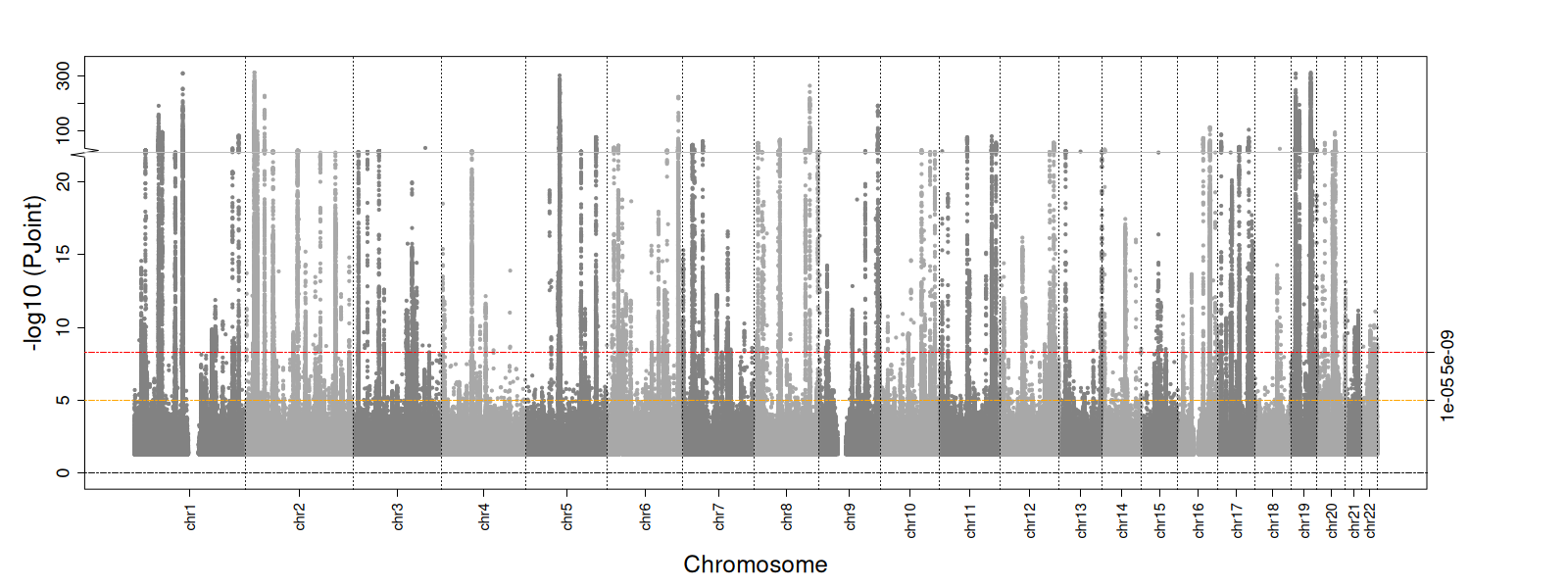 | 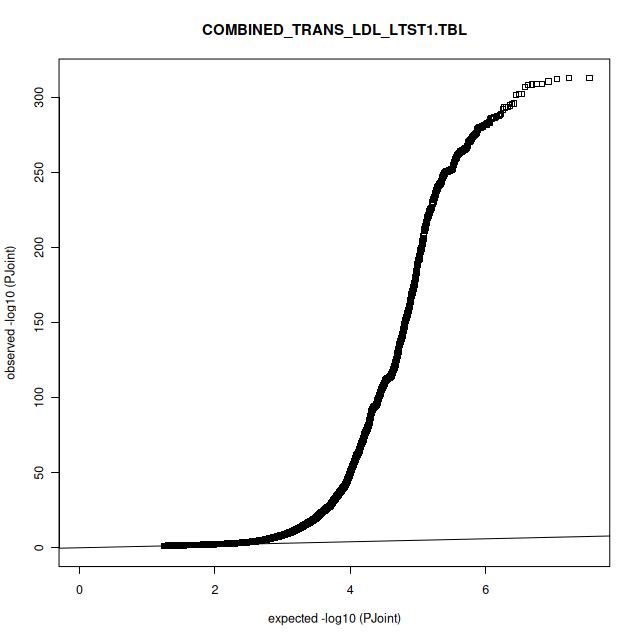 |
| LDL-c and STST |  |
| 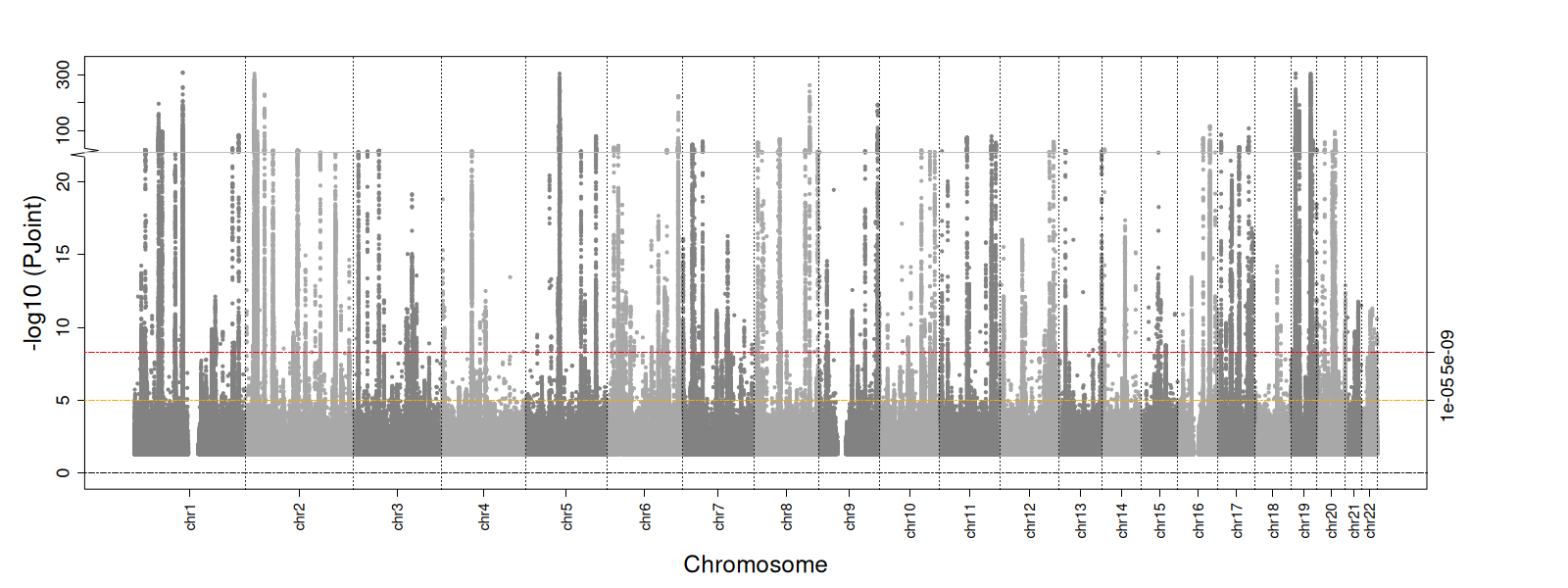 | 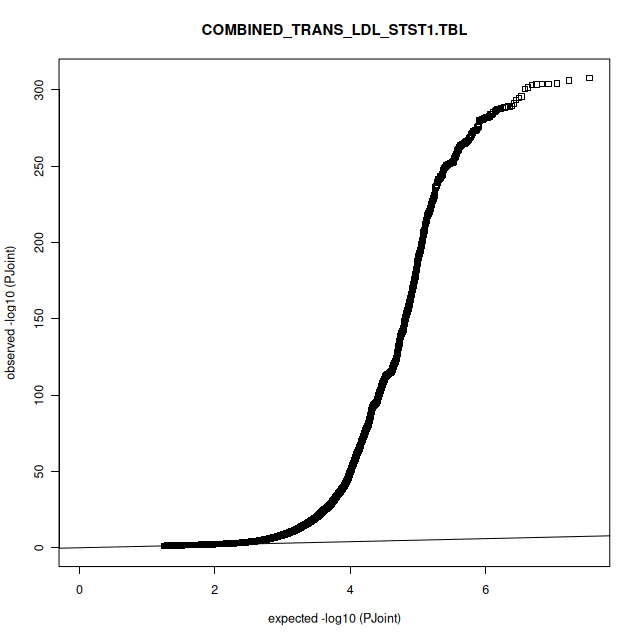 |
| TG and LTST |  |
| 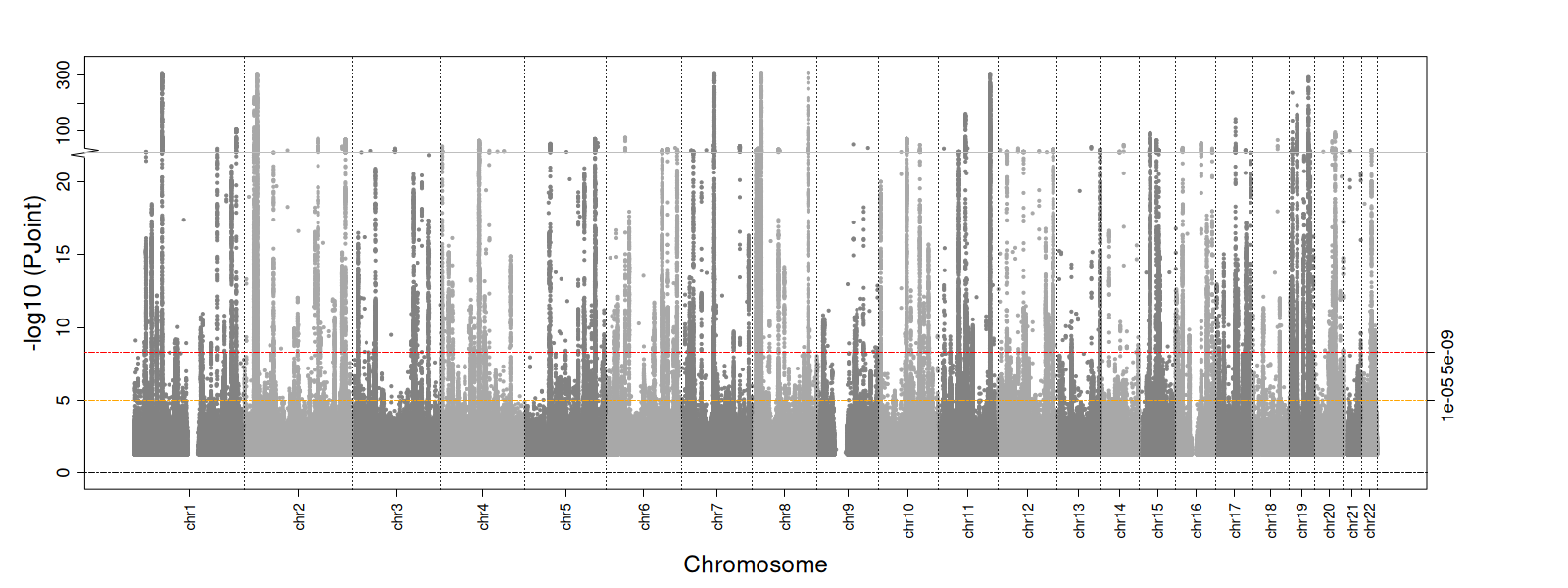 | 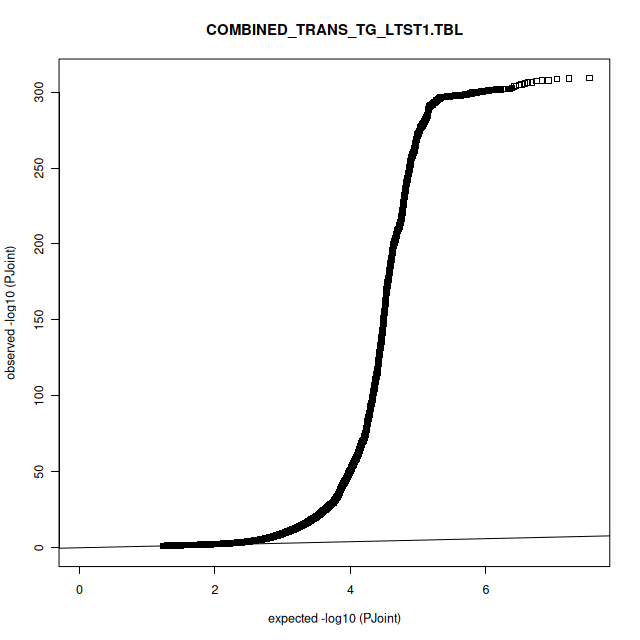 |
| TG and STST |  |
| 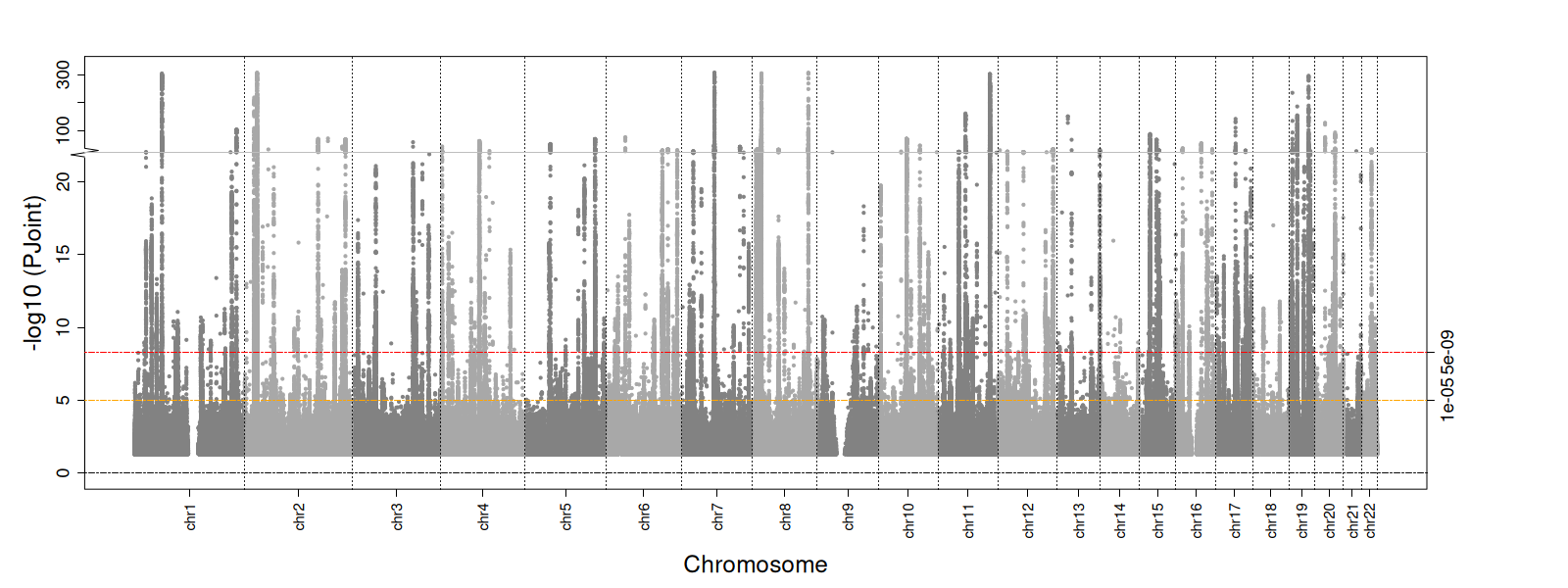 | 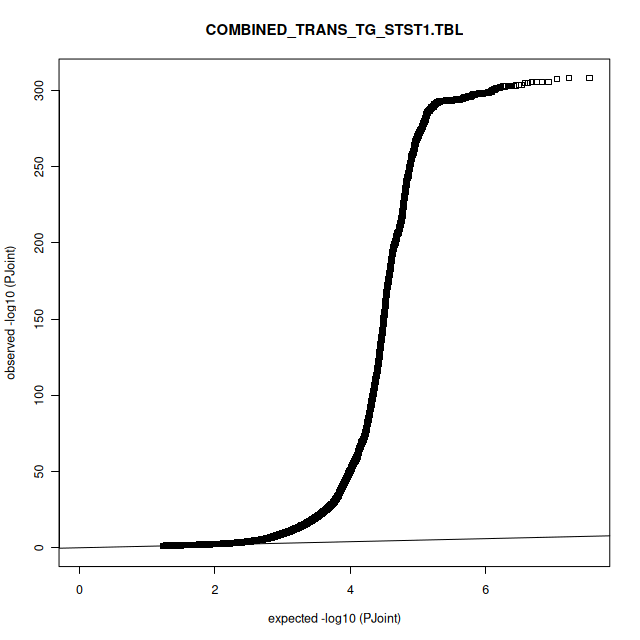 |

**Supplementary Table 6: -log(p) and accompanying Q-Q plots for the European two-df freedom sleep interaction analyses**

| HDL-c and LTST |  |
| --- | --- |
| 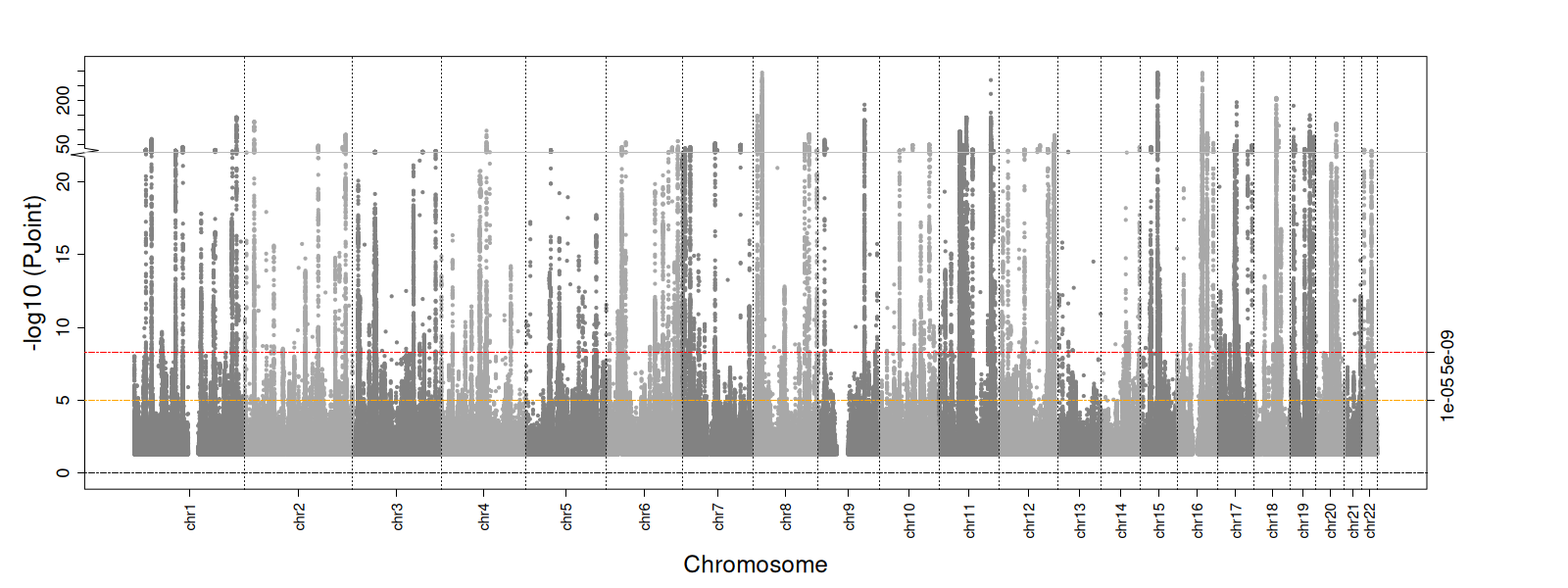 | 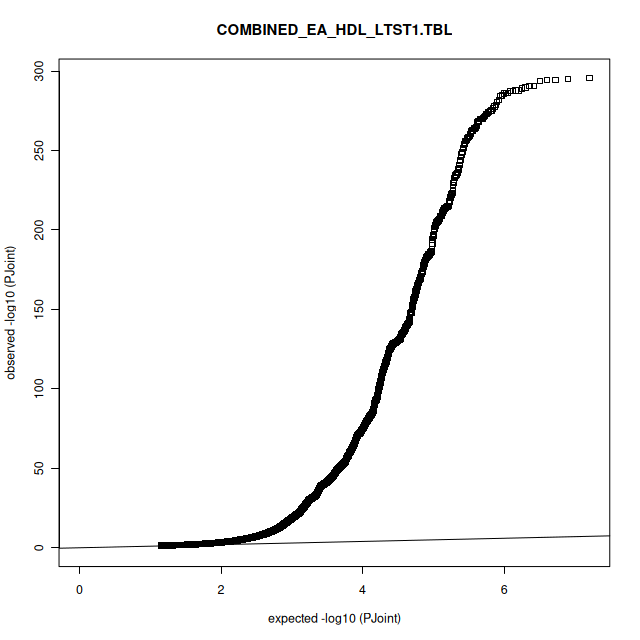 |
| HDL-c and STST |  |
| 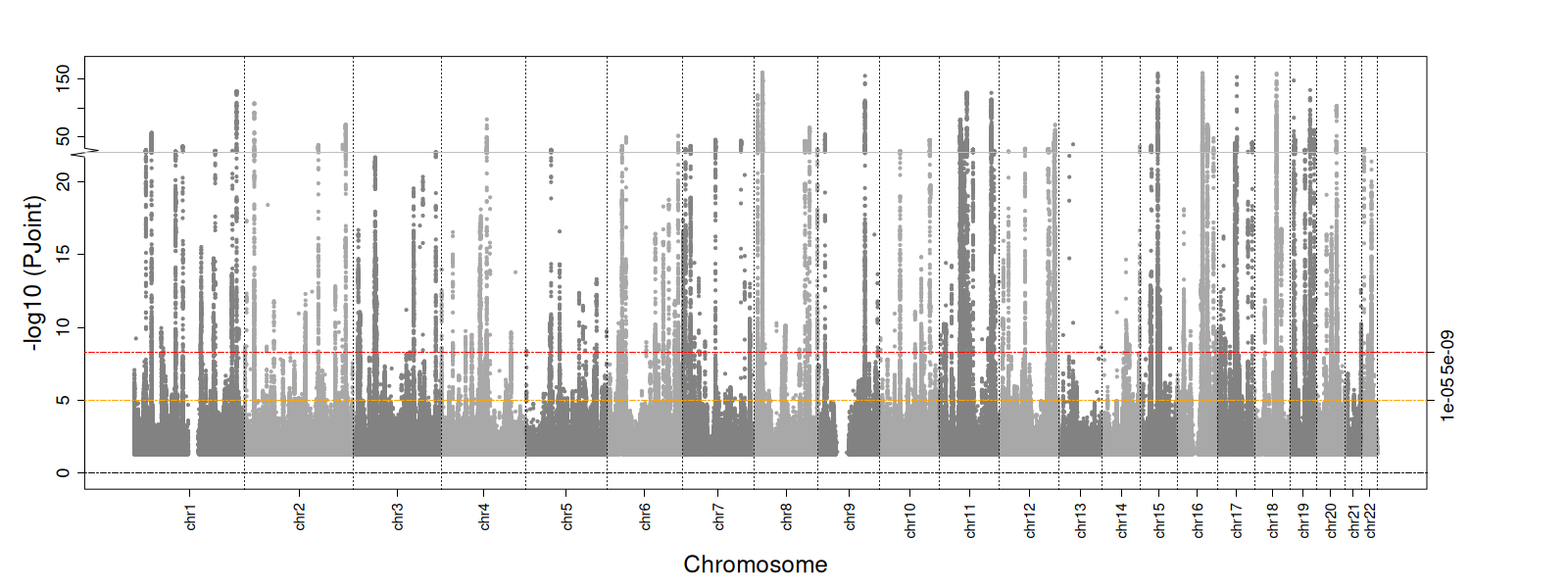 | 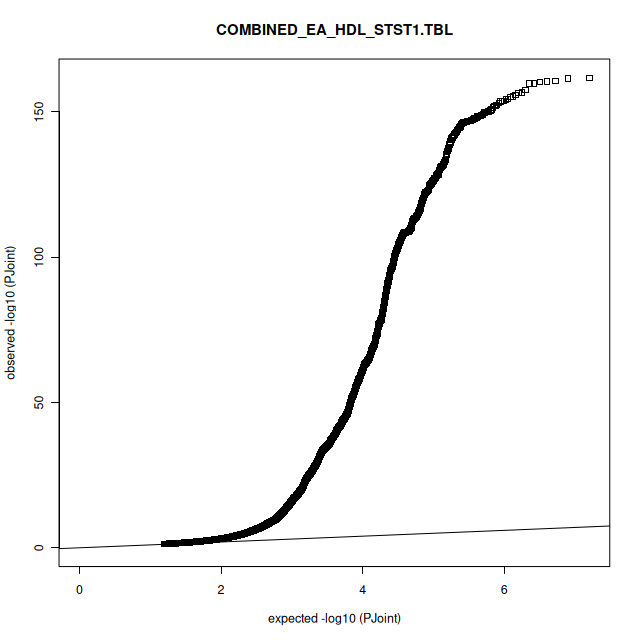 |
| LDL-c and LTST |  |
| 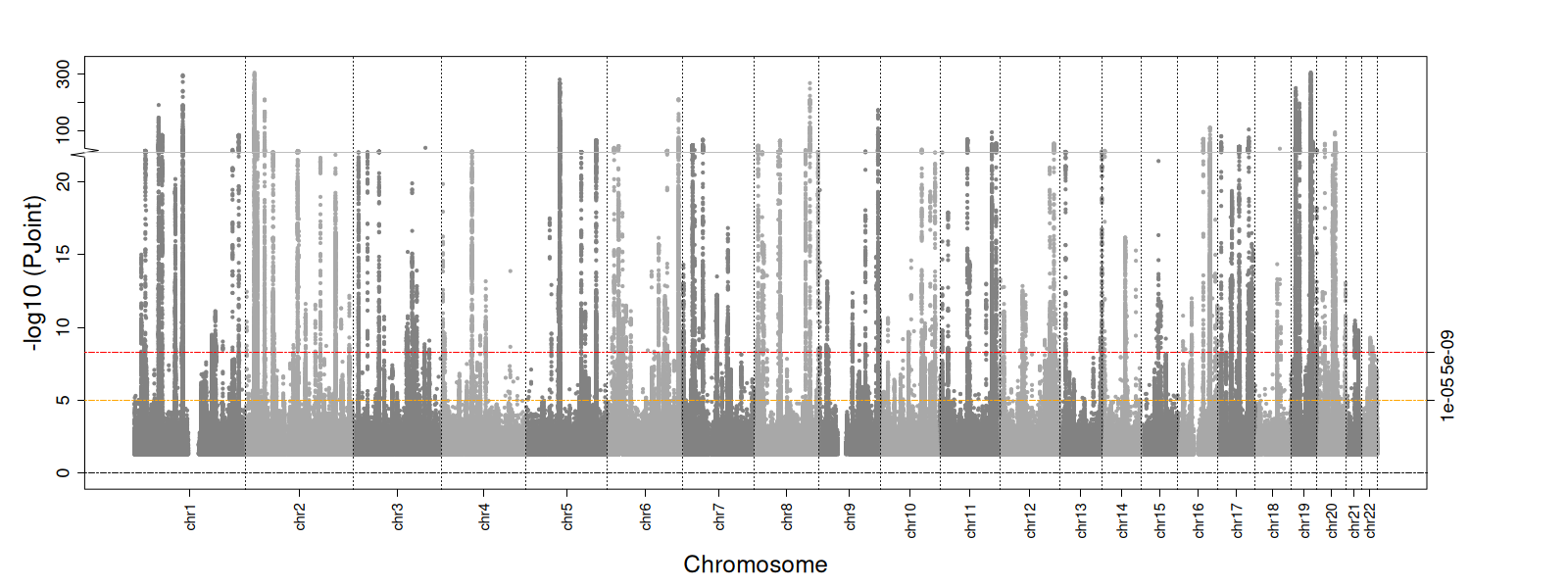 | 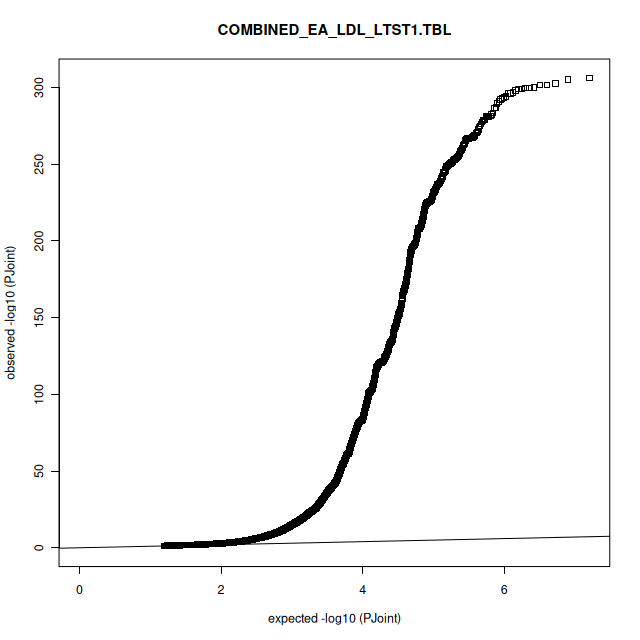 |
| LDL-c and STST |  |
| 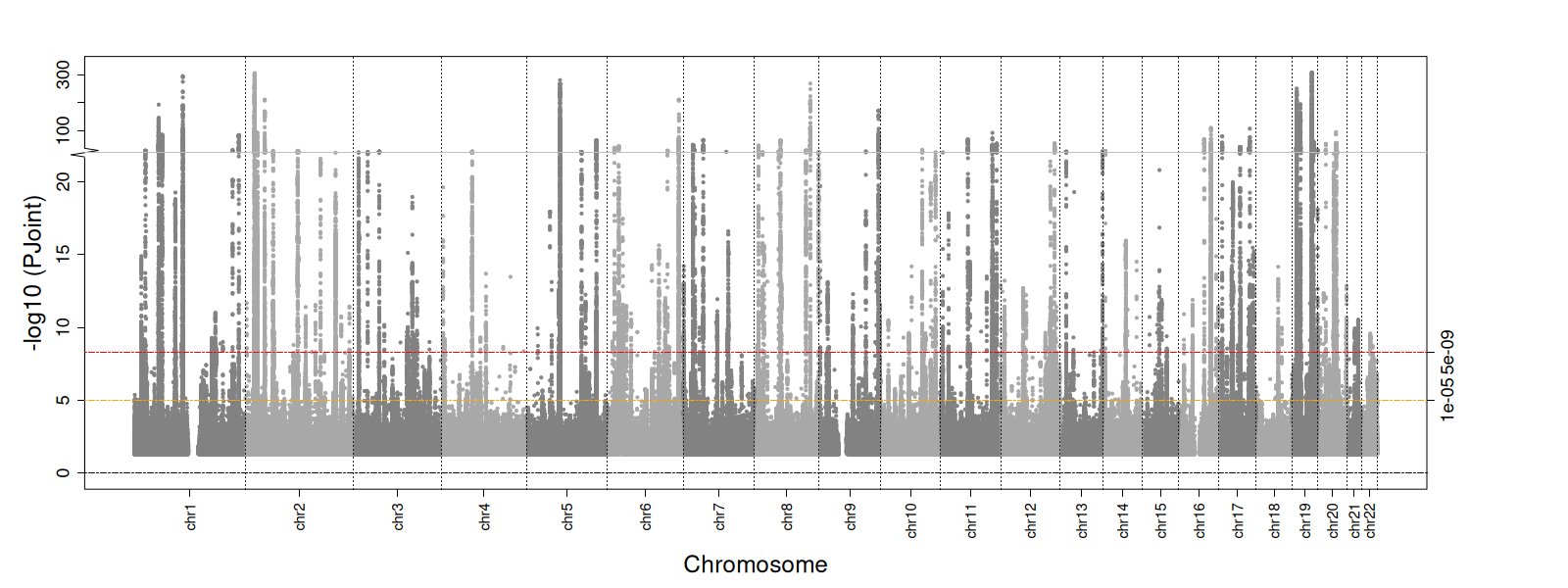 | 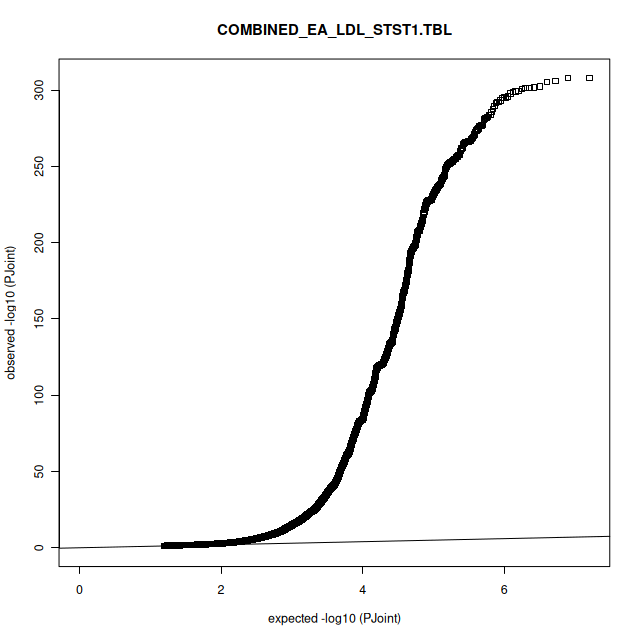 |
| TG and LTST |  |
| 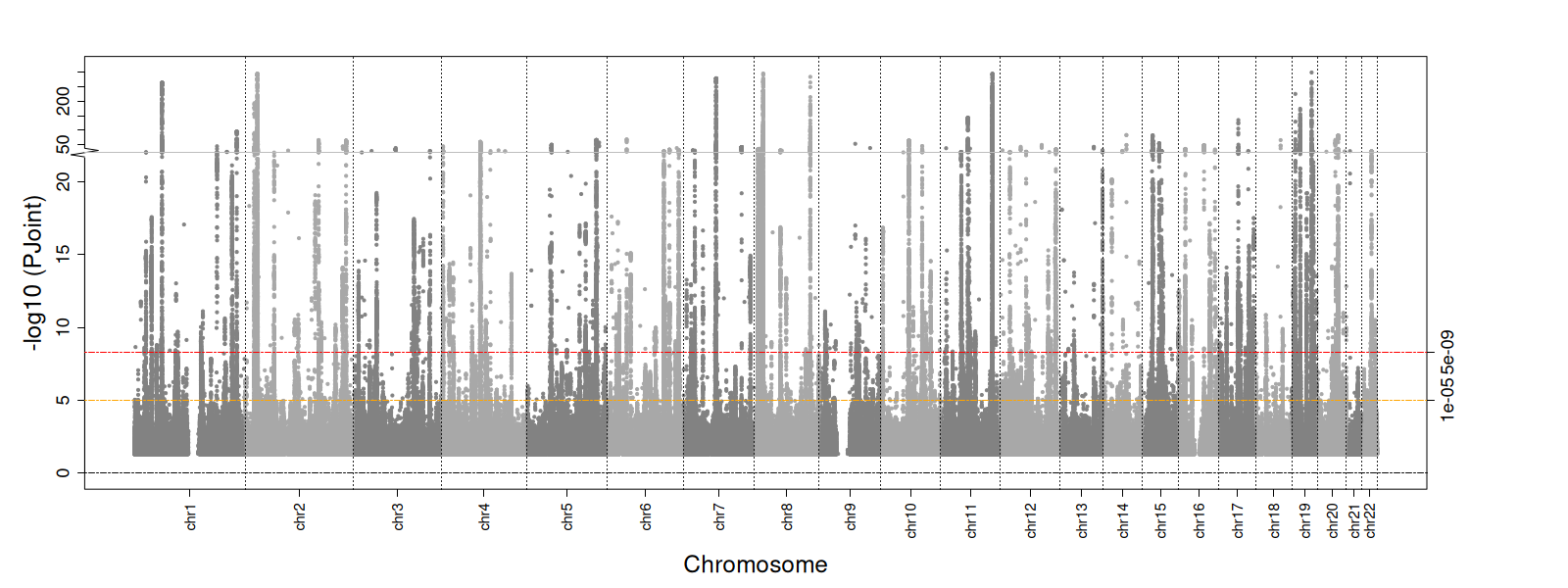 | 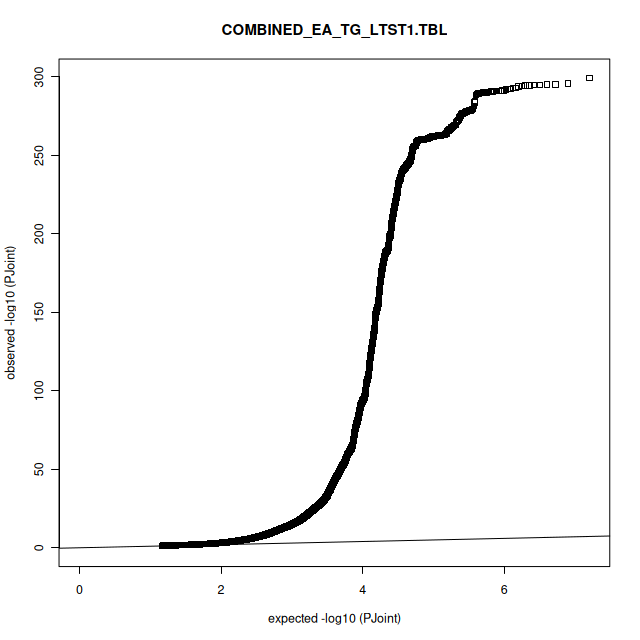 |
| TG and STST |  |
| 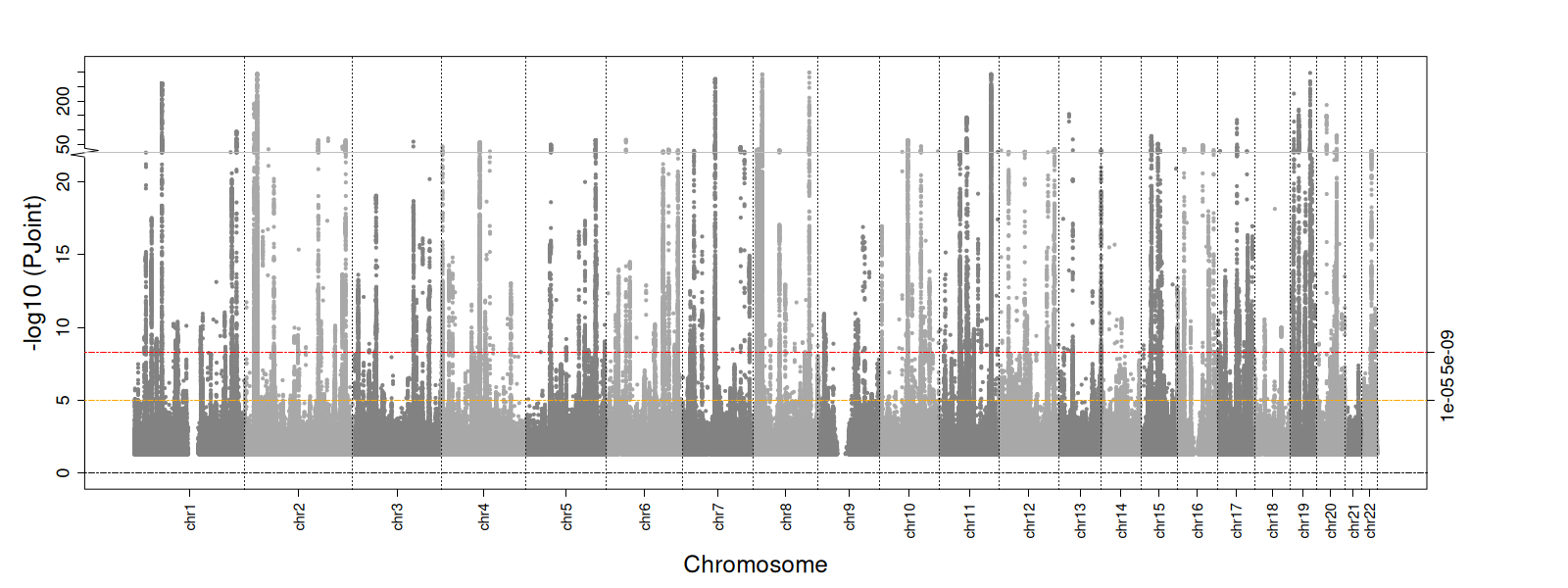 | 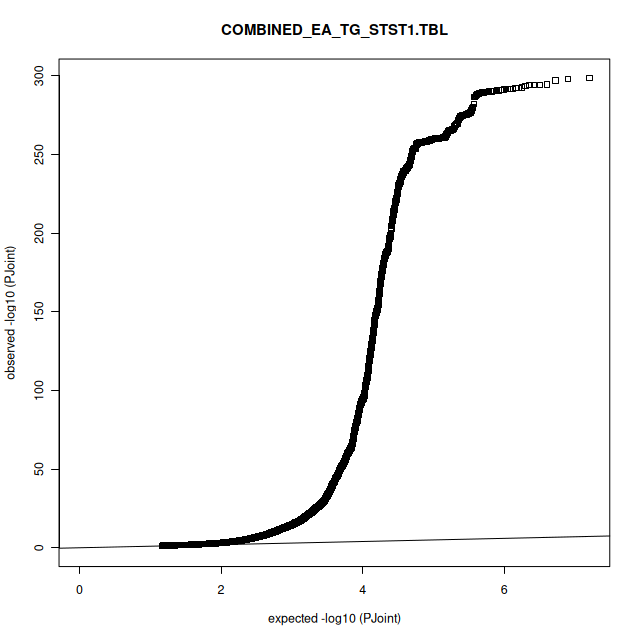 |

**Supplementary Table 7: -log(p) and accompanying Q-Q plots for the African two-df freedom sleep interaction analyses**

| HDL-c and LTST |  |
| --- | --- |
| 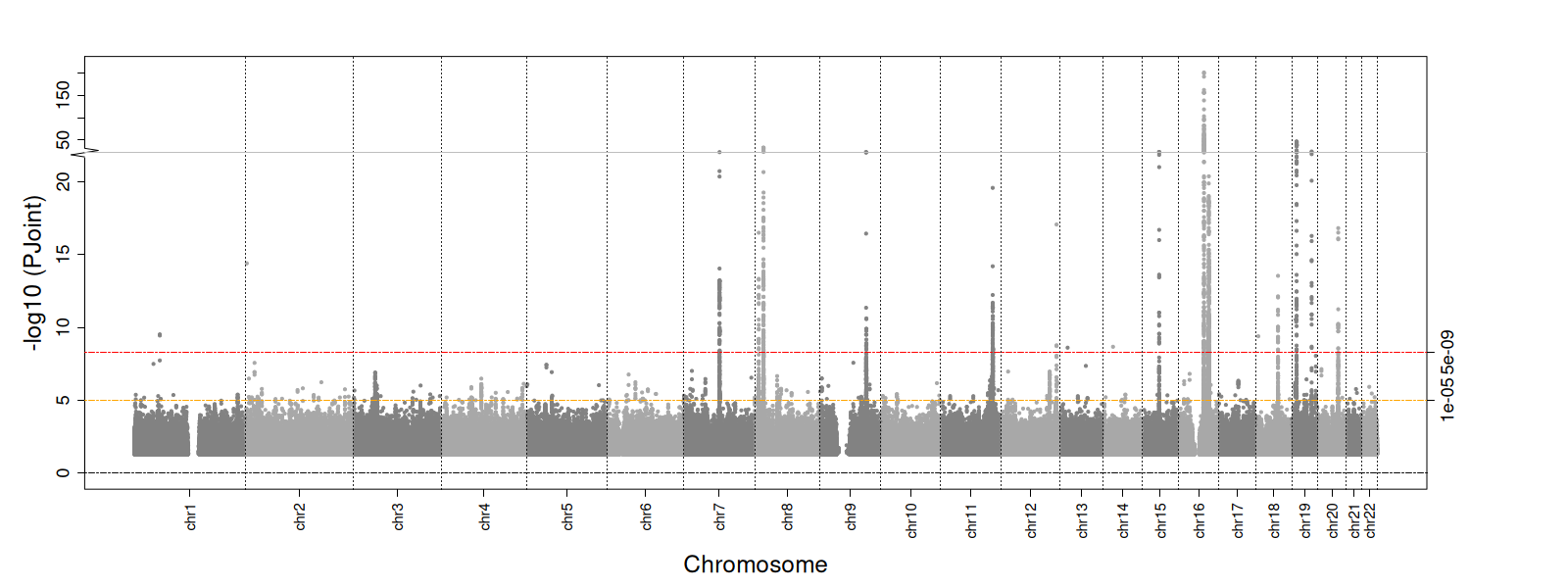 | 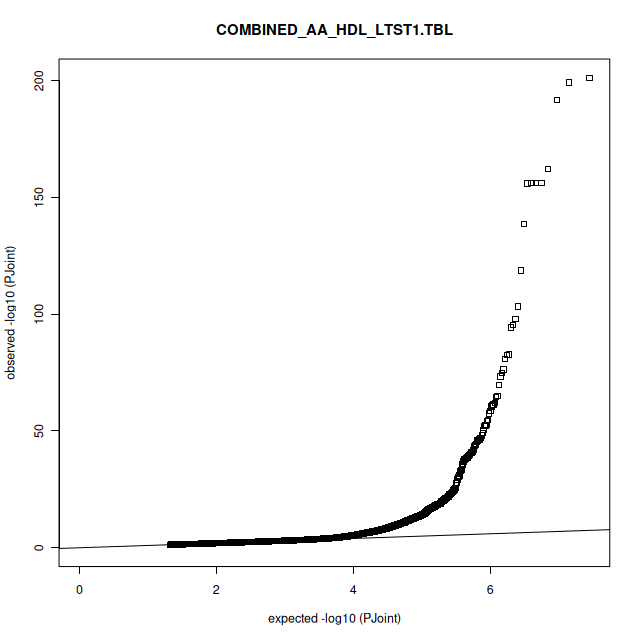 |
| HDL-c and STST |  |
| 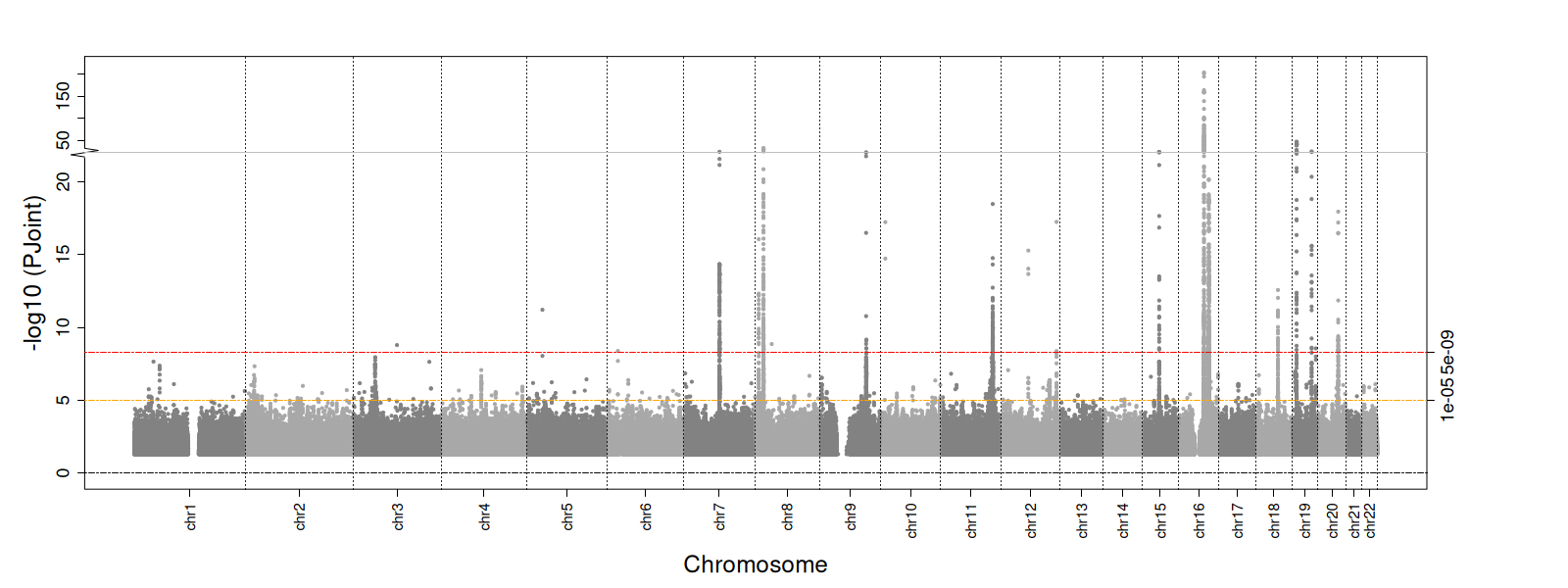 | 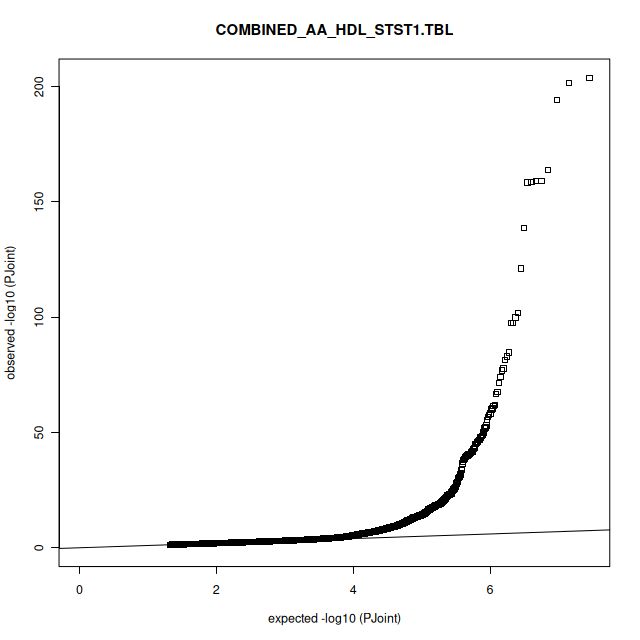 |
| LDL-c and LTST |  |
| 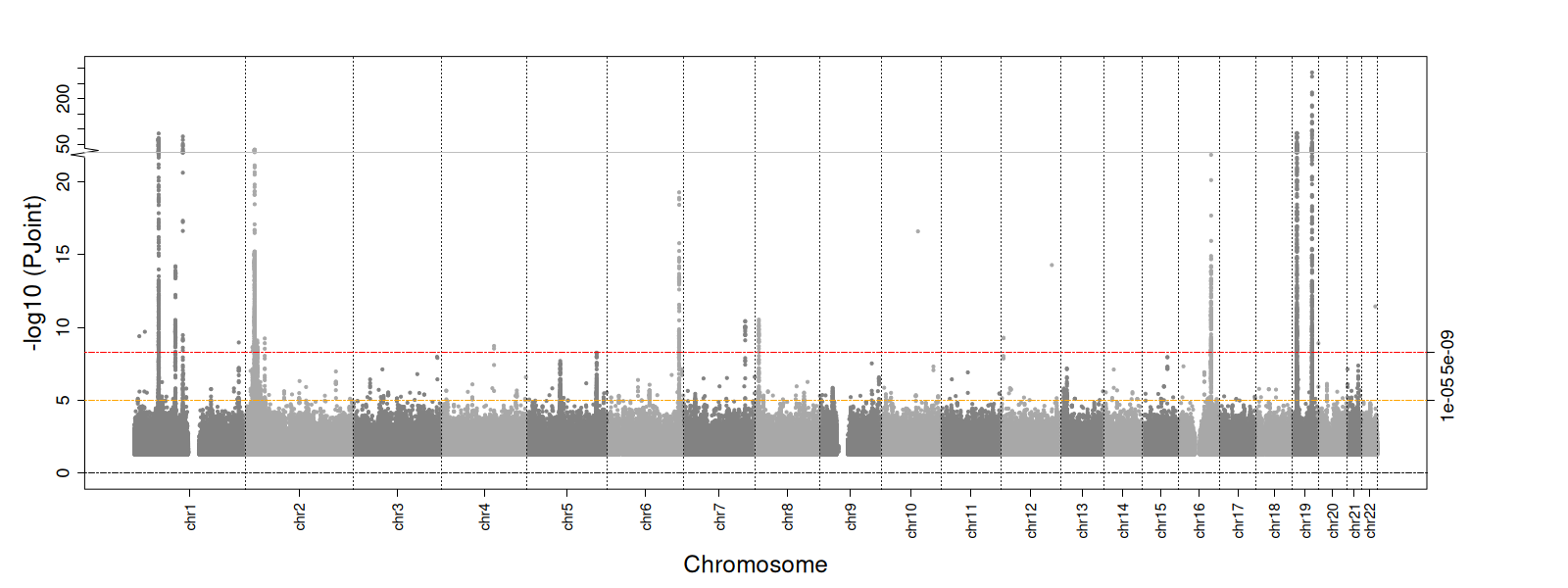 | 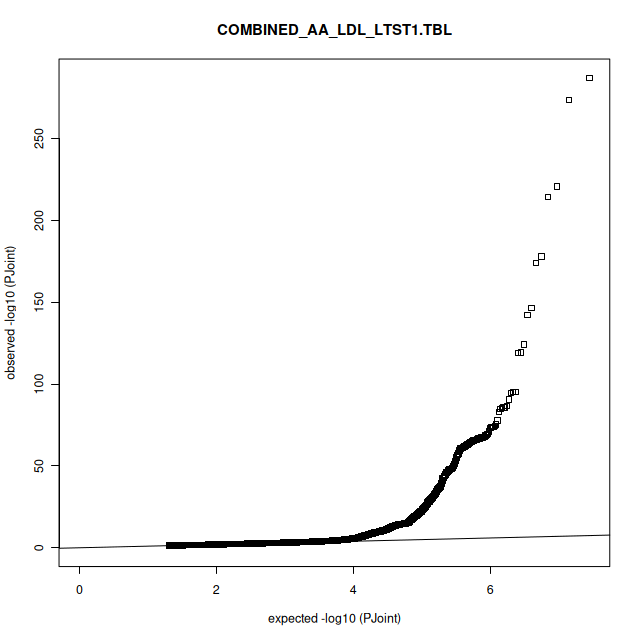 |
| LDL-c and STST |  |
| 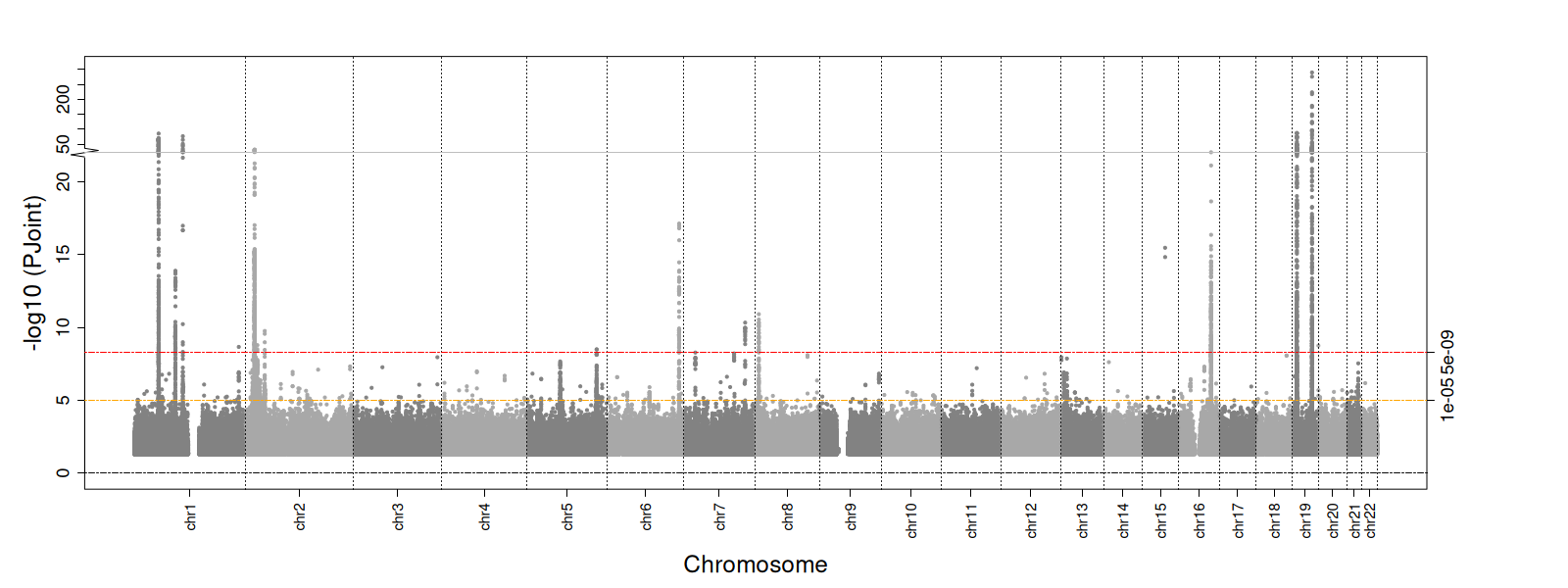 | 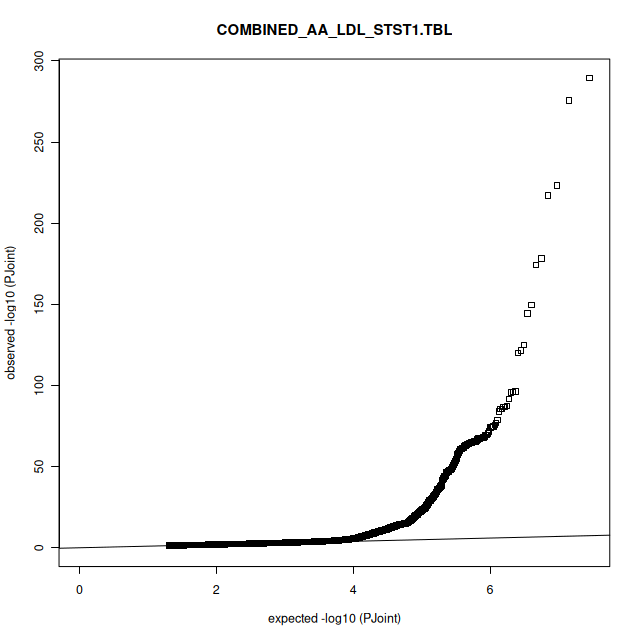 |
| TG and LTST |  |
| 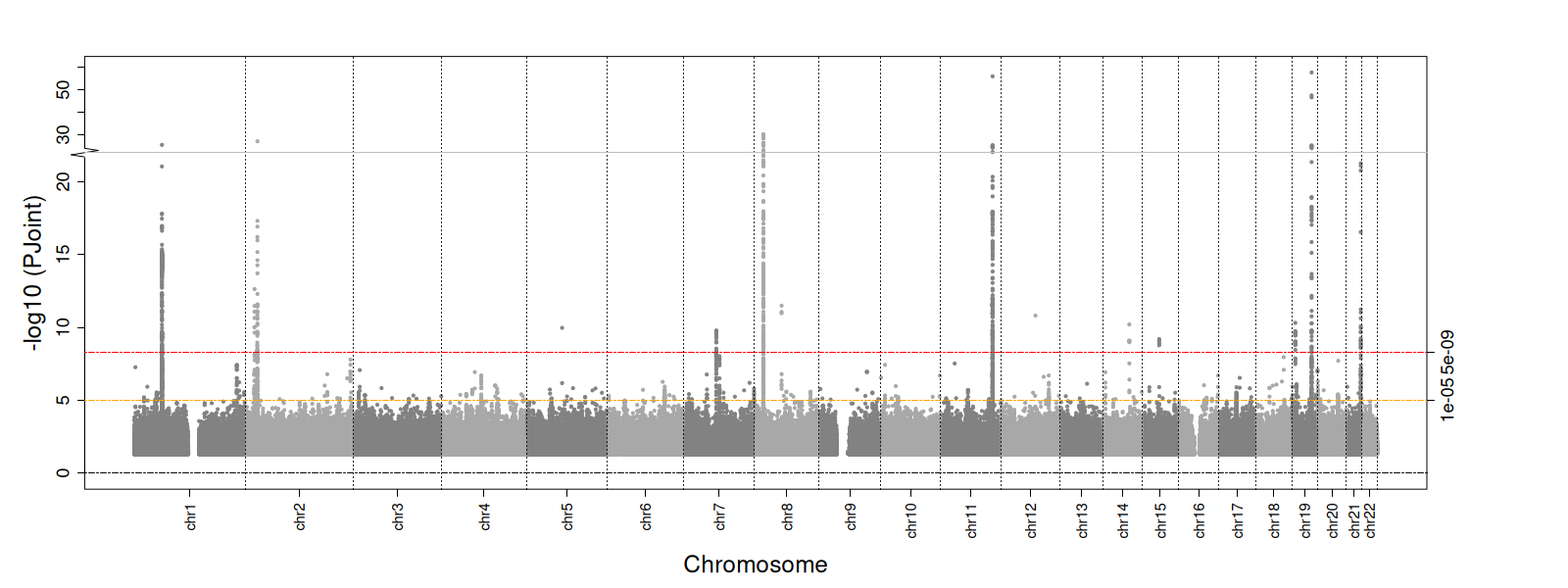 | 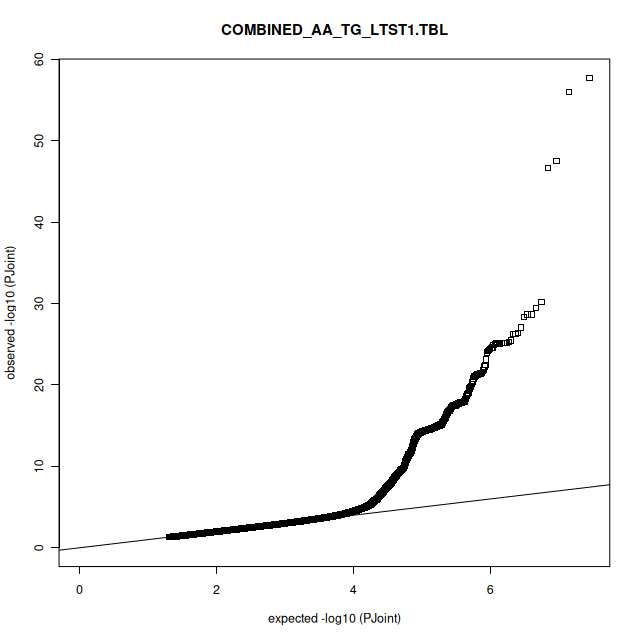 |
| TG and STST |  |
| 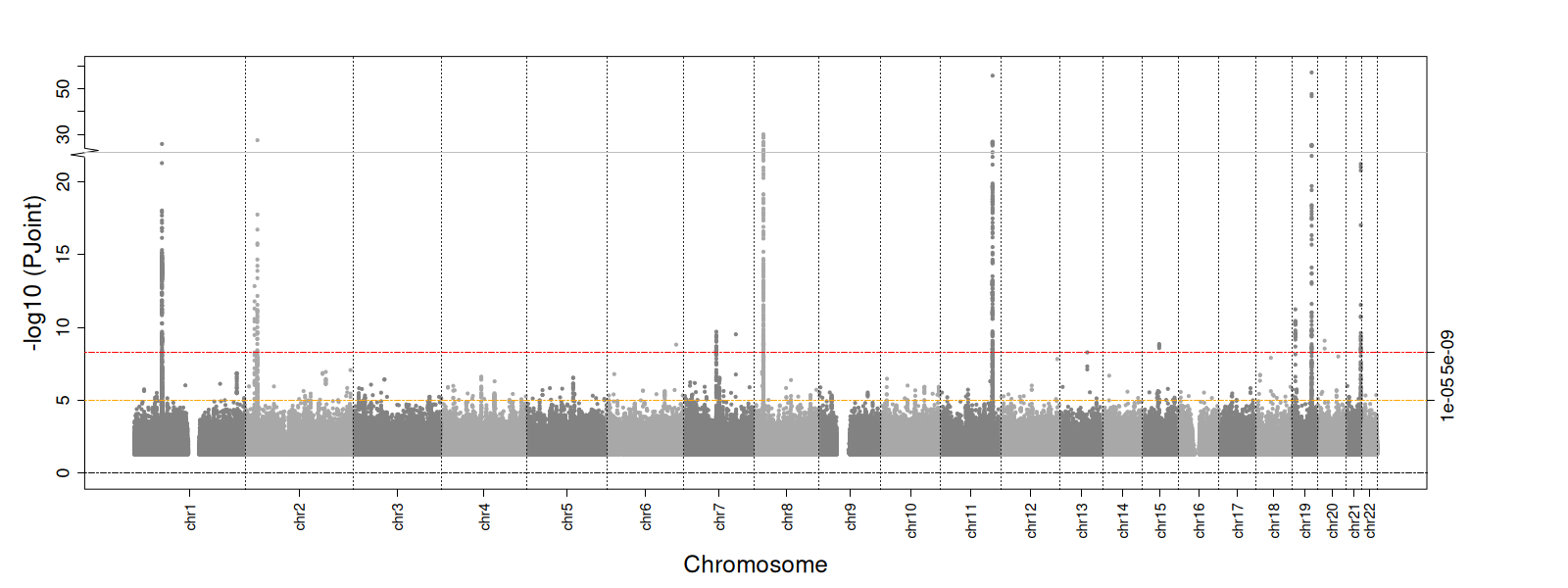 | 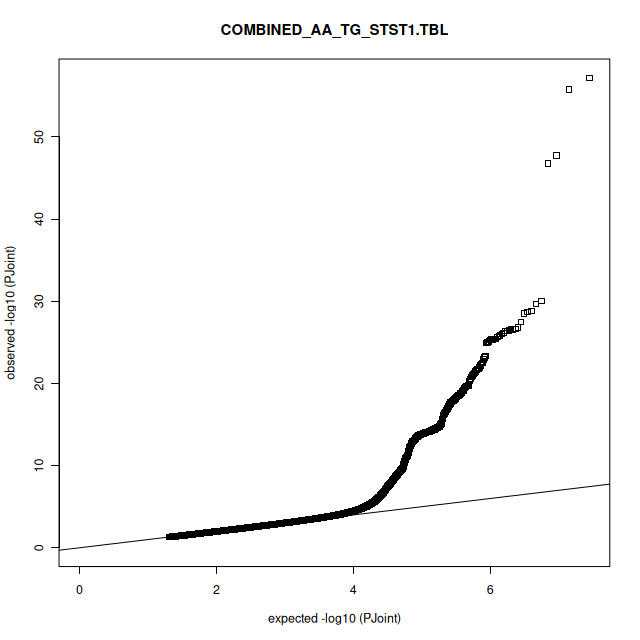 |

**Supplementary Figure 8:** **Regional plots for additional variants identified through the two-degree of freedom interaction analyses.** Linkage Disequilibrium (LD) information is presented in the figure through colors (blue represents low LD, yellow represents moderate LD, and red high LD). For variants colored in grey, no LD information was available.

A) 20:51830403:A_G

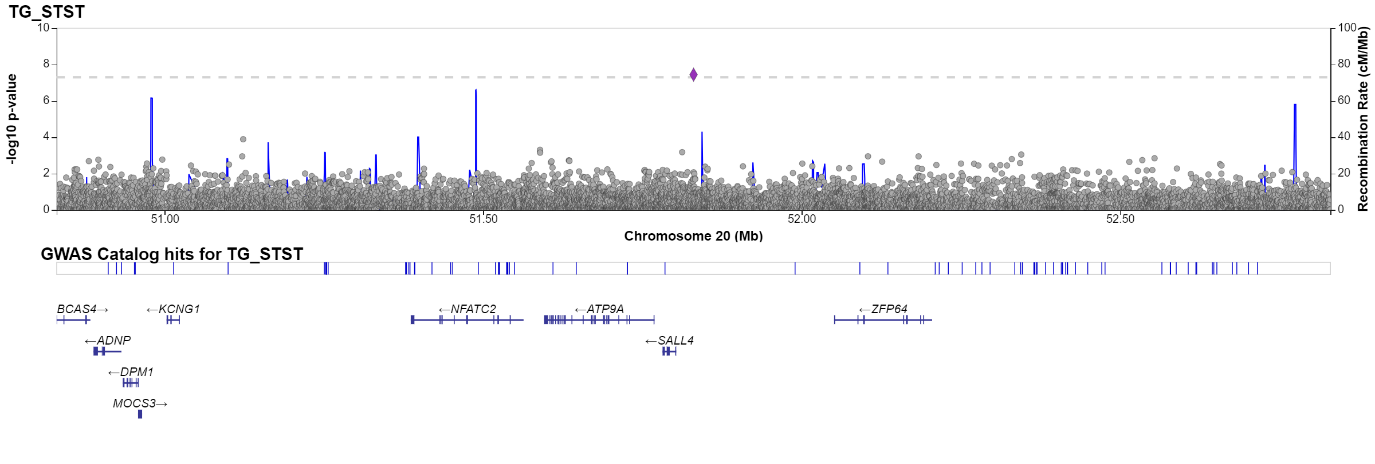

B) 11:13058160:C_T

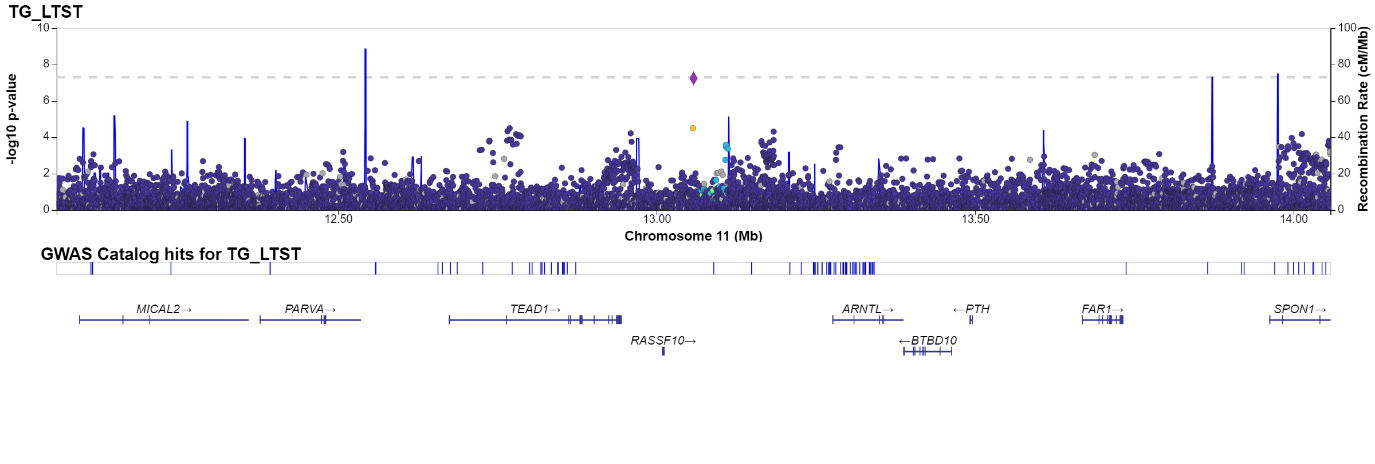

C) 10:97769146:A_G

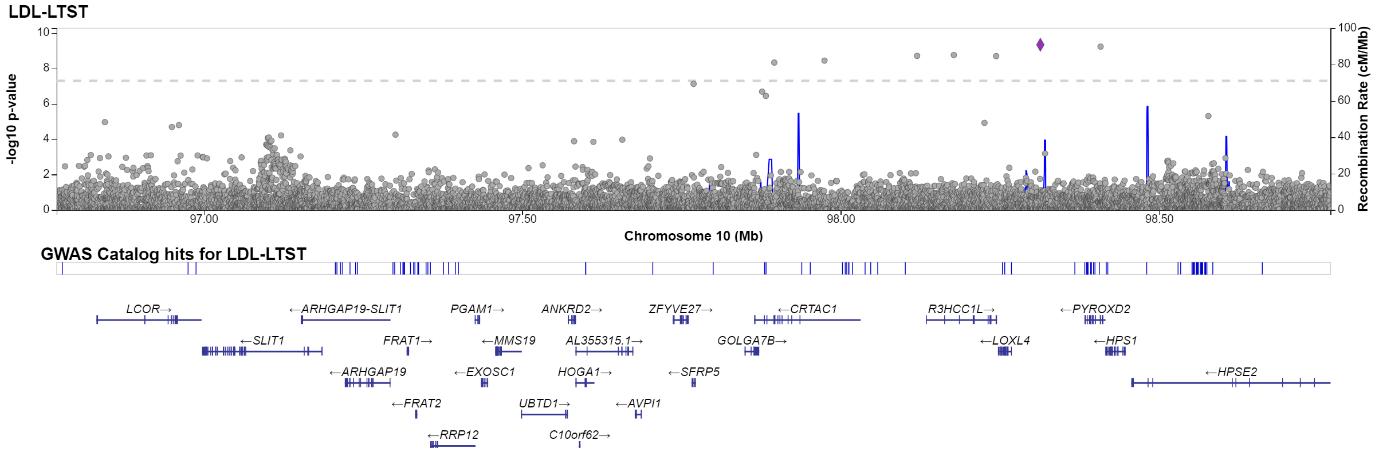

D) 21:35272725:A_T

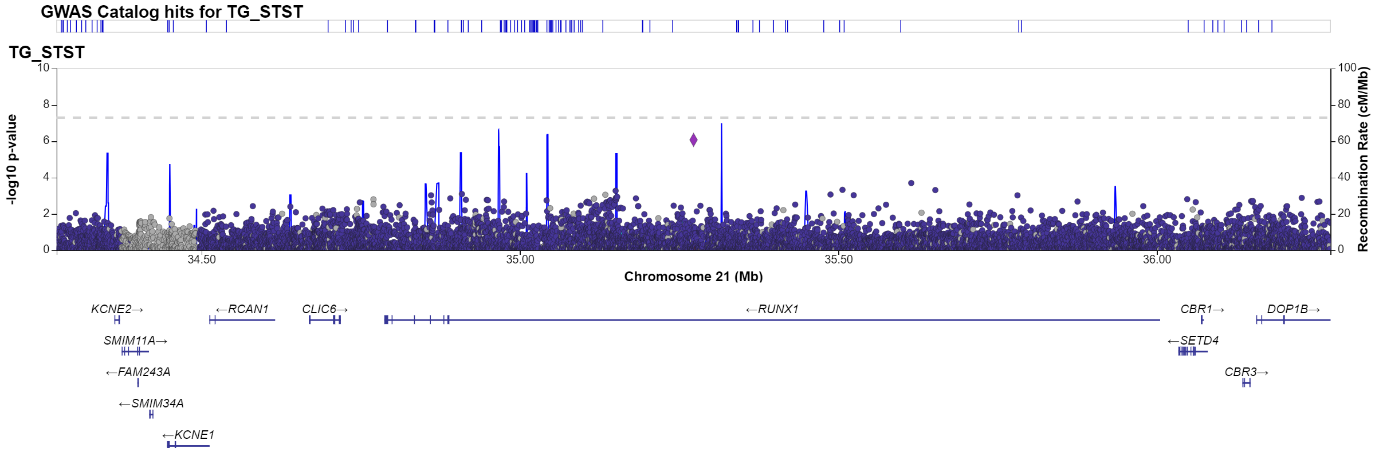

E) 18:55378517:A_T

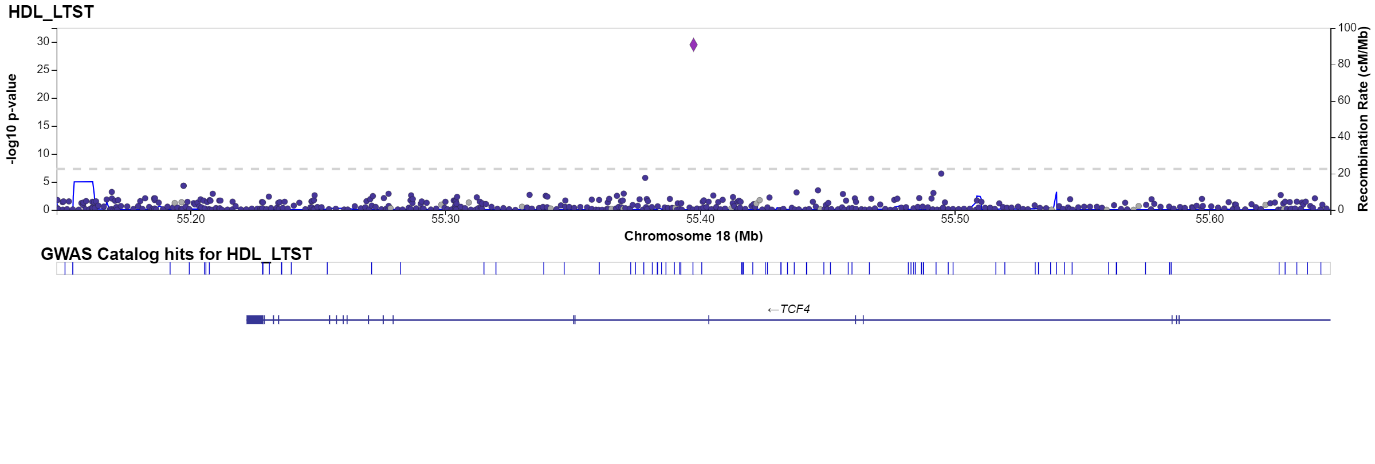

F) 2:40094191:A_T

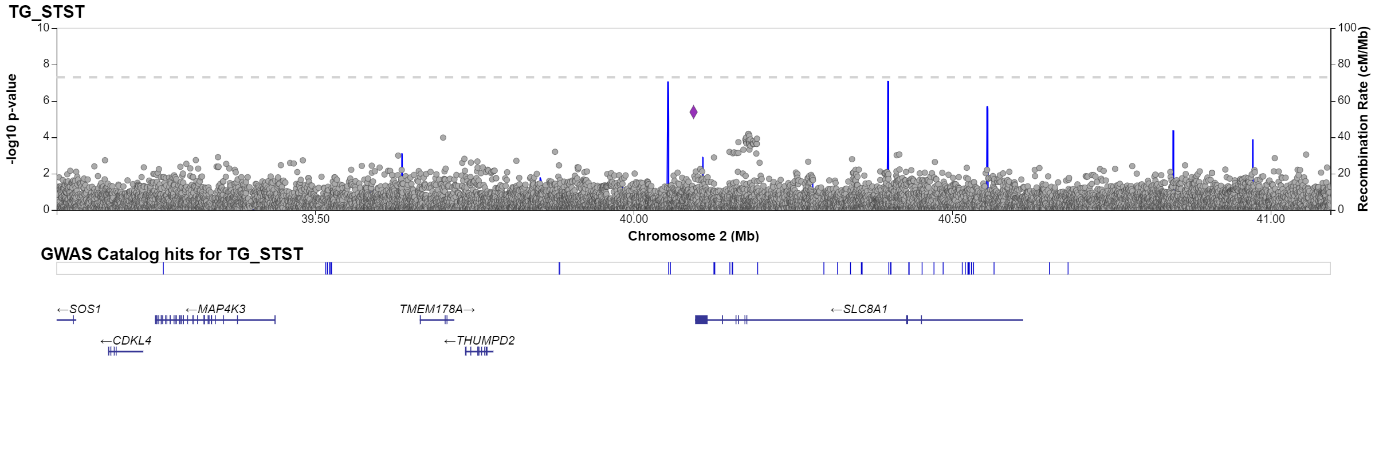

G) 20:23353740:A_G

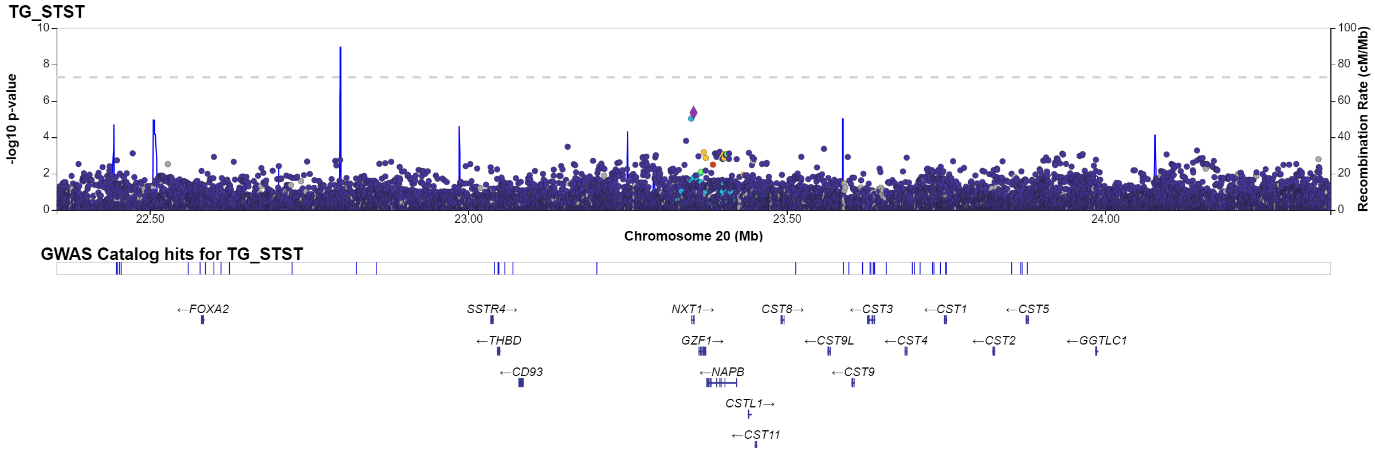

**Supplementary Figure 9**: Gene-set enrichment results for the short total sleep time-triglyceride results. Information on the proportion of overlapping genes in the gene set, enrichment p-value (on a -log10 scale) and information on the genes included in the gene set is presented.

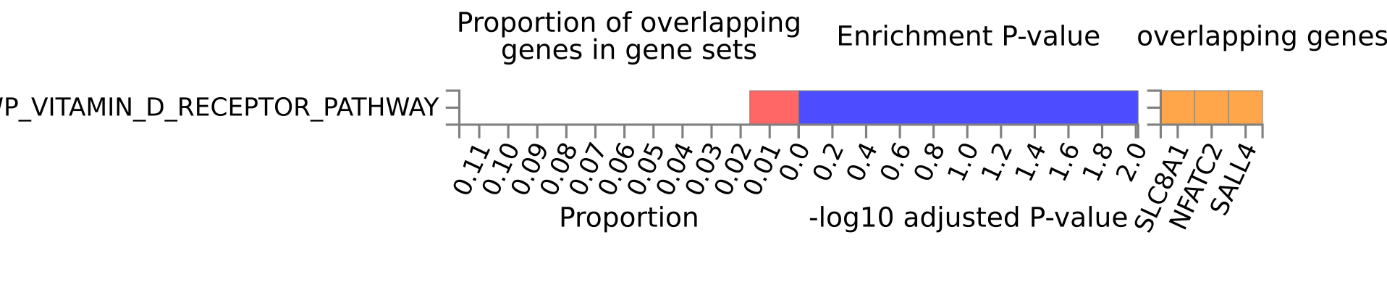
